# Efficacy and safety of esketamine for “treatment resistant depression”: registered report for a Systematic Review with an Individual Patient Data Meta-analysis of Randomized, Double-Blind, Placebo-Controlled Trials

**DOI:** 10.1101/2025.07.17.25331696

**Authors:** Florian Naudet, Claude Pellen, Liviu A Fodor, Chiara Gastaldon, Corrado Barbui, Erick H Turner, Estelle Le Pabic, Ioana A Cristea

## Abstract

**Background:** In 2019, the FDA and EMA approved intranasal esketamine for treatment-resistant depression (TRD). The current study re-evaluated its efficacy and safety.

**Methods:** This registered report presents a systematic review and individual patient data (IPD) meta-analysis of double-blind, randomised, placebo-controlled trials (RCTs) assessing intranasal esketamine for TRD. Two reviewers independently screened studies from multiple databases and obtained IPD via the Yale Open Data Access Project. Two independent researchers selected studies and assessed risk of bias. The primary outcome was the Montgomery-Åsberg Depression Rating Scale (MADRS) score at ≥4 weeks in initiation trials, benchmarked against the 6.5-point clinical significance threshold used in the design of pivotal trials. Evidence certainty was rated using GRADE. Secondary outcomes included additional efficacy and safety endpoints. A two-step IPD meta-analysis was conducted, with separate analyses by trial phase (initiation vs. continuation) and treatment type (combination vs. monotherapy). A one-stage meta-analysis explored moderators (e.g. age and resistance level).

**Results:** We re-analysed IPD from 7 RCTs including 1,505 patients. In five initiation trials of esketamine plus an antidepressant, esketamine reduced MADRS scores at 4 weeks (mean difference (MD) = –2.94, 95% CI [–5.39 to –0.48]; GRADE: moderate certainty). A continuation trial showed reduced relapse risk (HR = 0.38 [0.26–0.57]), though FDA concerns about one centre could not be addressed due to data privacy restrictions. A monotherapy trial (aggregate data only) showed a larger effect (MD = –6.32 [–8.62 to –4.03]), but there were concerns over selection bias and unblinding. Esketamine increased sedation (RR = 3.70 [2.02–6.78]), dissociation (RR = 2.36 [2.10–2.65]), and adverse events (IRR = 3.91 [2.37– 6.45]), with no increase in serious adverse events (IRR = 1.35 [0.54–3.40]). No treatment effect moderation was found by age or resistance level, with most patients displaying low-stage TRD.

**Conclusions:** Based on the IPD used for approval, esketamine in combination with an antidepressant showed an advantage over placebo that was statistically significant but small. The clinical relevance of this benefit is unclear, particularly given the risks of adverse events.

**Funding:** French Programme Hospitalier de Recherche Clinique (PHRC).

**Registration:** PROSPERO: CRD42021290721

## BACKGROUND

In 2019, the Food and Drug Administration (FDA) approved a nasal spray formulation of esketamine for “treatment resistant depression” (TRD). Esketamine’s licensing application was granted “breakthrough designation”, an expedited review process for drugs “intended to treat a serious condition and preliminary clinical evidence indicates that the drug may demonstrate substantial improvement over available therapy on a clinically significant endpoint(s)” [1]. The European Medicines Agency EMA promptly followed and approved esketamine [2]. However, the evidence base for its use proved to be controversial, as outlined by Schatzberg [3], Gastaldon and colleagues [4] and in various comments by our group in the *Lancet Psychiatry* [5–8].

The randomised controlled initiation trials (RCTs) barely demonstrate short-term benefits and did not investigate long-term efficacy of esketamine in that two [9, 10] out of the 3 pivotal initiation RCTs [9–11] failed to demonstrate that esketamine was superior to placebo [5]. For the first time for the FDA’s Division of Psychiatry Products, a continuation RCT [12] was counted as a second positive pivotal trial. The definition used to define TRD (i.e., a depressive episode with inadequate response to at least two trials of antidepressants—even two antidepressants of the same class (e.g. two SSRIs) [7] represents a very low stage of “resistance” and may be not relevant to day-to-day clinical practice. More concerningly, the FDA described in its assessment some possible data integrity issues for two trials [5]. For the “negative” initiation RCT involving elderly participants, the FDA reviewer described an “unusual response curve shift” at post-test, “discrepancies between the locked datasets”, and “reported protocol violations” [13]. These issues raise particular concerns because the trial involved a vulnerable population. For the continuation study, the FDA reported that “one site in Poland drives the overall study result due to a 100% rate of placebo arm relapses” [13]. After our pieces drew attention to this issue, the sponsor reported a sensitivity analysis [14]: the P-value changed from highly significant to “barely significant” [8]. However, the sponsor did not explain why the FDA used the word “drives”, implying that its result was nonsignificant when this center was removed, nor why the FDA reported slightly different numbers of relapsed patients [8].

In August 2020, the FDA approved a supplemental New Drug Application for esketamine nasal spray (SPRAVATO) for the treatment of depressive symptoms in adults with major depressive disorder and acute suicidal ideation or behavior. However, even the sponsor’s press release admits that “effectiveness of SPRAVATO® in preventing suicide or in reducing suicidal ideation or behavior has not been demonstrated” [15], again suggesting a lack of strong evidence for its use in this indication. In contrast, a pharmacovigilance analysis showed that suicide ideation might be increased in people taking esketamine [16].

In UK, the National Institute for Health and Care Excellence (NICE), based on its independent review, did not recommend its use [17]. In France, the “Commission de la transparence” expressed a favorable opinion for reimbursement in combination with a Selective Serotonin Reuptake Inhibitor (SSRI) or a Serotonin and Norepinephrine Reuptake Inhibitor (SNRI) for the treatment of treatment-resistant major depressive disorder. However, this opinion was restricted to a subgroup of patients. This included adults under 65 years of age a) who had not responded to at least two different treatments with antidepressants in the current severe depressive episode and b) who had not responded to electroconvulsive therapy (or had contraindication, or lack of access or refusal). The commission expressed an unfavorable opinion regarding reimbursement in remaining indications and stressed that there was no clinical added value in the therapeutic strategy [18].

Following approval, a single-blind, randomized, active-controlled trial was conducted in which raters were unaware of participant allocation. The study evaluated the efficacy of flexible doses of esketamine nasal spray compared to extended-release quetiapine, both administered alongside an SSRI or SNRI, in patients with treatment-resistant depression (TRD). At 8 weeks, esketamine nasal spray demonstrated superior efficacy [19]. However, the study was criticized [20, 21] for its design and analytical choices, which may have exaggerated the observed differences between the groups, despite the arguably small difference in means (−2.2 points [95% CI, −3.6 to −0.8]).

Based on these divergent viewpoints, concerns surrounding data integrity, and doubts about esketamine’s efficacy in TRD patients, an independent re-appraisal of the evidence from pivotal trials is urgently warranted. The new norm, where data sharing for randomized controlled trials becomes standard [22], where data sharing for randomized controlled trials becomes standard, offers a unique opportunity for independent researchers to re-analyse and synthesize (e.g. individual patient data [IPD] meta-analyses) evidence from RCTs [23].

Esketamine’s sponsor, Johnson & Johnson, has adopted a policy that upholds sharing clinical trial data, including both recently completed, as well as older, trials (https://yoda.yale.edu/jj-available-data) via the Yale Open Data (YODA) project [24].

Data sharing holds promise in such a controversial case to re-evaluate esketamine’s benefit-risk ratio, in the spirit of personalized medicine, in clinically relevant subgroups, specifically patients with different levels of treatment “resistance” and vulnerable populations, especially the elderly. Importantly, a pharmacovigilance analysis found some specific patients reported more serious adverse events (women, obese people, people with comorbidities and with poly-pharmacy) [16].

The main objective is to independently reappraise the efficacy of esketamine in TRD using an individual participant data meta-analysis methodology. The secondary objectives are to reappraise the safety of esketamine in the treatment of TRD and to explore moderating factors of esketamine efficacy and safety, including level of treatment resistance, patient age, site-specific effects, among others.

## METHODS

### Design

We conducted a systematic review and meta-analysis of individual patient data derived from double-blind, randomised, placebo-controlled [placebo or active placebo] trials for the indication of TRD. This study follows a registered report format, with in-principle acceptance granted on 2021-11-15. The protocol was registered on the Open Science Framework (OSF) on 2021-11-15, where the full protocol is publicly available, and subsequently on PROSPERO (CRD42021290721) on 2021-12-11. The reporting adhered to the PRISMA [25] and PRISMA-IPD [25] guidelines.

### Participants, interventions, comparators, outcomes, and study design (PICOS)

We considered studies that include all patients with TRD meeting the inclusion criteria of esketamine’s development program for this indication. Following the definition used in this program, TRD refers to a depressive episode with inadequate response to at least two antidepressant trials of adequate doses and duration. This definition encompasses both low and high levels of resistance, as classified by Thase and Rush’s staging method for treatment-resistant depression (**see e-Box 1 in Web appendix 1**) [26]. There was no age limit. In terms of strategy, the systematic review search criteria included all studies of esketamine for the primary treatment of depression. The IPD analysis was then restricted to participants/patients with TRD within the included studies (i.e., have failed > 2 adequate antidepressant trials).

Included were studies that compared the use of intranasal esketamine (with the following approved doses: 56 mg, 84 mg, or “flexible” doses) to a placebo. In the trials, placebo was described as a solution with a bittering agent added to simulate the taste of the esketamine solution. All randomised controlled trials (initiation and continuation trials) were considered.

### Outcomes

The primary outcome was the MADRS score (continuous outcome, absolute score at study endpoint) assessed after at least 4 weeks (the end of treatment in the pivotal initiation trials / it was expected to be 4 weeks but could be longer depending on the studies selected for the meta-analysis). To take into account the possible rapid effect of esketamine, we also studied the MADRS score at week 1 as a secondary outcome.

All of the following secondary outcomes were assessed at end of treatment as well as at the last follow-up visit post-treatment (expected to be 24 weeks) in initiation trials. These were assessed at the end of the study in continuation trial (this was expected to be over 500 days).

Secondary outcomes included efficacy outcomes, namely (a) suicide/suicide attempt (binary outcome); b) suicidal ideations (binary outcome); c) MADRS (continuous outcome, similar to the primary outcome but, here using the last follow-up visit post-treatment); d) remission (binary outcome, usually MADRS<=12); e) Sheehan Disability Scale (continuous outcome); f) PHQ-9 (continuous outcome); g) relapse (censored outcome) for continuation trials, and safety outcomes, namely a) serious adverse events (count outcome, i.e. number of events per patient); b) dropout for any cause (binary outcome); c) dropout due to adverse events (binary outcome); d) Adverse events (count outcome i.e. number of events per patient); e) Blood pressure (continuous repeatedly measured outcome); f) Dissociation (binary outcome) g) Sedation (binary outcome).

### Search strategy

Systematic searches were conducted using PubMed, the Cochrane library, Embase, ClinicalTrials.gov, Clinicaltrialsregister.eu, FDA, EMA and the manufacturer website. All records were managed (and deduplicated) using Endnote. **e-Table 1** (in **Web appendix 1**) provides a detailed overview of the search strategy. Following completion of the analysis, an update was conducted utilizing findings from an ongoing Cochrane review [27] as well as a recent meta-analysis [28].

**Table 1:** Summary of included studies evaluating the efficacy of esketamine in the treatment of major depressive disorder. * For the re-analyses, all doses were considered; however, for the meta-analyses, only doses of 56 mg, 84 mg, or flexible dosing were included. a: The protocol specified the full Analysis Set (FAS) with n=229 (2 patients were not treated with the study treatment, and 2 other patients did not receive the oral antidepressant treatment). b: Due to Good Clinical Practice issues identified at a study site during an on-site audit, 9 subjects from this site were excluded from all analysis sets. The data of these 9 patients are available on Yoda, with no means of identification. c: Due to Good Clinical Practice issues identified at a study site during an on-site audit, 1 subject from this site was excluded from all analysis sets. The data of this patient are available on Yoda, with no means of identification. d: The protocol specified the full Analysis Set (FAS) with n=250 (2 patients did not receive the oral antidepressant treatment). e: 226 patients were randomized to esketamine and 250 to placebo.

| Study identification (NCT) | Participants | Esketamine dose* | Study duration (blinded) | Number of participants | Mean age | Sex (women) | Details of observed major deviations |
| --- | --- | --- | --- | --- | --- | --- | --- |
| **Initiation trials (combination with an antidepressant)** |  |  |  |  |  |  |  |
| NCT01998958<br>- panel A<br>- panel B<br>(unpublished) | Patients (aged 20 to 64 years) with TRD | 28, 56 and 84 mg (panel A)<br>14, 56 mg (panel B) | 8 days | Panel A:<br>- Esketamine: 34<br>- Placebo: 33<br>Panel B:<br>- Esketamine: 20<br>- Placebo: 21 | 42.9 ± 10.1 (panel A)<br>42.2 ± 8.0 (panel B) | 38/67 (56.7%, panel A)<br>17/41 (41.5%, panel B) | There were 7 major deviations. In panel A, 1 patient was enrolled but did not meet the inclusion criteria, and 4 patients had major deviations categorized as 'other,' with no further details provided. In panel B, 2 patients had major deviations categorized as 'other,' with no further details provided. |
| NCT02918318 | Patients (aged 20 to 64 years) with TRD | 28, 56 and 84 mg | 28 days | - Esketamine: 122<br>- Placebo: 80 | 43.4 ± 10.4 | 96/202 (47.5%) | There were 25 major deviations: 5 patients received disallowed concomitant treatments, 10 patients received the wrong treatment or an incorrect dose, and 9 patients had deviations categorized as 'other' (including 1 patient with 2 deviations). No further details were provided. |
| NCT02417064 | Patients (aged 20 to 64 years) with TRD | 56 and 84 mg | 28 days | - Esketamine: 229 (233 in the published paper)<sup>a</sup><br>- Placebo: 113 | 46.7 ± 11.2 | 241/342 (70.5%) | There were 62 major deviations: 28 patients entered but did not satisfy criteria (of those, one had 2 deviations), 9 patients received disallowed concomitant treatments, 1 patient received wrong treatment or incorrect dose, 2 patients developed withdrawal criteria but not withdrawn, and 21 patients had deviations categorized as 'other'. No further details were provided. |
| NCT02418585 | Patients (aged 20 to 64 years) with TRD | Flexible (56 or 84 mg) | 28 days | - Esketamine: 118 (114 in the published paper)<sup>b</sup><br>- Placebo: 114 (109 in the published paper)<sup>b</sup> | 46.3 ± 11.8 | 142/232 (61.2%) | There were 14 major deviations: 3 patients received disallowed concomitant treatments, 3 patients received the wrong treatment or an incorrect dose and 8 patients had deviations categorized as 'other'. No further details were provided. |
| NCT02422186 | Patients (≥65 years old) with TRD | Flexible (28, 56 or 84 mg) | 28 days | - Esketamine: 72<br>- Placebo: 66 (65 in the published paper)<sup>c</sup> | 70.3 ± 4.5 | 86/138 (62.3%) | There were 22 major deviations: 6 patients received the wrong treatment or an incorrect dose, 6 entered but did not satisfy criteria and 10 had deviations categorized as 'other'. No further details were provided. |
| NCT03434041 | Patients (aged 18–64 years) with TRD | Flexible (56 or 84 mg) | 28 days | - Esketamine: 124 (126 in the published paper)<sup>d</sup><br>- Placebo: 126 | 36.4 ± 12.2 | 113/250 (45.2%) | There were 21 major deviations: 10 patients received disallowed concomitant treatments, 2 patients received the wrong treatment or an incorrect dose, 2 patients entered but did not satisfy criteria and 7 patients had deviations categorized as 'other'. No further details were provided. |
| **Initiation trial (monotherapy)** |  |  |  |  |  |  |  |
| NCT04599855 | Patients (over 18 years old) with TRD | 56 and 84 mg | 28 days | Esketamine: 171 (226 randomised)<br>- Placebo: 185 (250 randomised) | 45.3 ± 14.0 | 284/476 (59.7%) | No additional information is available, as we relied exclusively on results reported in ClinicalTrials.gov and in the publication. |
| **Continuation trial (combination with an antidepressant)** |  |  |  |  |  |  |  |
| NCT02493868 | Patients (aged 18–64 years) with TRD | Flexible (56 or 84 mg) | Time to event | Remitters:<br>- Esketamine: 90<br>- Placebo: 86<br>Responders:<br>- Esketamine: 62<br>- Placebo: 59 | Remitters:<br>46.9 ± 11.6<br>Responders:<br>47.9 ± 10.5 | Remitters:<br>118/175<br>(67%)<br>Responders:<br>79/121<br>(65.3%) | There were 5 major deviations: 2 patients developed withdrawal criteria but not withdrawn, 1 patient received the wrong treatment or an incorrect dose, 1 patient received disallowed concomitant treatments and 1 patient had deviations categorized as 'other'. No further details were provided. |

### Data collection and assessment of risk of bias

Selection and coding of the different study characteristics was performed by two independent reviewers (among CP, FN, IAC, and LF) independently from each other. A third reviewer arbitrated in case of disagreement. A data extraction sheet based on the Cochrane Handbook for Systematic Reviews of Interventions guidelines was developed.

Studies appearing to duplicate authors, treatment comparisons, sample sizes and outcomes were checked against one other to avoid any duplicates and to avoid integrating data from several reports on the same study.

For each study included, information was extracted regarding: a) characteristics of the study [year, country, selection criteria, co-treatments (psychological interventions and their type), number of arms, funding]; b) Type of intervention (treatments and comparators, duration). Importantly, mean characteristics of the trial participants and outcome measures were not extracted, as we relied on IPD.

Two researchers (FN and CG) independently assessed each trial (article, trial registration, protocol, SAP and IPD) for risk of bias according to the Cochrane Collaboration tool for assessing risk of bias in its current version, RoB2 [29]. We solved any disagreements by involving a third reviewer (IAC).

A data sharing request (IPD, analytical code, metadata, study reports, statistical analysis plan and any other relevant documents) was sent to the relevant data sharing platforms and/or sponsor and /or authors. Importantly, data and metadata of the pivotal trials were available on the YODA project (https://yoda.yale.edu/jj-available-data). Data requests were submitted on January 5, 2022, and access was granted on February 8, 2022.

### Analysis strategy

The statistical analysis plan (SAP) was finalized on October 21, 2022, prior to any analyses, following an initial review of the data structure for feasibility purpose (**Web appendix 2**). For efficacy and safety outcomes a two-step approach was adopted, while a one-stage approach was used to explore moderators of esketamine efficacy. All the analyses were performed with R (R core Team, 2021).

### IPD integrity and reanalysis

First, we performed a reanalysis of each study following the initial protocol of each study and the International Council for Harmonisation of Technical Requirements for Pharmaceuticals for Human Use [30], as detailed in **e-Box 2** (in **Web appendix 1**). This first approach allowed for in-depth description of any deviations from the protocol and for documentation of the data integrity issues mentioned in the FDA appraisal. For each RCT, we planned to explore a center effect by performing sensitivity analyses removing a center at a time (leave-one-out analysis [31]). This could not be achieved because information about the center was missing from the provided datasets for anonymization purposes.

### Meta-analysis

Then, a fixed effect meta-analysis, or a random effect meta-analysis (in case of heterogeneity defined as an I^2^ index > 25 % and/or detection of significant heterogeneity (p<0.10) detected using Cochran’s Q test statistic, or tau^2^), was used to pool the different efficacy indexes (mean differences or relative risks or odds ratio or hazard ratios) that were derived from these re-analyses. Initiation and continuation designs were analyzed separately. We used the original study analysis set, without any imputation, for our main analysis. For the primary outcome, mean differences in MADRS scores observed (point estimates and confidence intervals was compared with 0 (absence of difference) and 6.5 points (the threshold defined a priori in initiation studies [6]). When IPD were not available we used aggregated data insofar as possible. If applicable (> 10 studies), we planned to use a funnel plot to look for any small study effects. Two reviewers (CG and FN) independently assessed the overall certainty of evidence using GRADE [32].

### Moderators of response to esketamine

In order to study the impact of resistance stage and age, a one-step approach was used: each participant had to be classified according to Thase and Rush resistance stages (see **e-Box 1** in **Web appendix 1**) [26] in accordance with their baseline characteristics. This classification, as well as the age of the patient, were explored as treatment effect modifiers in a one-stage IPD meta-analysis using a mixed model [33].

Another sensitivity analysis was performed to explore if efficacy varies according to the World Bank categorization into low, middle, and high income (https://data.worldbank.org/country) as initial evidence on antidepressants suggests that per capita gross national income is associated with trial results [34].

### Anticipated number of patients

**e-Table 2** (in **Web appendix 1**) presents studies available on the YODA repository when we submitted this registered report. A minimum of 721 patients were expected in the pivotal initiation trials and 719 patients (of whom 455 entered in optimization phase and 297 were randomised) were expected in the continuation trial. These numbers were expected to increase by additional studies available on YODA (with a maximum of 834 depending on the inclusion criteria of participants) and/or identified during the systematic review process.

### Changes to the initial protocol

#### PICOS

Although esketamine was approved for use in combination with a novel antidepressant in TRD, we included a monotherapy study (NCT04599855 [35]) in our analysis. This study wasn’t pooled with combination studies as it addresses a different research question.

#### Re-analyses

The information about study centers was not provided due to data anonymization requirements. As leave-one-out analysis by centers was not possible, we instead used an exploratory approach excluding countries. This method was applied solely to the continuation trial [12] that prompted this analysis, as highlighted by the FDA.

Our re-analysis of NCT02422186 [10] revealed differing outcomes before and after the interim analysis. Consequently, we conducted an exploratory analysis to investigate this discrepancy.

#### Data-management

Upon reviewing the data structure, prior to any analysis, we refined the definitions of dissociation, sedation, and suicidal ideation to align with the binary outcome definitions specified in the protocol. We defined CADSS values over 4 as a dissociated state [36, 37]. For sedation, we categorized it as “no sedation” versus any other type, using the MOAA-S scale. For both scales, a complementary analysis was conducted using them as continuous outcomes. Regarding suicidal ideations, we initialy planned to analyse it as a continuous outcome. Because the intended scale was unavailable and some data were reported as binary, we analysed it as a binary outcome. We used the Columbia-Suicide Severity Rating Scale (C-SSRS) part on Suicidal ideation, along with adverse events reported as “Suicide ideation,” “Passive suicidal ideation,” “Suicidal intention,” and “Depression suicidal” to identify suicidal ideations. More precisely, suicidal ideation in C-SSRS was defined if the sum of 5 items exceeded 3 (questions 3, 4, and 5 were answered only if question 2 was “yes”). To identify suicide attempts or suicides, the “Actual Attempt” and “Suicide” items of the Columbia-Suicide Severity Rating Scale (C-SSRS) part Suicidal behavior were selected, along with adverse events reported as “Suicide attempt”, “Suicide”, “Attempted suicide”, “Suicidal behavior”, “Completed suicide”.

Regarding the “stage of resistance”, it was not feasible to adhere strictly to the Thase and Rush classification for our moderator analysis as initially planned. First, information on ECT - needed to define stage 5-was inconsistent across studies. Second, many patients were prescribed various antidepressants without following the specific sequence required by this classification. Consequently, we modified the Thase and Rush classification, interpreting it based on the number of classes tested irrespective of the sequence (i.e., one class = stage I, two classes = stage II, three classes = stage III, four or more classes = stage IV). We performed a sentitivity analysis excluding all patients that received Monoamine oxidase inhibitors (MOAIs). This approach was further validated through discussions with Thase and Rush, who concurred with the adaptation (personal communication, July 12, 2024).

#### Meta-analysis

For the two-stage meta-analysis, we indicated in the SAP that the confidence interval would be calculated using the Hartung-Knapp Sidik-Jonkman method, as recommended [38]. Sensitivity analyses based on multiple imputation were performed to explore robustness of our findings. The Peto method was used to take account of rare events.

## RESULTS

### Study selection and IPD obtained

Literature searches were conducted on 14/01/2022 (bibliographic databases), yielding a total of 975 citations after duplicate removal. Of these, 808 records were excluded based on their titles and abstracts for not meeting the selection criteria. Following a full-text review of the remaining 167 articles, an additional 125 references were excluded. We also identified 56 potentially relevant reports based on searches of the manufacturer’s website, the YODA website, health authority websites, and an update prior to publication informed by 1/ an ongoing Cochrane review on the same topic [27], which identified 19 studies (of which several were ineligible) and 2/ another recent meta-analysis on aggregated data [28], which identified 10 additional potentially eligible studies. Ultimately, after deduplication 60 reports on 8 studies were included in the quantitative review, with 7 incorporated into the main analysis based on individual participant data availability. A flowchart illustrating the study selection process is presented in **Figure 1**.

**Figure 1:**
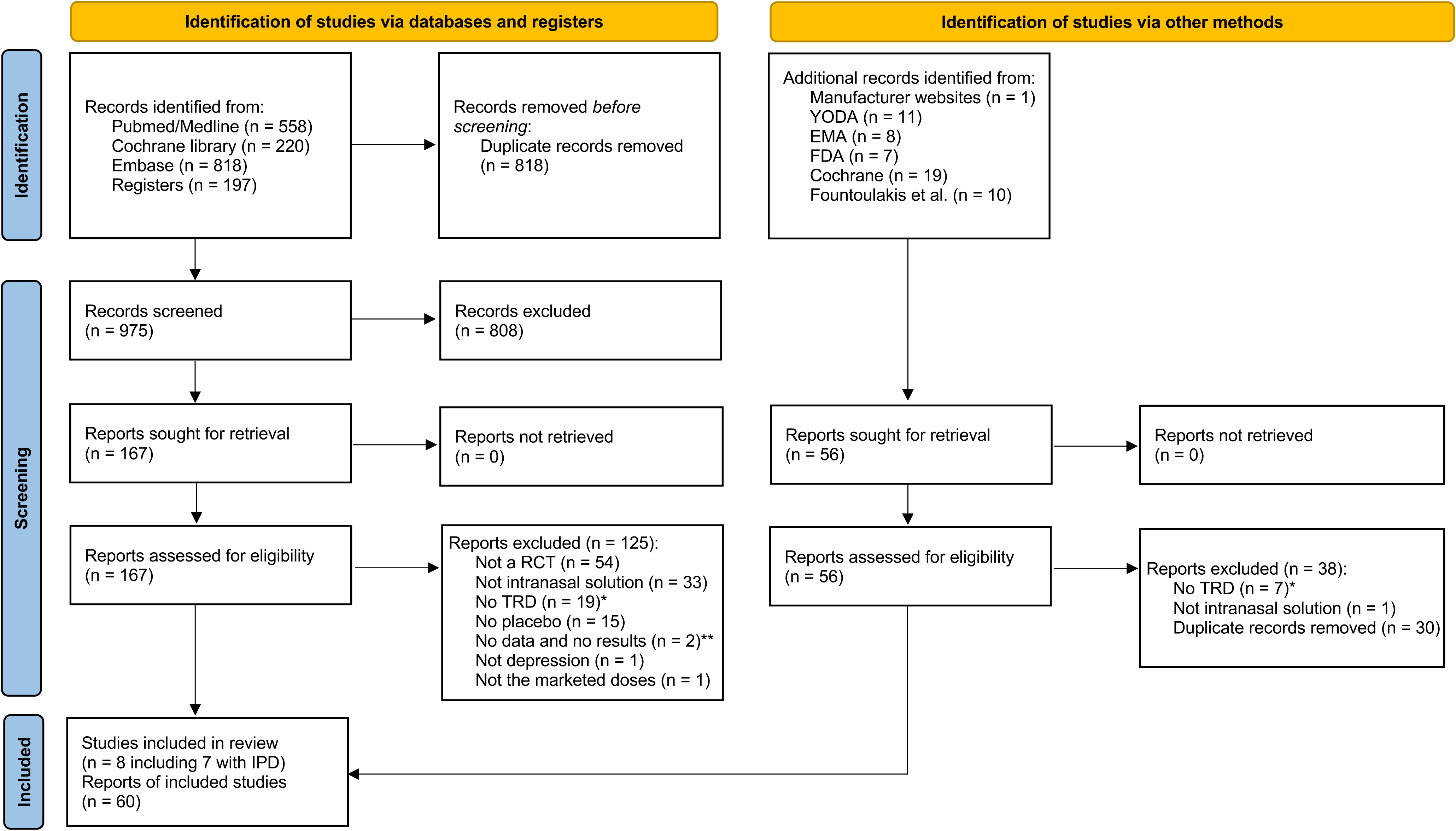
Flow diagram. * NCT02133001, NCT03039192 and NCT03097133 were not limited to treatment-resistant depression (TRD) and provided insufficient information regarding prior resistance in the individual patient data (IPD); NCT03185819 and NCT02919579 were not limited to treatment-resistant depression (TRD) and did not provided IPD. ** NCT03852160 has stopped before enrolling its first participant; ChiCTR2000038915 has not responded to our request to share data.

### Study characteristics

We identified 1505 randomized participants across 7 RCTs exploring the efficacy of esketamine in combination with antidepressants. Among these, 6 were initiation studies (1 with a 1-week duration [39], 5 with a 4-week duration [9-11, 40, 41] and one was a continuation study [12]. One monotherapy RCT (NCT04599855 [35], 476 participants) was analyzed separately because it addressed a different question and its data were not yet available despite a request to YODA. All studies focused on adults with TRD. The key characteristics of the included studies are summarized in **Table 1**.

### IPD integrity and re-analyses

Detailed results of the re-analyses are available on OSF (https://osf.io/z2bna/) and summarized in **Figure 2**. **Table 1** provides an overview of all protocol deviations identified using IPD. For all studies, after an initial re-analysis based solely on the study protocol, the statistician (ELP) had to refer to each study’s statistical analysis plan to format the data in a way comparable to the published report (e.g., using least square (LS) means instead of observed means), enabling the assessment of inferential reproducibility. Then, the study conclusions were fully reproduced for 5 out of 7 studies. Among the three pivotal trials, one (NCT02418585 [11]) demonstrated positive results on its primary endpoint, while two (NCT02417064 [9] and NCT02422186 [10]) did not meet their primary endpoints. In one of those (NCT02417064 [9]), statistical significance should not have been tested for the 56 mg treatment group due to the hierarchical analysis and the absence of a significant difference in the 84 mg group tested first. Because this was statistically tested when it should not have been, it introduced some spin into the published report [9], despite the numerical results being deemed reproducible. Additionally, while we were able to reproduce NCT02422186 [10], we observed a drop in placebo response between week 3 and week 4 after an interim analysis, but not before the interim analysis. However, when tested, this apparent difference did not show statistical significance (p = 0.25 for an interaction effect, **Figure 3**). The FDA statistical reviewer observed a similar pattern before and after the interim analysis of the 84 mg dose arm in NCT02417064, noting “no obvious reasons for the difference between stage 1 LS mean difference (−0.29) and stage 2 LS mean difference (−5.68)”. Of the 3 remaining initiation trials, we confirmed the negative conclusions of NCT02918318 [40] and NCT03434041 [41]. We verified the positive efficacy at one week in NCT01998958 [39] but could not compare 4 out of 10 analyses due to unpublished results. In the continuation trial (NCT02493868 [12]), the primary outcome of remission remained consistent except when data from all Polish centers were excluded; in this case, the effect size decreased and the analysis was not statistically significant. A comparable decrease in effect size occurred for the secondary outcome of response and overall (**Figure 4**); however, statistical significance was maintained.

**Figure 2:**
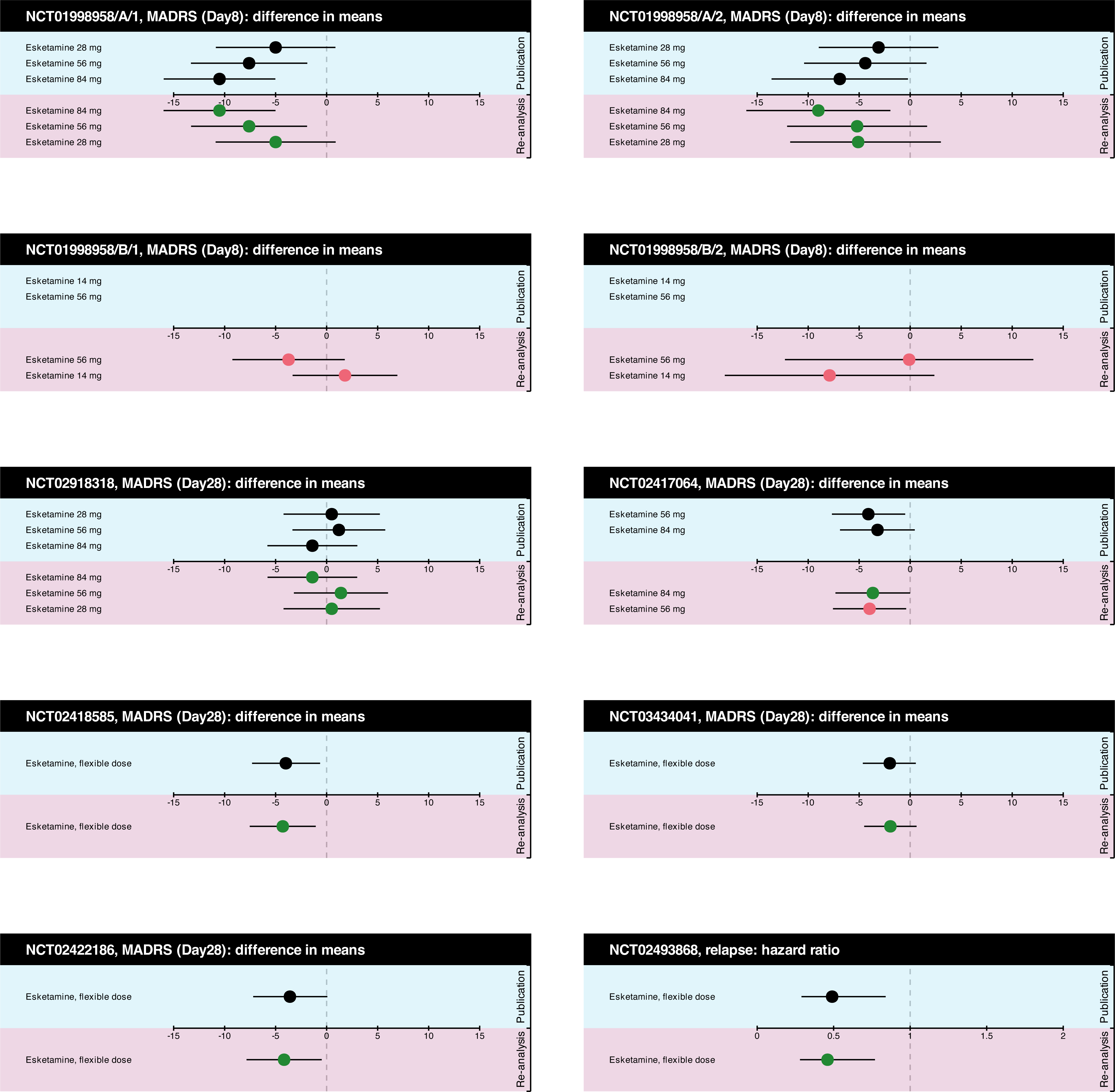
Results of re-analyses compared to those reported in published papers. Point estimates in green represent studies where the main results were considered to achieve inferential reproducibility. In contrast, point estimates in red indicate analyses where inferential reproducibility was not achieved, either due to reporting bias in NCT01998958 or failure to adhere to the hierarchical analysis in NCT02417064

**Figure 3:**
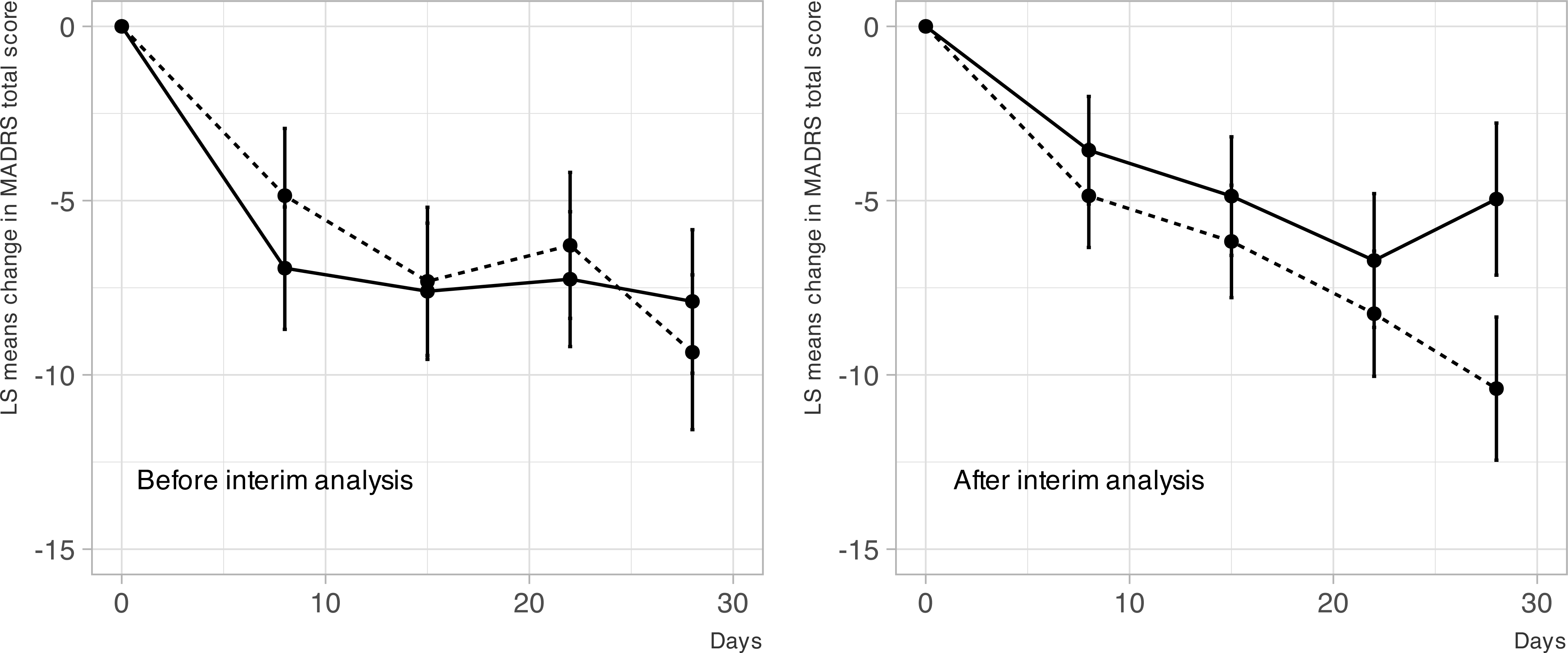
Hierarchical analysis details in NCT02417064 - Data integrity checks.

**Figure 4:**
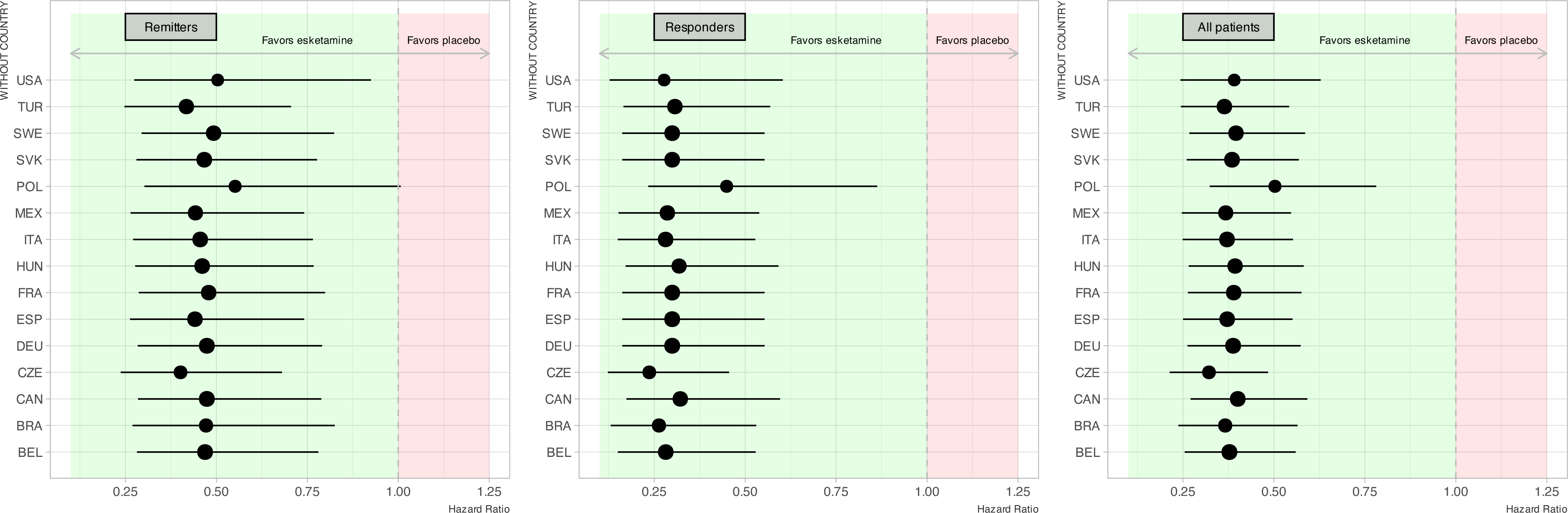
Leave-one-out analysis of NCT02493868 at the country level - Data integrity checks.

Results of our re-analyses were consistent with FDA reports and European public assessment reports (**e-Table 3** in **Web Appendix 1**).

### Results of individual studies and results of syntheses

#### Efficacy: initiation trials in combination with antidepressants

In terms of statistical significance, superiority of esketamine, compared to placebo, was evidenced for the primary outcome (**Figure 5**), specifically the MADRS score assessed after at least 4 weeks in 5 initiation trials (Mean Difference, MD = −2.94 points, 95% CI [−5.39; −0.48], random effects model, tau^2^ = 1.15). However, in terms of clinical significance, the entire 95% confidence interval failed to meet the prespecified threshold of −6.5 points. **Figure 5** presents the results and the risk of bias assessment within the studies. Similar results were observed in the meta-analysis using multiple imputation, although the magnitude of the drug-placebo difference was smaller (MD = −2.47 points, 95% CI [−3.81; −1.13], fixed effects model, tau^2^ = 0.47). Similar effects were noted for the MADRS score assessed at 1 week in 6 initiation trials (MD = −2.51 points, 95% CI [−4.81; −0.20], random effects model, tau^2^ = 2.06), where the entire 95% confidence interval also failed to meet the threshold.

**Figure 5:**
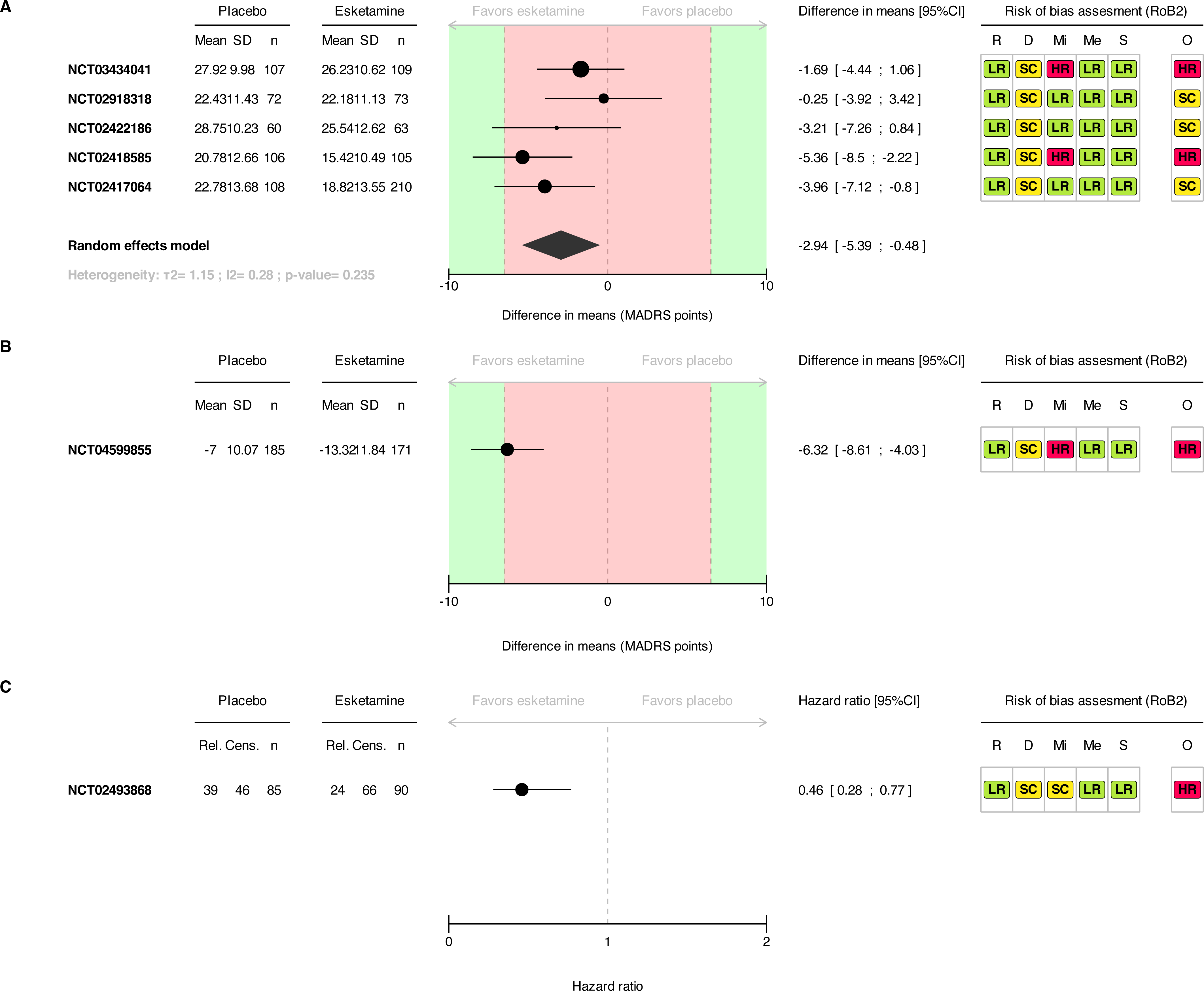
Forest plot and risk of bias (RoB2) assessment for MADRS at 4 weeks in initiation trials (in combination A, and monotherapy B) and relapse in continuation trial (combination study C) Pannel A and C are based on IPD. Pannel B is based on aggregated data. Dashed lines represent the lower and upper prespecified thresholds (± 6.5 points) and the null effect (0). Green-shaded areas denote effects exceeding the threshold, while the red-shaded area indicates effects below the threshold. CI: Confidence interval SD: Standard deviation Rel: Relapsed Cens: Censored R: Bias arising from the randomisation process D: Bias due to deviations from intended interventions Mi: Bias due to missing outcome data Me: Bias in measurement of the outcome S: Bias in selection of the reported results O: Overall risk of bias

**Table 2** shows the results for secondary outcomes from two-stage meta-analyses. Blood pressure was analyzed with a one-stage individual participant data meta-analysis, revealing increases in LS means of 8.96 [7.55; 10.37], 6.09 [4.68; 7.50], and 3.58 [2.17; 4.99] mmHg at 40 minutes, 1 hour, and 1 hour 30 minutes respectively for systolic blood pressure. Similar trends were observed for diastolic blood pressure.

**Table 2:** Summary of findings from secondary outcomes. MD = Mean differences, RR = Risk Ratio, OR = Odds Ratio, IRR = Incidence Rate Ratio C: Common effect model R: Random effect model P: Peto method ** 176 in a stable remission and 121 in a stable response

|  | Meta-analyses of initiation trials |  |  |  |  | Continuation trial |  |  |
| --- | --- | --- | --- | --- | --- | --- | --- | --- |
| Outcomes | Effect size (type) | Number of studies | Number of participants | Effect size (95%CI) | Tau2 | Effect size (type) | Number of participants | Effect size (95% CI) |
| **Depression severity** |  |  |  |  |  |  |  |  |
| MADRS (1 week) | MD | 6 | 1173 | -2.51 (-4.81; -0.20)<sup>R</sup> | 2.06 | . | . | . |
| Remission (binary outcome) | RR | 5 | 1013 | 1.39 (0.95; 2.04)<sup>R</sup> | 0.02 | . | . | . |
| PHQ-9 (continuous outcome) | MD | 3 | 640 | -2.73 (-3.89; -1.57)<sup>C</sup> | 0.00 | . | . | . |
| Relapse (censored outcome) for continuation trials | . | . | . | . | . | HR | 297** | 0.38 (0.26; 0.57) |
| **Quality of life** |  |  |  |  |  |  |  |  |
| Sheehan Disability Scale | MD | 5 | 1008 | -1.85 (-3.86; 0.17)<sup>R</sup> | 1.28 | . | . | . |
| **Suicidality** |  |  |  |  |  |  |  |  |
| Suicide/suicide attempt | OR | 5 | 1123 | 7.51 (0.78; 72.41)<sup>C,P</sup> | 0.00 | OR | 297 | 4.83 (0.39; 668.94) |
| Suicidal ideations* | OR | 5 | 1123 | 2.09 (0.79; 5.53)<sup>C,P</sup> | 0.00 | OR | 297 | 0.95 (0.04; 24.27) |
| **Safety outcomes** |  |  |  |  |  |  |  |  |
| Serious adverse events | IRR | 5 | 1123 | 1.35 (0.54; 3.40)<sup>C</sup> | 0.00 | IRR | 297 | 3.34 (0.42; 26.21) |
| Adverse events | IRR | 5 | 1123 | 3.91 (2.37; 6.45)<sup>R</sup> | 0.15 | IRR | 297 | 5.45 (4.20; 7.06) |
| Dropout for any cause | RR | 5 | 1123 | 1.15 (0.86; 1.54)<sup>C</sup> | 0.00 | IRR | 297 | 1.04 (0.47; 2.28) |
| Dropout due to adverse events | RR | 5 | 1123 | 2.66 (1.32; 5.35)<sup>C</sup> | 0.00 | IRR | 297 | 0.48 (0.04; 5.20) |
| Dissociation | RR | 5 | 1122 | 2.36 (2.10; 2.65)<sup>C</sup> | 0.00 | RR | 297 | 4.14 (2.92; 5.87) |
| Sedation | RR | 4 | 873 | 3.70 (2.02; 6.78)<sup>R</sup> | 0.07 | RR | 297 | 2.86 (1.87; 4.38) |

Detailed results, meta-analyses, and heterogeneity assessments for all outcomes (including last follow-up visit post-treatment) are available in the statistical report (**Web appendix 3**). We considered including studies without IPD for sensitivity analysis, but this was unnecessary as the only such study -a monotherapy trial described below-could not be pooled due to differences in PICOS.

#### Efficacy: initiation trial in monotherapy

Based on the aggregated data in one monotherapy trial, the statistical superiority of esketamine was evidenced for the primary outcome (**Figure 5**), specifically the MADRS score assessed after at 4 weeks in the unique available initiation trial in monotherapy (MD = − 6.32 points, 95% CI [−8.62; −4.03]). The 95% confidence interval did overlap the prespecified clinical significance threshold of −6.5 points. However, as we had no access to the IPD, uncertainty remain regarding risk of bias (**Figure 5**). Moreover, participants who improved during a 2-week or longer antidepressant-free lead-in were excluded from the primary efficacy analysis, possibly resulting in an enriched sample that could bias efficacy estimates. On day 28, around 75% of the esketamine group and 50% of the control group correctly guessed their group allocation with certainty, suggesting functional unblinding.

#### Efficacy: continuation study

The superiority of esketamine in reducing relapse was overall demonstrated in the study NCT02493868 (Hazard ratio, HR= 0.38 [0.26; 0.57]), among both remitters (hazard ratio, HR= 0.46 [0.28; 0.77]) and responders (hazard ratio, HR= 0.30 [0.16; 0.55]). These efficacy estimates are based on data from all countries, including Poland. **Figure 4** presents the results for both remitters (the main outcome) and responders, along with the risk assessment of bias. Additional outcomes are in **Table 2**.

#### Safety

Esketamine showed no increase in serious adverse events or dropout for any cause. The details of all serious adverse events is provided in the statistical report (**Web appendix 3**). Several suicidal behaviors were reported in patients treated with esketamine across different studies. These include a completed suicide (day 4) and a suicide attempt (day 2) in study NCT03434041, as well as suicidal and self-injurious behavior (day 14) in study NCT02422186. An additional death due to a road accident occurred on study day 16 in an esketamine-treated patient from study NCT02418585. There was an increase in adverse events, dropout due to adverse events, dissociation, and sedation (see **Table 2** for effect estimates and Web appendix 3 for absolute numbers). Our complementary analyses on dissociation and sedation show a mean CADSS difference of MD = 9.68 [7.49; 11.87] points (random effects model, tau2 = 1.75). See **Figure 6** for detailed MOAAS results.

**Figure 6:**
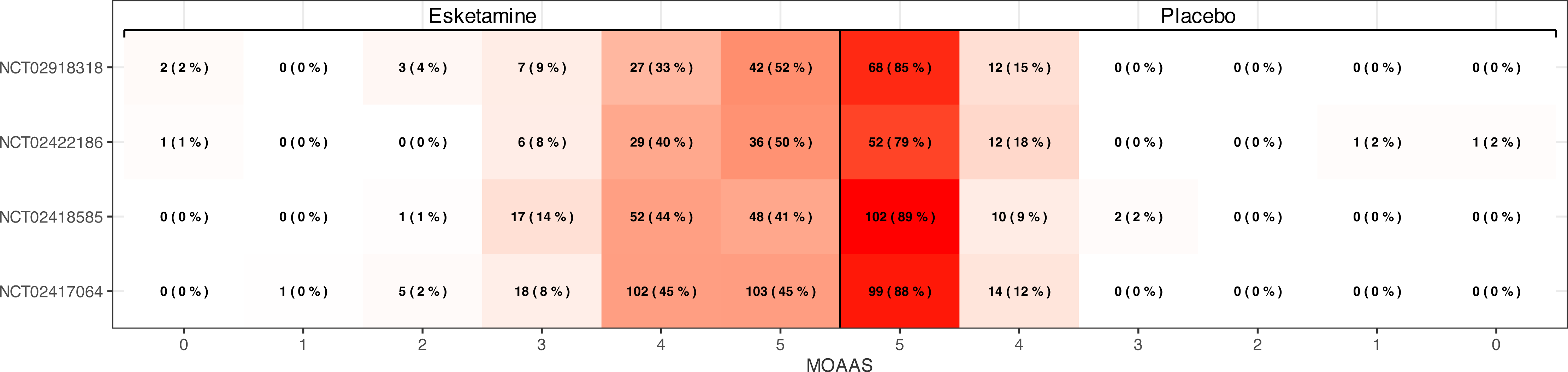
Distribution of Modified Observer Assessment of Alertness and Sedation (MOAAS) scores across trials. The categories of the MOAAS scale are: 5: Responds readily to name spoken in normal tone 4: Lethargic response to name spoken in normal tone 3: Responds only after name is called loudly and/or repeatedly 2: Responds only after mild prodding or shaking 1: Responds only after painful trapezius squeeze 0: Does not respond to painful trapezius squeeze

### Reporting bias

There were too few studies to generate a funnel plot [42]. In total, we identified one potentially relevant study without IPD and no aggregated data for analysis. For the included studies with available data, we had full access to IPD, ensuring a low risk of reporting bias. However, certain variables, such as center information, were unavailable due to the anonymization process.

### Additional analyses

**Figure 7** presents the exploration of moderators of response to esketamine using the one-step approach. For the resistance stage, the analysis included 334 patients who had received 1 class of antidepressants prior to randomisation, 648 patients who had received 2 classes of antidepressants prior to randomisation, 29 patients who had received 3 classes of antidepressants prior to randomisation, and 2 patients who had received 4 classes of antidepressants prior to randomisation. As only one study provided details on prior use of ECT we could not use this information. No significant differences in efficacy were observed across these groups (p-value = 0.80). Excluding MAOIs, the analysis confirmed the result’s robustness. The analysis found no significant difference in efficacy across low-, middle-, and high-income countries (p = 0.31). Age did not moderate efficacy outcomes either (p = 0.45).

**Figure 7:**
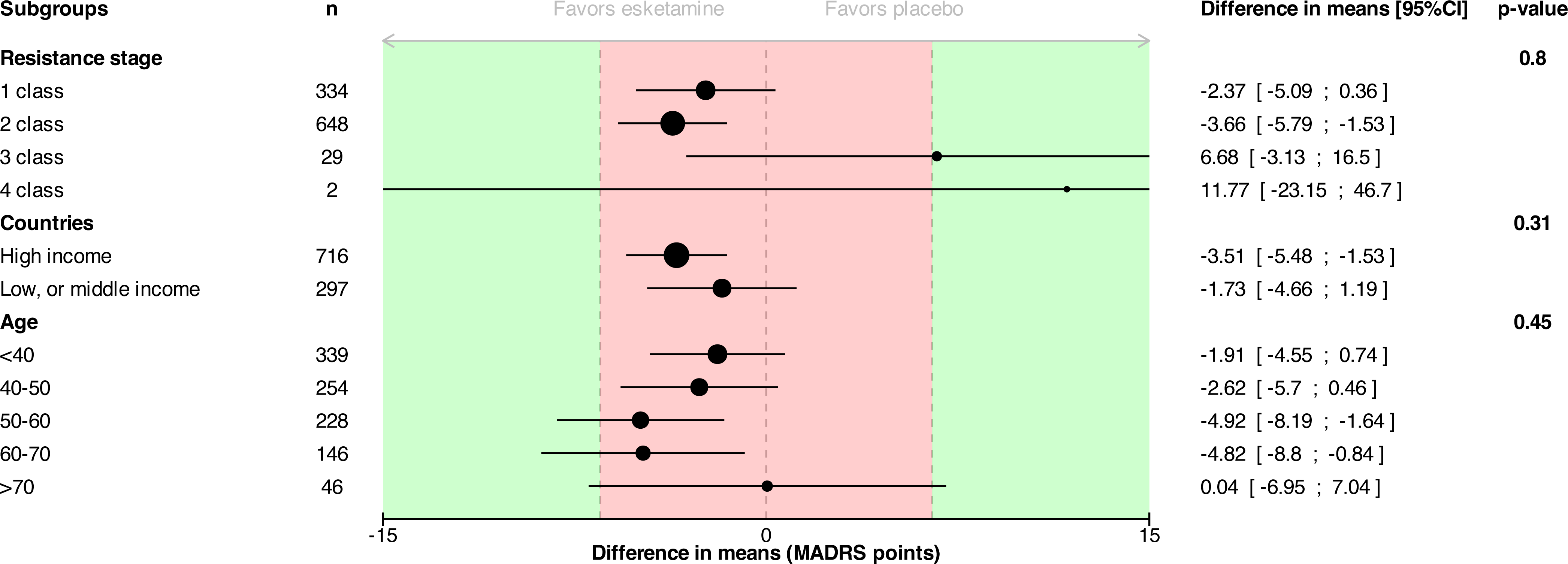
Subgroup analyses exploring moderators of esketamine efficacy. For age, differences in means are presented by age class for visualization purposes. However, the analysis corresponds to a test where age is used as a continuous variable, as pre-specified.

### Certainty of evidence

According to the GRADE approach, our main IPD meta-analysis yields moderate-certainty evidence, downgraded due to the overall high risk of bias in 2 of the 5 included studies.

## DISCUSSION

### Summary of Evidence

This individual patient data meta-analysis offers a comprehensive re-evaluation of clinical trials that assess the efficacy of esketamine in treatment-resistant depression when used alongside antidepressants. This assessment is conducted in accordance with the criteria set forth by the FDA marketing approval process. While esketamine demonstrated superiority in one pivotal initiation trials and in a phase 2 initiation trial, four other initiation trials (including 2 pivotal trials) failed to demonstrated efficacy. Nevertheless, the meta-analysis of initiation trials confirmed a statistically significant superiority of esketamine, with an effect size of approximately three points on the Montgomery-Åsberg Depression Rating Scale. The clinical relevance of this effect size warrants consideration. First, the observed difference raises concerns about its meaningfulness, in the context of TRD, a chronic condition where short-term trials (four weeks) may not adequately capture long-term outcomes [5]. Additionally, there is uncertainty regarding what constitutes a clinically relevant difference on the MADRS scale, with estimates ranging between 3 and 9 points [43]. Furthermore, the - arguably small-identified difference in means may be overestimated due to attrition bias, as suggested by the lower estimates in multiple imputation analyses.

In addition, regardless of the statistical approach, the observed difference falls below the predefined threshold used in the initiation trials. This may explain the inconsistent findings across individual trials, which were likely underpowered to detect esketamine’s effects reliably. In this registered report, we indeed set an *a priori* threshold of 6.5 MADRS points, based on the pivotal trials’ design. Johnson & Johnson, who reviewed our application at the Yale Open Data Access Project emphasized that this threshold was not intended to define clinical relevance. However, the fact that this threshold was used for sample size calculation, based on phase 2 results and “clinical judgement” (as stated in the protocols of the three initial pivotal trials [9–11]), is an acknowledgement that this was the minimal effect that the sponsor would consider clinically meaningful. Furthermore, the superiority observed on depression scales did not translate into a demonstrated improvement in quality of life. Additionally, the 3-point difference on the MARDS identified in our study corresponds with the efficacy range observed for many antidepressants and clinical significance of esketamine for TRD was previously contested, as the standardized mean difference from the three initiation pivotal trials was similar to that of other previously approved drugs for this condition [7]. Given the cost and safety concerns associated with esketamine, one would reasonably expect a greater benefit than that seen with existing antidepressants. For example, regarding economic evaluation, a report from the Canadian Agency for Drugs and Technologies in Health (CADTH) concluded that for a population such as the one in the pivotal trials, esketamine plus oral antidepressant would not be cost-effective at a willingness to pay threshold of $50000 per Quality-Adjusted Life Year (QALY), and a price reduction of 60% would be required for the combination to be considered cost-effective [44]. The monotherapy trial [35] with aggregated data indicates a stronger effect, but excluding participants who improved during the lead-in and possible functional unblinding may have inflated efficacy estimates.

While no increase in serious adverse events (SAEs) was observed in the trials, there was a clear increase in adverse events. Large effect sizes were reported for sedation and dissociation, alongside transient increases in blood pressure. Notably, esketamine did not demonstrate a reduction in suicidal behavior or ideation, despite its approval for depressed patients with suicidal ideation in the United States [45]. Of note, esketamine showed greater efficacy in the continuation trial, with an even larger effect size than in the initiation trials. Such finding is expected, as continuation trials are known to overestimate effect sizes due to potential withdrawal symptoms and the selection of a highly responsive subgroup of patients [46].

No association was found between per capita gross national income and the outcomes of esketamine trials. No significant moderators of efficacy were identified, including age, or stage of treatment resistance. Interestingly, most patients included in the trials were at a low stage of resistance (failure of one or two antidepressant classes). While this limits the power of subgroup analyses, it also informs the actual target population for esketamine. In this relatively less treatment-resistant population, alternative treatments with comparable efficacy exist, including other antidepressants and psychotherapies, which are not encompassed by the definition of treatment resistance.

### Strengths and Limitations

This study presents several strengths. The availability of IPD from the esketamine development program allowed for an independent in-depth examination of each study. The pooled estimates and conclusions matched previous meta-analyses [4, 28] and may offer more clarity on evidence certainty due to the IPD approach. Additionally, the IPD approach enabled the analysis of efficacy moderators, which would not have been possible with aggregated data.

However, some limitations remain. In the continuation trial, we were unable to investigate concerns raised by the U.S. FDA regarding a specific center in Poland, as this information was not available in the IPD dataset. While withholding this information may be justified for privacy, it prevents thorough re-analysis of the issue raised by the FDA. Although our country-based analysis corroborated these concerns, it likely remains insufficient and only provide weak and exploratory evidence on a potentially serious concern. Two initiation trials showed an unexpected data pattern before and after interim analysis, indicating potential bias. While this could be due to chance, it raises concerns about possible bias in those two studies.

More generally, bias is likely in these trials due to functional unblinding. Despite efforts to mitigate measurement bias through blinded evaluators, no formal assessment of unblinding was conducted in pivotal RCTs, leaving room for potential performance bias. However, this was investigated in the monotherapy randomized controlled trial [35], which indicated evidence of functional unblinding.

Additionally, some relevant patient characteristics, such as prior electroconvulsive therapy (ECT) use, were not systematically assessed, limiting our ability to characterize the target population more precisely. Moreover, reconstructing several variables from multiple assessed outcomes introduced methodological variability, potentially explaining discrepancies between our findings and previous meta-analyses. Reconstructing the resistance stage from trial data was challenging, requiring several decisions before analysis. For example, compared to the EMA documents, we identified fewer patients in the three-class antidepressant stage, though this data confirms that few patients had high resistance before esketamine [2].

Similarly, a previous meta-analysis reported esketamine caused dissociation seven times more frequently [4]. Our study found a smaller, but still large effect size. Interpreting differences in CADSS scores is likely to provide more informative insights than dichotomising outcomes. This approach emphasizes the large impact of esketamine on dissociation.

### Implications

These findings have important clinical implications. This meta-analysis resolves some uncertainties that persisted following its approval, though several issues still remain. Importantly, the patient population included in the esketamine trials does not represent a highly treatment-resistant group. In this population, alternative treatments with similar, thought still small effect sizes exist, sometimes supported by more extensive or comparative evidence. For example, in a network meta-analysis, intranasal esketamine was not superior to augmentation with either aripiprazole or lithium for response rates in patients with no response to one or more antidepressants, a somewhat broader population [47]. Regarding the efficacy of esketamine, it is important to consider our findings in the context of the open-label ESCAPE-TRD trial, which evaluated flexible dosing of esketamine nasal spray against extended-release quetiapine, both administered alongside an SSRI or SNRI in patients with treatment-resistant depression. The ESCAPE-TRD trial reported a comparatively smaller effect size for esketamine versus quetiapine (−2.2 MADRS points; 95% CI: −3.6 to −0.8) at 32 weeks [19]. The evidence regarding quetiapine’s efficacy in TRD varies among studies: earlier meta-analyses reported limited efficacy and noted safety concerns compared to placebo [48], whereas a recent open-label randomized controlled trial indicated potential benefits versus lithium [49]. Moreover, the approval of quetiapine for TRD was based on an even more lenient definition of resistance, requiring failure of only one prior antidepressant [48]. Whether quetiapine would have outperformed placebo in ESCAPE-TRD is currently unknown. Taken together, these results suggest that esketamine’s effects are small. Clinicians must weigh esketamine’s efficacy against its cost and safety profile when considering it as a treatment option. Possible safety issues have indeed been described in post-marketing studies [16, 50].

Regarding treatment personalization, we have not identified any patient subgroups which would derive enhanced benefits from esketamine, though the number of patients who had not responded to three or more antidepressant treatments was limited. These findings suggest that there is no evidence to support that esketamine should be primarily prescribed to the more severe or treatment-resistant patients, further constraining the clinical utility of this drug.

Beyond clinical practice, these findings are relevant to regulatory agencies, including the EMA and FDA. The case of esketamine in TRD illustrates how accelerated approval pathways, such as the breakthrough therapy designation, can facilitate the approval of drugs with limited evidence. Esketamine’s approval is not an isolated case, as several psychedelic compounds have now been granted similar regulatory designations [51]. This approach promotes the generation of short-term evidence against placebo, although the priority should be given to drugs that demonstrate long-term effectiveness against active comparators.

Finally, this IPD meta-analysis underscores the value of data sharing in ensuring research integrity. Re-analysis of clinical trial data enables thorough scrutiny of drug efficacy and safety, reinforcing the robustness of evidence-based medicine. However, our study also exemplifies limitations, such as the considerable time investment required for IPD analyses and challenges related to data anonymization, which can restrict access to key variables (e.g., study centers). Despite these constraints, IPD meta-analyses remain essential for strengthening our evidence synthesis framework, enhancing the reliability of clinical research and providing unique opportunities to identify patient-level predictors of treatment response, thereby supporting the development of more personalised and effective interventions.

## Supporting information

Web appendix 1

Web appendix 2

Web appendix 3

## Data Availability

The datasets that were analysed during the current study are available in the YODA repository (https://yoda.yale.edu/). The detailed SAP (https://osf.io/hmcas), the code to reproduce our analyses (https://osf.io/z2bna/files/osfstorage) are shared on the OSF.

## List of abbreviations

ANSM: Agence Nationale de Sécurité du Médicament et des produits de santé
BSS: Beck Scale for Suicidal Ideation
CHMP: Committee for Medicinal Products for Human Use
EMA: European Medicines Agency
FDA: Food and Drug Administration
HAS: Haute Autorité de santé
IPD: Individual Participant Data
MADRS: Montgomery-Åsberg Depression Rating Scale
NICE: National Institute for Health and Care Excellence
PHQ-9: Patient Health Questionnaire-9
RCT: Randomised Controlled Trial
RoB 2: Version 2 of the Cochrane risk-of-bias tool for randomized trials
SSRI: Selective serotonin reuptake inhibitors
SNRI: Serotonin and norepinephrine reuptake inhibitors
TRD: Treatment Resistant Depression

## DECLARATIONS

### Ethics approval and consent to participate

Not applicable

### Consent for publication

Not applicable

### Availability of data and materials

The datasets that were analysed during the current study are available in the YODA repository (https://yoda.yale.edu/). The detailed SAP (https://osf.io/hmcas), the code to reproduce our analyses (https://osf.io/z2bna/files/osfstorage) are shared on the OSF. Prior to the submission of the final manuscript, the results were shared on the MedRxiv preprint server [ADD HERE THE LINK].

### Authors’ contributions

FN, ET, and IC wrote the first draft of the protocol. FN secured funding. CP, LF, CG, CB contributed to the writing of the protocol. FN, CP, LF, and IC performed literature search and data extraction. CP and FN wrote the first draft of the SAP. ELP and FN analysed the data. FN, ELP, CP, LF, CG, CB, ET and IC interpreted the results. FN and IC wrote the first draft of the manuscript. CP, LF, CG, CB, ET, ELP contributed to the writing of the manuscript. All authors have read, and confirm that they meet, ICMJE criteria for authorship.

## Acknowledgements

This study, carried out under YODA Project #2021-4851 used data obtained from the Yale University Open Data Access Project, which has an agreement with JANSSEN RESEARCH & DEVELOPMENT, L.L.C.. The interpretation and reporting of research using this data are solely the responsibility of the authors and does not necessarily represent the official views of the Yale University Open Data Access Project or JANSSEN RESEARCH & DEVELOPMENT, L.L.C.. The original proposal can be found: https://yoda.yale.edu/data-request/2021-4851/.

## Authors’ information

Florian Naudet has developed a group dedicated to reanalysis of clinical trial data. His research interests are evaluating and developing methodological solutions to assess treatments in patients primarily, but not exclusively, in psychiatric research. He has a strong interest in studying research waste and data sharing practices. He has worked in the fields of clinical pharmacology, research methodology, epidemiology, and neurosciences.

Claude Pellen is a pharmacist and statistician. He was a PhD student at Rennes University during this study and has since completed his PhD. He has a strong interest in meta-research and open science practices. His research focuses in maximising the impact of clinical trials data sharing.

Liviu-Andrei Fodor is a research assistant at the International Institute for the Advanced Studies of Psychotherapy and Applied Mental Health and a doctoral student at the “Psychodiagnostics and scientifically validated psychological interventions” Doctoral School. His main research interests include meta-research procedures and advanced research methods for evaluating the efficiency / effectiveness of technologically augmented psychological interventions in the treatment of mental health disorders.

Chiara Gastaldon is a psychiatrist and post-doc research fellow at the WHO Collaborating Centre for Research and Training in Mental Health and Service Evaluation, Department of Neuroscience, Biomedicine and Movement Sciences, Section of Psychiatry and Cochrane Global Mental Health at the University of Verona, Italy. Her main research interest consists in pharmacoepidemiology, oriented to describe the beneficial and harmful effects of psychotropic drugs in ordinary practice. She collaborates with the Cochrane Collaboration, conducting systematic reviews and meta-analysis on efficacy and safety of psychotropic drugs and psychosocial intervention in low and middle income countries.

Corrado Barbui coordinates the activities of the WHO Collaborating Centre for Research and Training in Mental Health and Service Evaluation at the University of Verona, Italy, and conducts research in the field of global mental health. He founded in 2017 Cochrane Global Mental Health, a Cochrane network that supports the production, dissemination and implementation of systematic reviews relevant to mental health in low- and middle-income countries.

Erick H. Turner has expertise in regulatory science. His work is aimed at increasing medical research transparency to make the evidence base more complete, truthful, and reliable. He previously worked as an NIH intramural researcher, then as an FDA reviewer, essentially serving as a gatekeeper for drugs new to the US market.

Estelle Le Pabic is a senior statistician at Rennes University Hospital, with expertise in the analysis of academic randomized controlled trials.

Ioana A. Cristea is a clinical psychologist and Associate Professor of Clinical Psychology at the University of Padova, Italy. Her research focuses on critically appraising the efficacy of various psychological and pharmacological interventions for mental disorders, as well as well the role of financial and non-financial conflicts of interest.

## REFERENCES

1. Breakthrough Therapy [https://www.fda.gov/patients/fast-track-breakthrough-therapy-accelerated-approval-priority-review/breakthrough-therapy]

2. Spravato : EPAR - Public assessment report [https://www.ema.europa.eu/en/documents/assessment-report/spravato-epar-public-assessment-report_en.pdf]

3. Schatzberg AF: A Word to the Wise About Intranasal Esketamine. The American journal of psychiatry 2019, 176(6):422–424.

4. Gastaldon C, Papola D, Ostuzzi G, Barbui C: Esketamine for treatment resistant depression: a trick of smoke and mirrors? Epidemiology and psychiatric sciences 2019, 29:e79.

5. Cristea IA, Naudet F: US Food and Drug Administration approval of esketamine and brexanolone. The lancet Psychiatry 2019, 6(12):975–977.

6. Naudet F, Cristea IA: Approval of esketamine for treatment-resistant depression - Authors’ reply. The lancet Psychiatry 2020, 7(3):235–236.

7. Turner EH: Esketamine for treatment-resistant depression: seven concerns about efficacy and FDA approval. The lancet Psychiatry 2019, 6(12):977–979.

8. Turner EH: Approval of esketamine for treatment-resistant depression - Author’s reply. The lancet Psychiatry 2020, 7(3):236.

9. Fedgchin M, Trivedi M, Daly EJ, Melkote R, Lane R, Lim P, Vitagliano D, Blier P, Fava M, Liebowitz M et al: Efficacy and Safety of Fixed-Dose Esketamine Nasal Spray Combined With a New Oral Antidepressant in Treatment-Resistant Depression: Results of a Randomized, Double-Blind, Active-Controlled Study (TRANSFORM-1). The international journal of neuropsychopharmacology 2019, 22(10):616–630.

10. Ochs-Ross R, Daly EJ, Zhang Y, Lane R, Lim P, Morrison RL, Hough D, Manji H, Drevets WC, Sanacora G et al: Efficacy and Safety of Esketamine Nasal Spray Plus an Oral Antidepressant in Elderly Patients With Treatment-Resistant Depression-TRANSFORM-3. The American journal of geriatric psychiatry : official journal of the American Association for Geriatric Psychiatry 2020, 28(2):121–141.

11. Popova V, Daly EJ, Trivedi M, Cooper K, Lane R, Lim P, Mazzucco C, Hough D, Thase ME, Shelton RC et al: Efficacy and Safety of Flexibly Dosed Esketamine Nasal Spray Combined With a Newly Initiated Oral Antidepressant in Treatment-Resistant Depression: A Randomized Double-Blind Active-Controlled Study. The American journal of psychiatry 2019, 176(6):428–438.

12. Daly EJ, Trivedi MH, Janik A, Li H, Zhang Y, Li X, Lane R, Lim P, Duca AR, Hough D et al: Efficacy of Esketamine Nasal Spray Plus Oral Antidepressant Treatment for Relapse Prevention in Patients With Treatment-Resistant Depression: A Randomized Clinical Trial. JAMA psychiatry 2019, 76(9):893–903.

13. Briefing Information for the February 12, 2019 Joint Meeting of the Psychopharmacologic Drugs Advisory Committee (PDAC) and the Drug Safety and Risk Management Advisory Committee (DSaRM). [https://www.fda.gov/downloads/AdvisoryCommittees/CommitteesMeetingMaterials/Drugs/PsychopharmacologicDrugsAdvisoryCommittee/UCM630970.pdf]

14. Singh JB, Daly EJ, Mathews M, Fedgchin M, Popova V, Hough D, Drevets WC: Approval of esketamine for treatment-resistant depression. The lancet Psychiatry 2020, 7(3):232–235.

15. Janssen Announces U.S. FDA Approval of SPRAVATO® (esketamine) CIII Nasal Spray to Treat Depressive Symptoms in Adults with Major Depressive Disorder with Acute Suicidal Ideation or Behavior [https://www.janssen.com/janssen-announces-us-fda-approval-spravator-esketamine-ciii-nasal-spray-treat-depressive-symptoms]

16. Gastaldon C, Raschi E, Kane JM, Barbui C, Schoretsanitis G: Post-Marketing Safety Concerns with Esketamine: A Disproportionality Analysis of Spontaneous Reports Submitted to the FDA Adverse Event Reporting System. Psychotherapy and psychosomatics 2021, 90(1):41–48.

17. Mahase E: Esketamine for treatment resistant depression is not recommended by NICE. BMJ (Clinical research ed) 2020, 368:m329.

18. Avis de la commission de la transparence [https://www.has-sante.fr/jcms/p_3192924/en/spravato]

19. Reif A, Bitter I, Buyze J, Cebulla K, Frey R, Fu DJ, Ito T, Kambarov Y, Llorca PM, Oliveira-Maia AJ et al: Esketamine Nasal Spray versus Quetiapine for Treatment-Resistant Depression. The New England journal of medicine 2023, 389(14):1298–1309.

20. Horowitz MA, Plöderl M, Naudet F: Esketamine Nasal Spray versus Quetiapine for Resistant Depression. The New England journal of medicine 2024, 390(1):93.

21. Lan CH, Wei JC: Esketamine Nasal Spray versus Quetiapine for Resistant Depression. The New England journal of medicine 2024, 390(1):93–94.

22. Naudet F, Siebert M, Pellen C, Gaba J, Axfors C, Cristea I, Danchev V, Mansmann U, Ohmann C, Wallach JD et al: Medical journal requirements for clinical trial data sharing: Ripe for improvement. PLoS Med 2021, 18(10):e1003844.

23. Warren E: Strengthening Research through Data Sharing. The New England journal of medicine 2016, 375(5):401–403.

24. Krumholz HM, Waldstreicher J: The Yale Open Data Access (YODA) Project--A Mechanism for Data Sharing. The New England journal of medicine 2016, 375(5):403–405.

25. Page MJ, Moher D, Bossuyt PM, Boutron I, Hoffmann TC, Mulrow CD, Shamseer L, Tetzlaff JM, Akl EA, Brennan SE et al: PRISMA 2020 explanation and elaboration: updated guidance and exemplars for reporting systematic reviews. BMJ (Clinical research ed) 2021, 372:n160.

26. Thase ME, Rush AJ: When at first you don’t succeed: sequential strategies for antidepressant nonresponders. The Journal of clinical psychiatry 1997, 58 Suppl 13:23–29.

27. Gastaldon C, Laurenzi PF, Schoretsanitis G, Papola D, Cristea IA, Naudet F, Ostuzzi G, Barbui C: Esketamine for treatment-resistant depression in adults. Cochrane Database of Systematic Reviews 2022(1).

28. Fountoulakis KN, Saitis A, Schatzberg AF: Esketamine Treatment for Depression in Adults: A PRISMA Systematic Review and Meta-Analysis. The American journal of psychiatry 2025, 182(3):259–275.

29. Sterne JAC, Savović J, Page MJ, Elbers RG, Blencowe NS, Boutron I, Cates CJ, Cheng HY, Corbett MS, Eldridge SM et al: RoB 2: a revised tool for assessing risk of bias in randomised trials. BMJ (Clinical research ed) 2019, 366:l4898.

30. ICH E9 statistical principles for clinical trials [https://www.ema.europa.eu/en/ich-e9-statistical-principles-clinical-trials#current-version-section]

31. Viechtbauer W, Cheung MW: Outlier and influence diagnostics for meta-analysis. Research synthesis methods 2010, 1(2):112–125.

32. Guyatt GH, Oxman AD, Vist GE, Kunz R, Falck-Ytter Y, Alonso-Coello P, Schünemann HJ: GRADE: an emerging consensus on rating quality of evidence and strength of recommendations. 2008, 336(7650):924–926.

33. Kontopantelis E: A comparison of one-stage vs two-stage individual patient data meta-analysis methods: A simulation study. Research synthesis methods 2018, 9(3):417–430.

34. Klein T, Weinmann S, Becker T, Koesters M: Antidepressant treatment effects and country income: meta-regression analysis of individual participant data from duloxetine trials. Acta psychiatrica Scandinavica 2021, 144(3):277–287.

35. Janik A, Qiu X, Lane R, Popova V, Drevets WC, Canuso CM, Macaluso M, Mattingly GW, Shelton RC, Zajecka JM et al: Esketamine Monotherapy in Adults With Treatment-Resistant Depression: A Randomized Clinical Trial. JAMA psychiatry 2025.

36. Bremner J: The clinician administered dissociative states scale (CADSS): Instructions for administration. Emory University 2014.

37. Mello RP, Echegaray MVF, Jesus-Nunes AP, Leal GC, Magnavita GM, Vieira F, Caliman-Fontes AT, Telles M, Guerreiro-Costa LNF, Souza-Marques B et al: Trait dissociation as a predictor of induced dissociation by ketamine or esketamine in treatment-resistant depression: Secondary analysis from a randomized controlled trial. J Psychiatr Res 2021, 138:576–583.

38. Partlett C, Riley RD: Random effects meta-analysis: Coverage performance of 95% confidence and prediction intervals following REML estimation. Stat Med 2017, 36(2):301–317.

39. Daly EJ, Singh JB, Fedgchin M, Cooper K, Lim P, Shelton RC, Thase ME, Winokur A, Van Nueten L, Manji H et al: Efficacy and Safety of Intranasal Esketamine Adjunctive to Oral Antidepressant Therapy in Treatment-Resistant Depression: A Randomized Clinical Trial. JAMA psychiatry 2018, 75(2):139–148.

40. Takahashi N, Yamada A, Shiraishi A, Shimizu H, Goto R, Tominaga Y: Efficacy and safety of fixed doses of intranasal Esketamine as an add-on therapy to Oral antidepressants in Japanese patients with treatment-resistant depression: a phase 2b randomized clinical study. BMC Psychiatry 2021, 21(1):526.

41. Chen X, Hou X, Bai D, Lane R, Zhang C, Canuso C, Wang G, Fu DJ: Efficacy and Safety of Flexibly Dosed Esketamine Nasal Spray Plus a Newly Initiated Oral Antidepressant in Adult Patients with Treatment-Resistant Depression: A Randomized, Double-Blind, Multicenter, Active-Controlled Study Conducted in China and USA. Neuropsychiatr Dis Treat 2023, 19:693–707.

42. Sterne JA, Sutton AJ, Ioannidis JP, Terrin N, Jones DR, Lau J, Carpenter J, Rücker G, Harbord RM, Schmid CH et al: Recommendations for examining and interpreting funnel plot asymmetry in meta-analyses of randomised controlled trials. BMJ (Clinical research ed) 2011, 343:d4002.

43. Hengartner MP, Plöderl M: Estimates of the minimal important difference to evaluate the clinical significance of antidepressants in the acute treatment of moderate-to-severe depression. BMJ Evid Based Med 2022, 27(2):69–73.

44. Pharmacoeconomic Report: Esketamine Hydrochloride (Spravato): (Janssen Inc.): Indication: Major Depressive Disorder in Adults [Internet]. Ontario, Canada; 2021.

45. Canady VA: FDA approves esketamine treatment for MDD, suicidal ideation. Mental Health Weekly 2020, 30(31):6–7.

46. Kopec JA, Abrahamowicz M, Esdaile JM: Randomized discontinuation trials: utility and efficiency. Journal of clinical epidemiology 1993, 46(9):959–971.

47. Terao I, Tsuge T, Endo K, Kodama W: Comparative efficacy, tolerability and acceptability of intravenous racemic ketamine with intranasal esketamine, aripiprazole and lithium as augmentative treatments for treatment-resistant unipolar depression: A systematic review and network meta-analysis. Journal of affective disorders 2024, 346:49–56.

48. Saelens J, Gramser A, Watzal V, Zarate CA, Jr., Lanzenberger R, Kraus C: Relative effectiveness of antidepressant treatments in treatment-resistant depression: a systematic review and network meta-analysis of randomized controlled trials. Neuropsychopharmacology 2025, 50(6):913–919.

49. Cleare AJ, Kerr-Gaffney J, Goldsmith K, Zenasni Z, Yaziji N, Jin H, Colasanti A, Geddes JR, Kessler D, McAllister-Williams RH et al: Clinical and cost-effectiveness of lithium versus quetiapine augmentation for treatment-resistant depression: a pragmatic, open-label, parallel-group, randomised controlled superiority trial in the UK. The lancet Psychiatry 2025, 12(4):276–288.

50. Taillefer de Laportalière T, Horowitz MA, Soeiro T, Lepetit M, Jullien A, Yrondi A, Moncrieff J, Naudet F, Montastruc F: Association between major cardiovascular events and esketamine: A disproportionality analysis in the WHO pharmacovigilance database. Eur Neuropsychopharmacol 2025, 96:12–14.

51. Lemarchand C, Chopin R, Paul M, Braillon A, Cosgrove L, Cristea I, Fried EI, Turner EH, Naudet F: Fragile promise of psychedelics in psychiatry. BMJ (Clinical research ed) 2024, 387:e080391.

