## Supplementary material for "Efficacy and safety of esketamine for “treatment resistant depression”: registered report for a Systematic Review with an Individual Patient Data Meta-analysis of Randomized, Double-Blind, Placebo-Controlled Trials": Web appendix 1

**e-BOX 1: Thase and Rush staging method of treatment resistant depression**

Stage I: Failure of 1 adequate trial of 1 major class of antidepressants

Stage II: Failure of 2 adequate trial of 2 major class of antidepressants

Stage III: Stage II resistance + failure of an adequate trial of a tricyclic agent

Stage IV: Stage III resistance + failure of an adequate trial of a monoamine oxidase inhibitor

Stage V: Stage IV resistance + a course of bilateral electroconvulsive therapy

**e-BOX 2 General strategy for reanalyses:**

We followed a general plan detailed in a previous re-analysis project (https://bmcmedicine.biomedcentral.com/articles/10.1186/s12916-022-02377-2).

Reanalyses of the primary outcome(s) of each study was performed by a statistician who had no access to study reports, journal publications, statistical analysis plan, or analytical code, in order to ensure that the analysis was as blind as possible to the primary analysis. In addition, this reviewer was instructed not to try to find these documents or the published report.

For double-blind studies, all reanalyses were based on the latest version of the protocol issued before database lock and unblinding. If this information was not available, the date of the last visit of the last patient will be used as a proxy. In case of missing information for these dates, the study authors had to be contacted.

Although in therapeutic research statistical analysis is fairly simple, in some cases the reanalyses can involve difficult methodological choices. An independent senior statistician was available to discuss any difficult aspect or choice in the analysis plan before the reanalysis, so as to choose the most agreed upon analyses (e.g. intention-to-treat population for a superiority trial). Should insufficient information concerning the main analysis was provided in the protocol, the best practices for clinical research were used, following the ICH guidelines.

An analysis plan was developed for each study included and was recorded on the Open Science Framework prior any analysis (https://osf.io/z2bna/).

In the supplementary material e-Box 1, Table is provided with details of what was taken from the ICH guidelines in case of missing information.

Reanalyses entailed the following steps: 1/ identification of the outcome of interest (and detection of outcome switching), 2/ definition of the study population, 3/ reanalysis of the primary outcome. Any change identified between the first version of the protocol and the version used for the reanalysis of the primary outcome was tracked and described.

All results of these analyses were reported in terms of 1/ p-value, 2/ effect size and 3/ changes from the initial protocol.

These results were first compared with the results of the analyses reported in the initial study reports, and if not available, with the publications. All results from all available documents were gathered (FDA reports, European public assessment reports, study reports & publications) and were presented in the results section.

Because interpreting an RCT involves clinical expertise, and cannot be reduced to solely quantitative factors, an in-depth discussion between two researchers not involved in the re-analysis (CP and FN), based on both quantitative and qualitative (clinical judgment) factors enabled a decision on whether the changes in results described quantitatively could materialize into a change in conclusions.

If these two reviewers judged that the conclusions were the same, the study was considered as reproduced. If these two researchers judge that the conclusions are not the same, then the researcher in charge of the analysis was given the statistical analysis plan of the study and was asked to list the differences in terms of analysis. If the researcher finds a discrepancy between the study data analysis plan and their own analysis plan, the discrepancy was corrected in the analysis (e.g. analysis population, use of covariates). Again, an in-depth discussion between two researchers not involved in the reanalysis supported a decision on whether the changes in results described quantitatively could materialize into changes in conclusions, and whether the differences with respect to the analytical plan are understandable and acceptable.

If these two researchers judge that the conclusions were not the same or that the differences in the analytical plans are neither justified nor desirable, then a senior statistician had to perform their own reanalysis. He had to be kept blind to which treatment is which. Subsequently, a final in-depth discussion between two researchers not involved in the reanalysis based on the senior statistician's reanalysis supported a decision on whether the changes in results described quantitatively could materialize into a change in conclusions. Should the study be considered as not reproduced, the authors/ sponsors of the study had to be contacted to better understand the discrepancy, and this entire process was carefully documented.

**e-Box 1, Table: Example of a study analysis plan**

|  | | Information in the study documents | If present, is it substandard? | If not present,  replace according to | Information used in the reanalysis |
| --- | --- | --- | --- | --- | --- |
| General information | Study title |  | - | - |  |
|  | Registration number |  | - | - |  |
|  | Protocol and amendments number |  | - | - |  |
|  | Name of the sponsor |  | - | - |  |
|  | Trial objectives (PICO) |  |  | - |  |
|  | Type of documents available |  | - | - |  |
| Trial design | Design configuration |  |  | ICH E9, Point 3.1 |  |
|  | Type of comparison |  |  | ICH E9, Point 3.3  ICH E10 |  |
|  | Multicenter trial |  |  | ICH E9, Point 3.2 |  |
|  | Primary outcomes |  |  | ICH E9 |  |
|  | Sample size |  |  | ICH E9, point 3,4 |  |
|  | Methods to minimize bias |  |  | ICH E9, point 2.3  ICH E10, point 1.2 |  |
|  | Effect size |  |  | ICH E9, point 3.5 |  |
|  | Confidence interval |  |  | ICH E9, point 5.5 |  |
|  | Interim analysis |  |  | ICH E9, point 4.5 |  |
|  | Analysis method |  |  | ICH E9, point 5 |  |
|  | Handling missing data |  |  | ICH E9, point 5.3 |  |
|  | Hypothesis testing |  |  | ICH E9, point 5.5 |  |
|  | Statistical packages used for analysis |  |  | - |  |
|  | Protocol violations |  |  | ICH E9, point 2.3  ICH E6, point 6 |  |
|  | Analysis sets |  |  | ICH E9, point 5.2 |  |
| Funders | Funder type (commercial/non commercial) |  |  |  |  |
|  | Name |  |  |  |  |

**e-Table 1: Search strategy**

| **#** | **Searches** |
| --- | --- |
| **MEDLINE via PubMed** |  |
|  | (esketamine OR 's-ketamine') AND depress* |
| **The Cochrane Library** |  |
|  | (esketamine OR 's-ketamine') AND depress* |
| **EMBASE** |  |
|  | (esketamine OR 's-ketamine') AND depress* |
| **Clinicaltrials.gov** |  |
|  | Esketamine (also search for S-ketamine) |
| **Clinicaltrialsregister.eu** |  |
|  | s-ketamine OR esketamine |
| **EMA** |  |
|  | https://www.ema.europa.eu/en/medicines/human/EPAR/spravato |
| **FDA** |  |
|  | https://www.accessdata.fda.gov/scripts/cder/daf/ |

**e-Table 2: studies available on the YODA repository at the time of submission**

| **NCT** | **Title** | **Sample size** | **MADRS** | **Eligibility** |
| --- | --- | --- | --- | --- |
| NCT01627782 | A Double-blind, Randomized, Placebo-controlled, Parallel Group, Dose Frequency Study of Ketamine in Subjects With Treatment-resistant Depression | 68 | Yes | No, ketamine |
| NCT01640080 | A Double-Blind, Double-Randomization, Placebo-Controlled Study of the Efficacy of Intravenous Esketamine in Adult Subjects With Treatment-Resistant Depression | 30 | Yes | No, intravenous esketamine |
| NCT01998958 | A Double-Blind, Doubly-Randomized, Placebo-Controlled Study of Intranasal Esketamine in an Adaptive Treatment Protocol to Assess Safety and Efficacy in Treatment-Resistant Depression (SYNAPSE) | 108 | Yes | Possibly, depending on the inclusion criteria |
| NCT02133001 | A Double-blind, Randomized, Placebo Controlled Study to Evaluate the Efficacy and Safety of Intranasal Esketamine for the Rapid Reduction of the Symptoms of Major Depressive Disorder, Including Suicidal Ideation, in Subjects Who Are Assessed to be at Imminent Risk for Suicide | 68 | Yes | Possibly, depending on the inclusion criteria |
| NCT02417064 | A Randomized, Double-blind, Multicenter, Active-controlled Study to Evaluate the Efficacy, Safety, and Tolerability of Fixed Doses of Intranasal Esketamine Plus an Oral Antidepressant in Adult Subjects With Treatment-resistant Depression | 346 | Yes | Yes, pivotal |
| NCT02418585 | A Randomized, Double-blind, Multicenter, Active-controlled Study to Evaluate the Efficacy, Safety, and Tolerability of Flexible Doses of Intranasal Esketamine Plus an Oral Antidepressant in Adult Subjects With Treatment-resistant Depression | 236 | Yes | Yes, pivotal |
| NCT02422186 | A Randomized, Double-blind, Multicenter, Active-controlled Study to Evaluate the Efficacy, Safety, and Tolerability of Intranasal Esketamine Plus an Oral Antidepressant in Elderly Subjects With Treatment-resistant Depression | 139 | Yes | Yes, pivotal |
| NCT02493868 | A Randomized, Double-blind, Multicenter, Active-Controlled Study of Intranasal Esketamine Plus an Oral Antidepressant for Relapse Prevention in Treatment-resistant Depression | 719 | Yes | Yes, continuation |
| NCT02497287 | An Open-label, Long-term, Safety and Efficacy Study of Intranasal Esketamine in Treatment-resistant Depression | 802 | Yes | No, open label, single Group Assignment |
| NCT02918318 | A Randomized, Double-blind, Multicenter, Placebo-controlled Study to Evaluate the Efficacy, Safety and Tolerability of Fixed Doses of Intranasal Esketamine in Japanese Subjects With Treatment Resistant Depression | 202 | Yes | Possibly, depending on the inclusion criteria |
| NCT03039192 | A Double-blind, Randomized, Placebo-controlled Study to Evaluate the Efficacy and Safety of Intranasal Esketamine in Addition to Comprehensive Standard of Care for the Rapid Reduction of the Symptoms of Major Depressive Disorder, Including Suicidal Ideation, in Adult Subjects Assessed to be at Imminent Risk for Suicide | 226 | Yes | Possibly, depending on the inclusion criteria |
| NCT03097133 | A Double-blind, Randomized, Placebo-controlled Study to Evaluate the Efficacy and Safety of Intranasal Esketamine in Addition to Comprehensive Standard of Care for the Rapid Reduction of the Symptoms of Major Depressive Disorder, Including Suicidal Ideation, in Adult Subjects Assessed to be at Imminent Risk for Suicide | 230 | Yes | Possibly, depending on the inclusion criteria |

**e-Table 3:** Comparison of our re-analyses with FDA and European assessment reports

|  | | | **Re-analysis** | | | | **EMA** | | | | | **FDA** | | | | |
| --- | --- | --- | --- | --- | --- | --- | --- | --- | --- | --- | --- | --- | --- | --- | --- | --- |
| **Study** | **Outcome** | **Group** | **ES** | **LOWER** | **UPPER** | **P_value** | **ES** | **LOWER** | **UPPER** | **P_value** | **COMMENT** | **ES** | **LOWER** | **UPPER** | **P_value** | **COMMENT** |
| NCT02918318 | MADRS D-28 | Esketamine 28 mg | 0.5 | -4.22 | 5.22 | 0.741 | NA |  |  |  |  |  |  |  |  |  |
| NCT02918318 | MADRS D-28 | Esketamine 56 mg | 1.4 | -3.21 | 6.01 | 0.9527 | NA |  |  |  |  |  |  |  |  |  |
| NCT02918318 | MADRS D-28 | Esketamine 84 mg | -1.4 | -5.79 | 2.99 | 0.7817 | NA |  |  |  |  |  |  |  |  |  |
| NCT01998958 | MADRS D-8, PANEL A | Esketamine 28 mg | -5 | -10.88 | 0.88 | 0.1415 | NA | NA | NA | NA | Combination of both periods | -5 | NA | NA | 0,051 | Reproduced |
| NCT01998958 | MADRS D-8, PANEL A | Esketamine 56 mg | -7.6 | -13.284 | -1.92 | 0.0164 | NA | NA | NA | NA | Combination of both periods | -7,6 | NA | NA | 0,006 | Reproduced |
| NCT01998958 | MADRS D-8, PANEL A | Esketamine 84 mg | -10.5 | -15.988 | -5.01 | 0.001 | NA | NA | NA | NA | Combination of both periods | -10,5 | NA | NA | 0,001 | Reproduced |
| NCT01998958 | MADRS D-8, PANEL A, 2nd randomisation | Esketamine 28 mg | -5.1 | -11.764 | 3.01 | 0.2142 | NA | NA | NA | NA | Combination of both periods | -3,1 | NA | NA | 0,152 | Reproduced |
| NCT01998958 | MADRS D-8, PANEL A, 2nd randomisation | Esketamine 56 mg | -5.2 | -12.06 | 1.66 | 0.2176 | NA | NA | NA | NA | Combination of both periods | -4,4 | NA | NA | 0,083 | Reproduced |
| NCT01998958 | MADRS D-8, PANEL A, 2nd randomisation | Esketamine 84 mg | -9 | -16.06 | -1.94 | 0.0323 | NA | NA | NA | NA | Combination of both periods | -6,9 | NA | NA | 0,028 | Reproduced |
| NCT01998958 | MADRS D-8, PANEL B | Esketamine 14 mg | 1.8 | -3.34 | 6.93 | 0.9381 | NA | NA | NA | NA | NA | NA | NA | NA | NA | NA |
| NCT01998958 | MADRS D-8, PANEL B | Esketamine 56 mg | -3.73 | -9.24 | 1.78 | 0.1932 | NA | NA | NA | NA | NA | NA | NA | NA | NA | NA |
| NCT01998958 | MADRS D-8, PANEL B, 2nd randomisation | Esketamine 14 mg | -7.9 | -18.17 | 2.37 | 0.1697 | NA | NA | NA | NA | NA | NA | NA | NA | NA | NA |
| NCT01998958 | MADRS D-8, PANEL B, 2nd randomisation | Esketamine 56 mg | -0.1 | -12.27 | 12.07 | 0.7438 | NA | NA | NA | NA | NA | NA | NA | NA | NA | NA |
| NCT02417064 | MADRS D-28 | Esketamine 56 mg | -3.98 | -7.57 | -0.39 | NA | -4.1 | -7,53 | -0,6 | NA | Reproduced despite use of ANOVA | -4,12 | -7,66 | -0,58 | 0,02 | Should not have been tested |
| NCT02417064 | MADRS D-28 | Esketamine 84 mg | -3.66 | -7.33 | 0.01 | 0.051 | -2 | -5,52 | 1,42 | 0.25 | Reproduced despite use of ANOVA | -3,34 | -6,95 | 0,27 | 0,07 | Reproduced |
| NCT02418585 | MADRS D-28 | Esketamine, flexible dose | -4.3 | -7.53 | -1.06 | 0.0049 | -3,5 | -6,67 | -0,26 | 0.034 | Reproduced despite use of ANOVA | -3,98 | -7,46 | -0,5 | 0,0225 | Reproduced |
| NCT02493868 | RELAPSE | Esketamine, flexible dose | 0.46 | 0.28 | 0.77 | 0.003 | 0,49 | 0,29 | 0,84 | 0.003 | Reproduced | 0,49 | 0,29 | 0,83 | 0,0029 | Reproduced |
| NCT02422186 | MADRS D-28 | Esketamine, flexible dose | -4.17 | -7.85 | -0.49 | 0.067 | -3,6 | -7,16 | -0,03 | 0.052 | Reproduced despite use of ANOVA | -3,61 | -7,25 | 0,02 | 0,052 | Reproduced |
