## Supplementary material for "Efficacy and safety of esketamine for “treatment resistant depression”: registered report for a Systematic Review with an Individual Patient Data Meta-analysis of Randomized, Double-Blind, Placebo-Controlled Trials": Web appendix 2

### Statistical Analysis Plan (SAP)

**Version 1**

| | |
|---|---|
| **Full study title** | Efficacy and safety of esketamine for "treatment resistant depression": registered report for a Systematic Review with an Individual Participant Data Meta-analysis of Randomized, Double-Blind, Placebo-Controlled Trials |
| **Acronym** | ESK-T-DEP |
| **Prospero registry number** | CRD42021290721 |
| **OSF registration** | https://osf.io/xetvn |
| **Study protocol version** | V1.0 |
| **SAP version** | V1 |

**SAP revision history**

| Date | Document | Version | Description of and reason for change |
|---|---|---|---|
| 08.2022 | ESK-T-DEP_SAP | V1 | Creation of the document |

#### Roles and responsibility

| | |
|---|---|
| Author | Name, date, and signature:<br><br>Claude PELLEN<br>20/10/2022 |
| Senior statistician responsible | Name, date, and signature:<br><br>Estelle LE PABIC<br>19/10/2022 |
| Coordinator | Name, date, and signature:<br><br>Florian NAUDET<br>20/10/2022 |

### 1 Objectives

The main objective is to independently (independently from the sponsor) reappraise the efficacy of esketamine in "treatment resistant depression" (TRD) using an individual participant data meta-analysis methodology.

The secondary objectives are:
- to independently reappraise the safety of esketamine in the treatment of TRD
- and to explore moderating factors of esketamine efficacy and safety, including level of treatment resistance, participant age, site-specific effects, among others.

### 2 Study methods

#### 2.1 Design

Systematic review and meta-analysis of individual participant data (IPD) derived from double-blind, randomised, placebo-controlled [placebo or active placebo] trials for the indication of TRD.

#### 2.2 Participants, interventions, comparators, outcomes, and study design (PICOS)

**Participants**
Participants with TRD.
TRD refers to a depressive episode with inadequate response to at least two antidepressant trials of adequate doses and duration (i.e., have failed ≥ 2 adequate antidepressant trials). This definition includes "higher" level of "resistance" according to Thase and Rush staging method of treatment resistant depression.

There will be no age limit.

**Interventions**
Intranasal esketamine (with the following approved doses: 56 mg or 84 mg or "flexible" doses) for the primary treatment of depression.

**Comparators**
Placebo (a solution with a bittering agent added to simulate the taste of the esketamine solution) or active placebo.

**Study design**
Randomised controlled trials (initiation and continuation (i.e. maintenance) trials).

### 3 Trial population

#### 3.1 Search strategy

Systematic searches will be conducted using PubMed, the Cochrane Central Register of Controlled Trials (CENTRAL: https://www.cochranelibrary.com/central/about-central), Embase, ClinicalTrials.gov, Clinicaltrialsregister.eu, FDA, EMA and the manufacturer website. All records will be managed (and deduplicated) using Endnote.

#### 3.2 Data collection and assessment of risk of bias

Selection and coding of the different study characteristics will independently be performed by two reviewers . A third reviewer will arbitrate in case of disagreement. A data extraction sheet based on the Cochrane Handbook for Systematic Reviews of Interventions guidelines will be developed.

Studies appearing to duplicate authors, treatment comparisons, sample sizes and outcomes will be checked against one other to avoid any duplicates and to avoid integrating data from several reports on the same study.
For each included study, information will be extracted regarding: a) characteristics of the study [year, country, selection criteria, co-treatments (psychological interventions and their type), number of arms, funding]; b) type of intervention (treatments and comparators, duration). Importantly, mean characteristics of the trial participants and outcome measures will not be extracted, as we will rely on IPD.

Two researchers will independently assess each trial for risk of bias according to the Cochrane Collaboration tool for assessing risk of bias in its current version, RoB2 [21].

A data sharing request (IPD, analytical code, metadata, study reports, statistical analysis plan and any other relevant documents) will be sent to the relevant data sharing platforms and/or sponsor and /or authors. Importantly, data and metadata of the pivotal trials are available on the YODA project (https://yoda.yale.edu/jj-available-data).

#### 3.3 Flowchart

In terms of strategy the systematic review search criteria will include all studies of esketamine for the primary treatment of depression. The IPD analysis will then be restricted to participants with TRD within the included studies (i.e., have failed > 2 adequate antidepressant trials).

The PRISMA-IPD flowchart template will be used.

### 4 Analysis

#### 4.1 Generality

##### 4.1.1 Software

All the analyses will be performed with R (R core Team, 2021).

##### 4.1.2 Identification of variables of interest

See the appendix table “List of variables of interest”.

#### 4.2 Reanalysis of sponsor’s primary outcome

Reanalysis of the primary outcomes(s) of the study following the initial protocol and the International Council for Harmonisation of Technical Requirements for Pharmaceuticals for Human Use by a statistician who will have no access to study reports, journal publications, statistical analysis plan, or analytical code.
Reanalyses will be based on the latest version of the protocol issued before database lock and unblinding. If this information is not available, the date of the last visit of the last participant will be used as a proxy. In case of missing information for these dates, the study authors will be contacted.
Below are the tables ad figures to be completed during the reanalysis.

| |
|---|
| Date of database lock |
| Date of database unblinding |
| Date of the last visit of the last participant |
| Date and version of the latest version of the protocol issued before database lock and unblinding |

*Table 1: Dates of database lock, unblinding, last visit of the last participant and latest version of the protocol*

| | **Information in the study documents** | **If not present, replace according to ICH section** | **Information used in the reanalysis** |
|---|---|---|---|
| **Objectives and hypotheses** | | ICH E6 | |
| **Analysis method** | | ICH E9, point 5 | |
| **Handling missing data** | | ICH E9, point 5.3 | |
| **Protocol violations** | | ICH E9, point 2.3<br>ICH E6, point 6 | |
| **Analysis sets** | | ICH E9, point 5.2 | |

*Table 2: Statistical analysis plan of study*

**Description of any deviations from the protocol**
Any change, especially outcome switching, identified between the first version of the protocol and the version used for the reanalysis of the primary outcome will be tracked and described.

| Studies | Consent | Selection | Follow-up | Treatment | Others | At least one deviation |
|---|---|---|---|---|---|---|
| Study 1 | N | N | N | N | N | N |

*Table 3: Description of the deviations per study*

*Table 4: List of deviations per study with no subject ID*

**Results of the reanalysis**

| |
|---|
| **P-value** |
| **Effect size** |
| **Changes from the initial protocol** |

*Table 5: Results of the reanalysis*

**Exploration of center effect**

Center effect is explored by performing sensitivity analyses removing a center at a time.

| | Without centre 1 | Without centre 2 | Without centre 3 | … |
|---|---|---|---|---|
| **P-value** | | | | |
| **Effect size** | | | | |

*Table 6: Results of the sensitivity analyses*

X-axis: effect size by increasing values
Y-axis: each sensitivity analysis

*Figure 1*

**Data integrity issues mentioned in the FDA appraisal**

Not performed by the statistician who reanalyses the studies.

**Comparison of the analysis results with the analyses reported in the FDA reports, European public assessment reports, study reports & publications**

Not performed by the statistician who reanalyses the studies.

| Document | P-value | Effect size |
|---|---|---|

*Table 7: Results from all available documents (FDA reports, European public assessment reports, study reports & publications)*

#### 4.3 Meta-analyses

##### 4.3.1 Initiation studies

###### 4.3.1.1 Efficacy outcomes

###### 4.3.1.1.1 Meta-analysis of primary outcome

**MADRS after at least 4 weeks**

| | |
|---|---|
| Type of outcome | Continuous outcome. |
| Meta-analysis | Two-stage IPD meta-analysis of continuous outcome<br>- First stage:<br>o Methods: in line with each reanalysis (cf. section 4.2). The participants will be analysed according to their allocated treatment at baseline (ITT principle) and not the treatment they receive at the time of outcome assessment.<br>o Results: mean final score, standard deviation, number of subjects, per group.<br>- Second stage:<br>o Package *meta*, function *metacont*.<br>o Inverse variance method.<br>o Summary measure: mean difference.<br>o Random effects will be used in case of heterogeneity defined as an $I^2$ index > 25% and/or significant heterogeneity ($p < 0.10$) detected using Cochran's Q test statistic.<br>o In case of random effects, restricted maximum likelihood estimation (REML) estimation will be used and confidence interval will be derived by the Hartung-Knapp Sidik-Jonkman (HKSJ) approach.<br><br>Mean differences in MADRS observed (point estimates and confidence intervals) will be compared with 0 (absence of difference) and 6.5 points (the threshold defined *a priori* in initiation studies) on a forest plot (visual comparison). |
| Sensitivity analysis | A sensitivity analysis including the studies from which IPD are not available will be performed to explore the robustness of findings observed in the IPD meta-analysis. |
| Plot | Forest plot.<br>Funnel plot if applicable (> 10 studies). |
| Certainty of evidence | Rated by GRADE; not performed by the statistician. |

###### 4.3.1.1.2 Meta-analysis of secondary outcomes

###### 4.3.1.1.2.1 Outcome assessed at week 1

**MADRS at week 1**

| | |
|---|---|
| Type of outcome | Continuous outcome. |
| Meta-analysis | Two-stage IPD meta-analysis for continuous outcome<br>- First stage:<br>o Methods: in line with each reanalysis. The participants will be analysed according to their allocated treatment at baseline (ITT |

| | |
|---|---|
| | principle) and not the treatment they receive at the time of outcome assessment.<br>o Results: mean final score, standard deviation, number of subjects, per group.<br>- Second stage:<br>o Package *meta*, function *metacont*.<br>o Inverse variance method.<br>o Summary measure: mean difference.<br>o Random effects will be used in case of heterogeneity defined as an $I^2$ index > 25% and/or significant heterogeneity ($p < 0.10$) detected using Cochran's Q test statistic.<br>o In case of random effects, restricted maximum likelihood estimation (REML) estimation will be used and confidence interval will be derived by the Hartung-Knapp Sidik-Jonkman (HKSJ) approach.<br><br>Mean differences in MADRS observed (point estimates and confidence intervals) will be compared with 0 (absence of difference) and 6.5 points (the threshold defined *a priori* in initiation studies) on a forest plot (visual comparison). |
| Sensitivity analysis | A sensitivity analysis including the studies from which IPD are not available will be performed to explore the robustness of findings observed in the IPD meta-analysis. |
| Plot | Forest plot.<br>Funnel plot if applicable (> 10 studies). |
| Level of evidence | Rated by GRADE; not performed by the statistician. |

###### 4.3.1.1.2.2 Outcomes assessed at the end of treatment

**Suicide/suicide attempt**

| | |
|---|---|
| Type of outcome | Binary outcome. |
| Meta-analysis | Two-stage IPD meta-analysis for binary outcome<br>- First stage:<br>o Methods: in line with each reanalysis. The participants will be analysed according to their allocated treatment at baseline (ITT principle) and not the treatment they receive at the time of outcome assessment.<br>o Results: number of subjects with events, total number of subjects, per group.<br>- Second stage:<br>o Package meta, function metabin.<br>o Inverse variance method.<br>o Summary measure: relative risk.<br>o Random effects will be used in case of heterogeneity defined as an $I^2$ index > 25% and/or significant heterogeneity ($p < 0.10$) detected using Cochran's Q test statistic.<br>o In case of random effects, restricted maximum likelihood estimation (REML) estimation will be used and confidence interval will be derived by the Hartung-Knapp Sidik-Jonkman (HKSJ) approach. |

| | |
|---|---|
| Sensitivity analysis | A sensitivity analysis including the studies from which IPD are not available will be performed to explore the robustness of findings observed in the IPD meta-analysis. |
| Plot | Forest plot.<br>Funnel plot if applicable (> 10 studies). |
| Level of evidence | Rated by GRADE; not performed by the statistician. |

**Suicidal ideations, e.g. Beck Scale for Suicidal Ideation (BSS)**

| | |
|---|---|
| Type of outcome | Continuous outcome. |
| Meta-analysis | Two-stage IPD meta-analysis for continuous outcome<br>- First stage:<br>o Methods: in line with each reanalysis. The participants will be analysed according to their allocated treatment at baseline (ITT principle) and not the treatment they receive at the time of outcome assessment.<br>o Results: mean final score, standard deviation, number of subjects, per group.<br>- Second stage:<br>o Package meta, function metacont.<br>o Inverse variance method.<br>o Summary measure: mean difference.<br>o Random effects will be used in case of heterogeneity defined as an $I^2$ index > 25% and/or significant heterogeneity ($p < 0.10$) detected using Cochran's Q test statistic.<br>o In case of random effects, restricted maximum likelihood estimation (REML) estimation will be used and confidence interval will be derived by the Hartung-Knapp Sidik-Jonkman (HKSJ) approach. |
| Sensitivity analysis | A sensitivity analysis including the studies from which IPD are not available will be performed to explore the robustness of findings observed in the IPD meta-analysis. |
| Plot | Forest plot.<br>Funnel plot if applicable (> 10 studies). |
| Level of evidence | Rated by GRADE; not performed by the statistician. |

**Remission (MADRS score <8 if no binary)**

| | |
|---|---|
| Type of outcome | Binary outcome. |
| Meta-analysis | Two-stage IPD meta-analysis for binary outcome<br>- First stage:<br>o Methods: in line with each reanalysis. The participants will be analysed according to their allocated treatment at baseline (ITT principle) and not the treatment they receive at the time of outcome assessment.<br>o Results: number of subjects with events, total number of subjects, per group.<br>- Second stage:<br>o Package meta, function metabin.<br>o Inverse variance method.<br>o Summary measure: relative risk. |

| | |
|---|---|
| | o Random effects will be used in case of heterogeneity defined as an $I^2$ index > 25% and/or significant heterogeneity ($p < 0.10$) detected using Cochran's Q test statistic.<br>o In case of random effects, restricted maximum likelihood estimation (REML) estimation will be used and confidence interval will be derived by the Hartung-Knapp Sidik-Jonkman (HKSJ) approach. |
| Sensitivity analysis | A sensitivity analysis including the studies from which IPD are not available will be performed to explore the robustness of findings observed in the IPD meta-analysis. |
| Plot | Forest plot.<br>Funnel plot if applicable (> 10 studies). |
| Level of evidence | Rated by GRADE; not performed by the statistician. |

**Sheehan Disability Scale**

| | |
|---|---|
| Type of outcome | Continuous outcome. |
| Meta-analysis | Two-stage IPD meta-analysis for continuous outcome<br>- First stage:<br>o Methods: in line with each reanalysis. The participants will be analysed according to their allocated treatment at baseline (ITT principle) and not the treatment they receive at the time of outcome assessment.<br>o Results: mean final score, standard deviation, number of subjects, per group.<br>- Second stage:<br>o Package meta, function metacont.<br>o Inverse variance method.<br>o Summary measure: mean difference.<br>o Random effects will be used in case of heterogeneity defined as an $I^2$ index > 25% and/or significant heterogeneity ($p < 0.10$) detected using Cochran's Q test statistic.<br>o In case of random effects, restricted maximum likelihood estimation (REML) estimation will be used and confidence interval will be derived by the Hartung-Knapp Sidik-Jonkman (HKSJ) approach. |
| Sensitivity analysis | A sensitivity analysis including the studies from which IPD are not available will be performed to explore the robustness of findings observed in the IPD meta-analysis. |
| Plot | Forest plot.<br>Funnel plot if applicable (> 10 studies). |
| Level of evidence | Rated by GRADE; not performed by the statistician. |

**PHQ-9**

| | |
|---|---|
| Type of outcome | Continuous outcome. |
| Meta-analysis | Two-stage IPD meta-analysis for continuous outcome<br>- First stage:<br>o Methods: in line with each reanalysis. The participants will be analysed according to their allocated treatment at baseline (ITT principle) and not the treatment they receive at the time of outcome assessment. |

| | |
|---|---|
| | o Results: mean final score, standard deviation, number of subjects, per group.<br>- Second stage:<br>o Package meta, function metacont.<br>o Inverse variance method.<br>o Summary measure: mean difference.<br>o Random effects will be used in case of heterogeneity defined as an $I^2$ index > 25% and/or significant heterogeneity ($p < 0.10$) detected using Cochran's Q test statistic.<br>o In case of random effects, restricted maximum likelihood estimation (REML) estimation will be used and confidence interval will be derived by the Hartung-Knapp Sidik-Jonkman (HKSJ) approach. |
| Sensitivity analysis | A sensitivity analysis including the studies from which IPD are not available will be performed to explore the robustness of findings observed in the IPD meta-analysis. |
| Plot | Forest plot.<br>Funnel plot if applicable (> 10 studies). |
| Level of evidence | Rated by GRADE; not performed by the statistician. |

###### 4.3.1.1.2.3 Outcomes assessed at the last follow-up visit post-treatment

**Description of treatments received**

| | | **Treatment received at the last follow-up visit** | | |
|---|---|---|---|---|
| | | **Esketamine** | **Placebo** | **Total** |
| **Treatment allocation at randomisation** | **Esketamine** | N | N | N |
| | **Placebo** | N | N | N |
| | **Total** | N | N | N |

*Table 8: Description of treatment allocations and treatments received at the last follow-up visit*

**Suicide/suicide attempt**

| | |
|---|---|
| Type of outcome | Binary outcome. |
| Meta-analysis | Two-stage IPD meta-analysis for binary outcome<br>- First stage:<br>o Methods: in line with each reanalysis. The participants will be analysed according to their allocated treatment at baseline (ITT principle) and not the treatment they receive at the time of outcome assessment.<br>o Results: number of subjects with events, total number of subjects, per group.<br>- Second stage:<br>o Package meta, function metabin.<br>o Inverse variance method.<br>o Summary measure: relative risk.<br>o Random effects will be used in case of heterogeneity defined as an $I^2$ index > 25% and/or significant heterogeneity ($p < 0.10$) detected using Cochran's Q test statistic.<br>o In case of random effects, restricted maximum likelihood estimation (REML) estimation will be used and confidence |

| | |
|---|---|
| | interval will be derived by the Hartung-Knapp Sidik-Jonkman (HKSJ) approach. |
| Sensitivity analysis | A sensitivity analysis including the studies from which IPD are not available will be performed to explore the robustness of findings observed in the IPD meta-analysis. |
| Plot | Forest plot.<br>Funnel plot if applicable (> 10 studies). |
| Level of evidence | Rated by GRADE; not performed by the statistician. |

**Suicidal ideations, e.g. Beck Scale for Suicidal Ideation (BSS)**

| | |
|---|---|
| Type of outcome | Continuous outcome. |
| Meta-analysis | Two-stage IPD meta-analysis for continuous outcome<br>- First stage:<br>o Methods: in line with each reanalysis. The participants will be analysed according to their allocated treatment at baseline (ITT principle) and not the treatment they receive at the time of outcome assessment.<br>o Results: mean final score, standard deviation, number of subjects, per group.<br>- Second stage:<br>o Package meta, function metacont.<br>o Inverse variance method.<br>o Summary measure: mean difference.<br>o Random effects will be used in case of heterogeneity defined as an $I^2$ index > 25% and/or significant heterogeneity ($p < 0.10$) detected using Cochran's Q test statistic.<br>o In case of random effects, restricted maximum likelihood estimation (REML) estimation will be used and confidence interval will be derived by the Hartung-Knapp Sidik-Jonkman (HKSJ) approach. |
| Sensitivity analysis | A sensitivity analysis including the studies from which IPD are not available will be performed to explore the robustness of findings observed in the IPD meta-analysis. |
| Plot | Forest plot.<br>Funnel plot if applicable (> 10 studies). |
| Level of evidence | Rated by GRADE; not performed by the statistician. |

**MADRS**

| | |
|---|---|
| Type of outcome | Continuous outcome. |
| Meta-analysis | Two-stage IPD meta-analysis for continuous outcome<br>- First stage:<br>o Methods: in line with each reanalysis. The participants will be analysed according to their allocated treatment at baseline (ITT principle) and not the treatment they receive at the time of outcome assessment.<br>o Results: mean final score, standard deviation, number of subjects, per group.<br>- Second stage:<br>o Package meta, function metacont.<br>o Inverse variance method.<br>o Summary measure: mean difference. |

| | |
|---|---|
| | o Random effects will be used in case of heterogeneity defined as an $I^2$ index > 25% and/or significant heterogeneity ($p < 0.10$) detected using Cochran's Q test statistic.<br>o In case of random effects, restricted maximum likelihood estimation (REML) estimation will be used and confidence interval will be derived by the Hartung-Knapp Sidik-Jonkman (HKSJ) approach.<br><br>Mean differences in MADRS observed (point estimates and confidence intervals) will be compared with 0 (absence of difference) and 6.5 points (the threshold defined *a priori* in initiation studies) on a forest plot (visual comparison). |
| Sensitivity analysis | A sensitivity analysis including the studies from which IPD are not available will be performed to explore the robustness of findings observed in the IPD meta-analysis. |
| Plot | Forest plot.<br>Funnel plot if applicable (> 10 studies). |
| Level of evidence | Rated by GRADE; not performed by the statistician. |

**Remission (MADRS score <8 if no binary)**

| | |
|---|---|
| Type of outcome | Binary outcome. |
| Meta-analysis | Two-stage IPD meta-analysis for binary outcome<br>- First stage:<br>o Methods: in line with each reanalysis. The participants will be analysed according to their allocated treatment at baseline (ITT principle) and not the treatment they receive at the time of outcome assessment.<br>o Results: number of subjects with events, total number of subjects, per group.<br>- Second stage:<br>o Package meta, function metabin.<br>o Inverse variance method.<br>o Summary measure: relative risk.<br>o Random effects will be used in case of heterogeneity defined as an $I^2$ index > 25% and/or significant heterogeneity ($p < 0.10$) detected using Cochran's Q test statistic.<br>o In case of random effects, restricted maximum likelihood estimation (REML) estimation will be used and confidence interval will be derived by the Hartung-Knapp Sidik-Jonkman (HKSJ) approach. |
| Sensitivity analysis | A sensitivity analysis including the studies from which IPD are not available will be performed to explore the robustness of findings observed in the IPD meta-analysis. |
| Plot | Forest plot.<br>Funnel plot if applicable (> 10 studies). |
| Level of evidence | Rated by GRADE; not performed by the statistician. |

**Sheehan Disability Scale**

| | |
|---|---|
| Type of outcome | Continuous outcome. |
| Meta-analysis | Two-stage IPD meta-analysis for continuous outcome<br>- First stage: |

| | |
|---|---|
| | ○ Methods: in line with each reanalysis. The participants will be analysed according to their allocated treatment at baseline (ITT principle) and not the treatment they receive at the time of outcome assessment.<br>○ Results: mean final score, standard deviation, number of subjects, per group.<br>- Second stage:<br>○ Package meta, function metacont.<br>○ Inverse variance method.<br>○ Summary measure: mean difference.<br>○ Random effects will be used in case of heterogeneity defined as an $I^2$ index > 25% and/or significant heterogeneity ($p < 0.10$) detected using Cochran's Q test statistic.<br>○ In case of random effects, restricted maximum likelihood estimation (REML) estimation will be used and confidence interval will be derived by the Hartung-Knapp Sidik-Jonkman (HKSJ) approach. |
| Sensitivity analysis | A sensitivity analysis including the studies from which IPD are not available will be performed to explore the robustness of findings observed in the IPD meta-analysis. |
| Plot | Forest plot.<br>Funnel plot if applicable (> 10 studies). |
| Level of evidence | Rated by GRADE; not performed by the statistician. |

**PHQ-9**

| | |
|---|---|
| Type of outcome | Continuous outcome. |
| Meta-analysis | Two-stage IPD meta-analysis for continuous outcome<br>- First stage:<br>○ Methods: in line with each reanalysis. The participants will be analysed according to their allocated treatment at baseline (ITT principle) and not the treatment they receive at the time of outcome assessment.<br>○ Results: mean final score, standard deviation, number of subjects, per group.<br>- Second stage:<br>○ Package meta, function metacont.<br>○ Inverse variance method.<br>○ Summary measure: mean difference.<br>○ Random effects will be used in case of heterogeneity defined as an $I^2$ index > 25% and/or significant heterogeneity ($p < 0.10$) detected using Cochran's Q test statistic.<br>○ In case of random effects, restricted maximum likelihood estimation (REML) estimation will be used and confidence interval will be derived by the Hartung-Knapp Sidik-Jonkman (HKSJ) approach. |
| Sensitivity analysis | A sensitivity analysis including the studies from which IPD are not available will be performed to explore the robustness of findings observed in the IPD meta-analysis. |
| Plot | Forest plot.<br>Funnel plot if applicable (> 10 studies). |
| Level of evidence | Rated by GRADE; not performed by the statistician. |

4.3.1.2 Safety outcomes

4.3.1.2.1 Outcomes assessed at the end of treatment

**Serious adverse events**

| | |
|---|---|
| Type of outcome | Count outcome, i.e. number of events per participant. |
| Meta-analysis | Two-stage IPD meta-analysis for count outcome<br>- First stage:<br>o Methods: in line with each reanalysis. The participants will be analysed according to the treatment they receive at the time of outcome assessment.<br>o Results: number of events, person time at risk, per group.<br>- Second stage:<br>o Package meta, function metainc.<br>o Inverse variance method.<br>o Summary measure: incidence rate ratio.<br>o Random effects will be used in case of heterogeneity defined as an $I^2$ index > 25% and/or significant heterogeneity ($p < 0.10$) detected using Cochran's Q test statistic.<br>o In case of random effects, restricted maximum likelihood estimation (REML) estimation will be used and confidence interval will be derived by the Hartung-Knapp Sidik-Jonkman (HKSJ) approach. |
| Sensitivity analysis | A sensitivity analysis including the studies from which IPD are not available will be performed to explore the robustness of findings observed in the IPD meta-analysis. |
| Plot | Forest plot.<br>Funnel plot if applicable (> 10 studies). |
| Level of evidence | Rated by GRADE; not performed by the statistician. |

**Dropout for any cause**

| | |
|---|---|
| Type of outcome | Binary outcome. |
| Meta-analysis | Two-stage IPD meta-analysis for binary outcome<br>- First stage:<br>o Methods: in line with each reanalysis. The participants will be analysed according to the treatment they receive at the time of outcome assessment.<br>o Results: number of subjects with events, total number of subjects, per group.<br>- Second stage:<br>o Package meta, function metabin.<br>o Inverse variance method.<br>o Summary measure: relative risk.<br>o Random effects will be used in case of heterogeneity defined as an $I^2$ index > 25% and/or significant heterogeneity ($p < 0.10$) detected using Cochran's Q test statistic.<br>o In case of random effects, restricted maximum likelihood estimation (REML) estimation will be used and confidence interval will be derived by the Hartung-Knapp Sidik-Jonkman (HKSJ) approach. |

| | |
|---|---|
| Sensitivity analysis | A sensitivity analysis including the studies from which IPD are not available will be performed to explore the robustness of findings observed in the IPD meta-analysis. |
| Plot | Forest plot.<br>Funnel plot if applicable (> 10 studies). |
| Level of evidence | Rated by GRADE; not performed by the statistician. |

**Dropout due to adverse events**

| | |
|---|---|
| Type of outcome | Binary outcome. |
| Meta-analysis | Two-stage IPD meta-analysis for binary outcome<br>- First stage:<br>o Methods: in line with each reanalysis. The participants will be analysed according to the treatment they receive at the time of outcome assessment.<br>o Results: number of subjects with events, total number of subjects, per group.<br>- Second stage:<br>o Package meta, function metabin.<br>o Inverse variance method.<br>o Summary measure: relative risk.<br>o Random effects will be used in case of heterogeneity defined as an $I^2$ index > 25% and/or significant heterogeneity ($p < 0.10$) detected using Cochran's Q test statistic.<br>o In case of random effects, restricted maximum likelihood estimation (REML) estimation will be used and confidence interval will be derived by the Hartung-Knapp Sidik-Jonkman (HKSJ) approach. |
| Sensitivity analysis | A sensitivity analysis including the studies from which IPD are not available will be performed to explore the robustness of findings observed in the IPD meta-analysis. |
| Plot | Forest plot.<br>Funnel plot if applicable (> 10 studies). |
| Level of evidence | Rated by GRADE; not performed by the statistician. |

**Adverse events**

| | |
|---|---|
| Type of outcome | Count outcome, i.e. number of events per participant. |
| Meta-analysis | Two-stage IPD meta-analysis for count outcome<br>- First stage:<br>o Methods: in line with each reanalysis. The participants will be analysed according to the treatment they receive at the time of outcome assessment.<br>o Results: number of events, person time at risk, per group.<br>- Second stage:<br>o Package meta, function metainc.<br>o Inverse variance method.<br>o Summary measure: incidence rate ratio.<br>o Random effects will be used in case of heterogeneity defined as an $I^2$ index > 25% and/or significant heterogeneity ($p < 0.10$) detected using Cochran's Q test statistic.<br>o In case of random effects, restricted maximum likelihood estimation (REML) estimation will be used and confidence |

| | |
|---|---|
| | - interval will be derived by the Hartung-Knapp Sidik-Jonkman (HKSJ) approach. |
| Sensitivity analysis | A sensitivity analysis including the studies from which IPD are not available will be performed to explore the robustness of findings observed in the IPD meta-analysis. |
| Plot | Forest plot.<br>Funnel plot if applicable (> 10 studies). |
| Level of evidence | Rated by GRADE; not performed by the statistician. |

**Blood pressure**

| | |
|---|---|
| Type of outcome | Continuous outcome. |
| Meta-analysis | Two-stage IPD meta-analysis for continuous outcome<br>- First stage:<br>o Methods: in line with each reanalysis. The participants will be analysed according to the treatment they receive at the time of outcome assessment.<br>o Results: mean final score, standard deviation, number of subjects, per group.<br>- Second stage:<br>o Package meta, function metacont.<br>o Inverse variance method.<br>o Summary measure: mean difference.<br>o Random effects will be used in case of heterogeneity defined as an $I^2$ index > 25% and/or significant heterogeneity (p < 0.10) detected using Cochran's Q test statistic.<br>o In case of random effects, restricted maximum likelihood estimation (REML) estimation will be used and confidence interval will be derived by the Hartung-Knapp Sidik-Jonkman (HKSJ) approach. |
| Sensitivity analysis | A sensitivity analysis including the studies from which IPD are not available will be performed to explore the robustness of findings observed in the IPD meta-analysis. |
| Plot | Forest plot.<br>Funnel plot if applicable (> 10 studies). |
| Level of evidence | Rated by GRADE; not performed by the statistician. |

**Dissociation**

| | |
|---|---|
| Type of outcome | Binary outcome. |
| Meta-analysis | In case of dissociation is not explicitly reported, the AEs will be assessed in a blind manner (i.e. without knowledge of the treatment group) and discussed by a clinician.<br><br>Two-stage IPD meta-analysis for binary outcome<br>- First stage:<br>o Methods: in line with each reanalysis. The participants will be analysed according to the treatment they receive at the time of outcome assessment.<br>o Results: number of subjects with events, total number of subjects, per group.<br>- Second stage:<br>o Package meta, function metabin. |

| | |
|---|---|
| | ○ Inverse variance method.<br>○ Summary measure: relative risk.<br>○ Random effects will be used in case of heterogeneity defined as an $I^2$ index > 25% and/or significant heterogeneity ($p < 0.10$) detected using Cochran's Q test statistic.<br>○ In case of random effects, restricted maximum likelihood estimation (REML) estimation will be used and confidence interval will be derived by the Hartung-Knapp Sidik-Jonkman (HKSJ) approach. |
| Sensitivity analysis | A sensitivity analysis including the studies from which IPD are not available will be performed to explore the robustness of findings observed in the IPD meta-analysis. |
| Plot | Forest plot.<br>Funnel plot if applicable (> 10 studies). |
| Level of evidence | Rated by GRADE; not performed by the statistician. |

**Sedation**

| | |
|---|---|
| Type of outcome | Binary outcome. |
| Meta-analysis | In case of sedation is not explicitly reported, the AEs will be assessed in a blind manner (i.e. without knowledge of the treatment group) and discussed by a clinician.<br><br>Two-stage IPD meta-analysis for binary outcome<br>- First stage:<br>○ Methods: in line with each reanalysis. The participants will be analysed according to the treatment they receive at the time of outcome assessment.<br>○ Results: number of subjects with events, total number of subjects, per group.<br>- Second stage:<br>○ Package meta, function metabin.<br>○ Inverse variance method.<br>○ Summary measure: relative risk.<br>○ Random effects will be used in case of heterogeneity defined as an $I^2$ index > 25% and/or significant heterogeneity ($p < 0.10$) detected using Cochran's Q test statistic.<br>○ In case of random effects, restricted maximum likelihood estimation (REML) estimation will be used and confidence interval will be derived by the Hartung-Knapp Sidik-Jonkman (HKSJ) approach. |
| Sensitivity analysis | A sensitivity analysis including the studies from which IPD are not available will be performed to explore the robustness of findings observed in the IPD meta-analysis. |
| Plot | Forest plot.<br>Funnel plot if applicable (> 10 studies). |
| Level of evidence | Rated by GRADE; not performed by the statistician. |

4.3.1.2.2 Outcomes assessed at the last follow-up visit post-treatment

**Serious adverse events**

| | |
|---|---|
| Type of outcome | Count outcome, i.e. number of events per participant. |

| | |
|---|---|
| Meta-analysis | Two-stage IPD meta-analysis for count outcome<br>- First stage:<br>o Methods: in line with each reanalysis. The participants will be analysed according to the treatment they receive at the time of outcome assessment.<br>o Results: number of events, person time at risk, per group.<br>- Second stage:<br>o Package meta, function metainc.<br>o Inverse variance method.<br>o Summary measure: incidence rate ratio.<br>o Random effects will be used in case of heterogeneity defined as an $I^2$ index > 25% and/or significant heterogeneity ($p < 0.10$) detected using Cochran's Q test statistic.<br>o In case of random effects, restricted maximum likelihood estimation (REML) estimation will be used and confidence interval will be derived by the Hartung-Knapp Sidik-Jonkman (HKSJ) approach. |
| Sensitivity analysis | A sensitivity analysis including the studies from which IPD are not available will be performed to explore the robustness of findings observed in the IPD meta-analysis. |
| Plot | Forest plot.<br>Funnel plot if applicable (> 10 studies). |
| Level of evidence | Rated by GRADE; not performed by the statistician. |

**Dropout for any cause**

| | |
|---|---|
| Type of outcome | Binary outcome. |
| Meta-analysis | Two-stage IPD meta-analysis for binary outcome<br>- First stage:<br>o Methods: in line with each reanalysis. The participants will be analysed according to the treatment they receive at the time of outcome assessment.<br>o Results: number of subjects with events, total number of subjects, per group.<br>- Second stage:<br>o Package meta, function metabin.<br>o Inverse variance method.<br>o Summary measure: relative risk.<br>o Random effects will be used in case of heterogeneity defined as an $I^2$ index > 25% and/or significant heterogeneity ($p < 0.10$) detected using Cochran's Q test statistic.<br>o In case of random effects, restricted maximum likelihood estimation (REML) estimation will be used and confidence interval will be derived by the Hartung-Knapp Sidik-Jonkman (HKSJ) approach. |
| Sensitivity analysis | A sensitivity analysis including the studies from which IPD are not available will be performed to explore the robustness of findings observed in the IPD meta-analysis. |
| Plot | Forest plot.<br>Funnel plot if applicable (> 10 studies). |
| Level of evidence | Rated by GRADE; not performed by the statistician. |

**Dropout due to adverse events**

| | |
|---|---|
| Type of outcome | Binary outcome. |
| Meta-analysis | Two-stage IPD meta-analysis for binary outcome<br>- First stage:<br>o Methods: in line with each reanalysis. The participants will be analysed according to the treatment they receive at the time of outcome assessment.<br>o Results: number of subjects with events, total number of subjects, per group.<br>- Second stage:<br>o Package meta, function metabin.<br>o Inverse variance method.<br>o Summary measure: relative risk.<br>o Random effects will be used in case of heterogeneity defined as an $I^2$ index > 25% and/or significant heterogeneity ($p < 0.10$) detected using Cochran's Q test statistic.<br>o In case of random effects, restricted maximum likelihood estimation (REML) estimation will be used and confidence interval will be derived by the Hartung-Knapp Sidik-Jonkman (HKSJ) approach. |
| Sensitivity analysis | A sensitivity analysis including the studies from which IPD are not available will be performed to explore the robustness of findings observed in the IPD meta-analysis. |
| Plot | Forest plot.<br>Funnel plot if applicable (> 10 studies). |
| Level of evidence | Rated by GRADE; not performed by the statistician. |

**Adverse events**

| | |
|---|---|
| Type of outcome | Count outcome, i.e. number of events per participant. |
| Meta-analysis | Two-stage IPD meta-analysis for count outcome<br>- First stage:<br>o Methods: in line with each reanalysis. The participants will be analysed according to the treatment they receive at the time of outcome assessment.<br>o Results: number of events, person time at risk, per group.<br>- Second stage:<br>o Package meta, function metainc.<br>o Inverse variance method.<br>o Summary measure: incidence rate ratio.<br>o Random effects will be used in case of heterogeneity defined as an $I^2$ index > 25% and/or significant heterogeneity ($p < 0.10$) detected using Cochran's Q test statistic.<br>o In case of random effects, restricted maximum likelihood estimation (REML) estimation will be used and confidence interval will be derived by the Hartung-Knapp Sidik-Jonkman (HKSJ) approach. |
| Sensitivity analysis | A sensitivity analysis including the studies from which IPD are not available will be performed to explore the robustness of findings observed in the IPD meta-analysis. |
| Plot | Forest plot.<br>Funnel plot if applicable (> 10 studies). |

| | |
|---|---|
| Level of evidence | Rated by GRADE; not performed by the statistician. |

**Blood pressure**

| | |
|---|---|
| Type of outcome | Continuous outcome. |
| Meta-analysis | Two-stage IPD meta-analysis for continuous outcome<br>- First stage:<br>o Methods: in line with each reanalysis. The participants will be analysed according to the treatment they receive at the time of outcome assessment.<br>o Results: mean final score, standard deviation, number of subjects, per group.<br>- Second stage:<br>o Package meta, function metacont.<br>o Inverse variance method.<br>o Summary measure: mean difference.<br>o Random effects will be used in case of heterogeneity defined as an $I^2$ index > 25% and/or significant heterogeneity ($p < 0.10$) detected using Cochran's Q test statistic.<br>o In case of random effects, restricted maximum likelihood estimation (REML) estimation will be used and confidence interval will be derived by the Hartung-Knapp Sidik-Jonkman (HKSJ) approach. |
| Sensitivity analysis | A sensitivity analysis including the studies from which IPD are not available will be performed to explore the robustness of findings observed in the IPD meta-analysis. |
| Plot | Forest plot.<br>Funnel plot if applicable (> 10 studies). |
| Level of evidence | Rated by GRADE; not performed by the statistician. |

**Dissociation**

| | |
|---|---|
| Type of outcome | Binary outcome. |
| Meta-analysis | In case of dissociation is not explicitly reported, the AEs will be assessed in a blind manner (i.e. without knowledge of the treatment group) and discussed by a clinician.<br><br>Two-stage IPD meta-analysis for binary outcome<br>- First stage:<br>o Methods: in line with each reanalysis. The participants will be analysed according to the treatment they receive at the time of outcome assessment.<br>o Results: number of subjects with events, total number of subjects, per group.<br>- Second stage:<br>o Package meta, function metabin.<br>o Inverse variance method.<br>o Summary measure: relative risk.<br>o Random effects will be used in case of heterogeneity defined as an $I^2$ index > 25% and/or significant heterogeneity ($p < 0.10$) detected using Cochran's Q test statistic.<br>o In case of random effects, restricted maximum likelihood estimation (REML) estimation will be used and confidence |

| | |
|---|---|
| | interval will be derived by the Hartung-Knapp Sidik-Jonkman (HKSJ) approach. |
| Sensitivity analysis | A sensitivity analysis including the studies from which IPD are not available will be performed to explore the robustness of findings observed in the IPD meta-analysis. |
| Plot | Forest plot.<br>Funnel plot if applicable (> 10 studies). |
| Level of evidence | Rated by GRADE; not performed by the statistician. |

**Sedation**

| | |
|---|---|
| Type of outcome | Binary outcome. |
| Meta-analysis | In case of sedation is not explicitly reported, the AEs will be assessed in a blind manner (i.e. without knowledge of the treatment group) and discussed by a clinician.<br><br>Two-stage IPD meta-analysis for binary outcome<br>- First stage:<br>o Methods: in line with each reanalysis. The participants will be analysed according to the treatment they receive at the time of outcome assessment.<br>o Results: number of subjects with events, total number of subjects, per group.<br>- Second stage:<br>o Package meta, function metabin.<br>o Inverse variance method.<br>o Summary measure: relative risk.<br>o Random effects will be used in case of heterogeneity defined as an $I^2$ index > 25% and/or significant heterogeneity ($p < 0.10$) detected using Cochran's Q test statistic.<br>o In case of random effects, restricted maximum likelihood estimation (REML) estimation will be used and confidence interval will be derived by the Hartung-Knapp Sidik-Jonkman (HKSJ) approach. |
| Sensitivity analysis | A sensitivity analysis including the studies from which IPD are not available will be performed to explore the robustness of findings observed in the IPD meta-analysis. |
| Plot | Forest plot.<br>Funnel plot if applicable (> 10 studies). |
| Level of evidence | Rated by GRADE; not performed by the statistician. |

###### 4.3.1.3 Moderators of esketamine efficacy

Based on the individual data, the interactions will be assessed first with a two-stage approach. One-stage IPD meta-analysis may be used if necessary. The use of two-stage approach is a change from the initial protocol that was made to handle the risk for ecological bias inherent with the one-stage approach.

**Impact of resistance stage**

Each participant will be classified according to Thase and Rush resistance stages in accordance with their baseline characteristics. In case of missing data, the possibility to classify participants will be explored in a blind manner (i.e. without knowledge of the treatment group) with a clinician.

| **Stage** | **Definition** |
|---|---|
| Stage I | Failure of 1 adequate trial of 1 major class of antidepressants |
| Stage II | Failure of 2 adequate trial of 2 major class of antidepressants |
| Stage III | Stage II resistance + failure of an adequate trial of a tricyclic agent |
| Stage IV | Stage III resistance + failure of an adequate trial of a monoamine oxidase inhibitor |
| Stage V | Stage IV resistance + a course of bilateral electroconvulsive therapy |

*Table 9: Thase and Rush staging method of treatment resistant depression*

| | |
|---|---|
| Two-stage IPD meta-analysis | First stage:<br>- Outcome: MADRS after at least 4 weeks (continuous outcome).<br>- Covariate: resistance stage (quantitative outcome, 1, 2, 3, 4, 5).<br>- Estimation of the interaction term, between resistance stage and treatment group, and its variance in each study, in line with each reanalysis.<br>- The participants will be analysed according to their allocated treatment at baseline (ITT principle) and not the treatment they receive at the time of outcome assessment.<br>- Results: interaction term, standard deviation, number of subjects, per group.<br>Second stage:<br>- Outcome: interaction term (continuous outcome).<br>- Package meta, function metacont.<br>- Inverse variance method.<br>- Summary measure: interaction effect.<br>- Random effects will be used in case of heterogeneity defined as an $I^2$ index > 25% and/or significant heterogeneity ($p < 0.10$) detected using Cochran's Q test statistic.<br>- In case of random effects, restricted maximum likelihood estimation (REML) estimation will be used and confidence interval will be derived by the Hartung-Knapp Sidik-Jonkman (HKSJ) approach. |
| One-stage IPD meta-analysis | Outcome: MADRS after at least 4 weeks (continuous outcome).<br>$MADRS_{ij} = \phi_i + \beta1_i z_{ij} + \theta_i treat_{ij} + \gamma_W (treat_{ij} * (z_{ij}-\bar{z}_i)) + \varepsilon_{ij} \quad \varepsilon_{ij} \sim N(0, \sigma_i^2)$<br>i = 1 to k trials<br>j participants<br>Esketamine group ($treat_{ij} = 1$), control group ($treat_{ij} = 0$)<br>$MADRS_{ij}$ : outcome<br>$\phi_i$ : control effect (intercept term)<br>$\theta_i$ : treatment effect<br>$\varepsilon_{ij}$ : residual error (error term)<br>$\gamma_W$ = within-study interaction term<br>$z_{ij}$ : participant-level covariate<br><br>Model should allow:<br>- One intercept for each study (stratification)<br>- One treatment effect for each study (random effects) |

| | |
|---|---|
| | - The separation of within-study and across-study interaction (by centering covariates and including their mean or proportion)<br>- One within-study interaction effect for each study (random effects)<br>- One residual variance for each study (stratification)<br>- Uncorrelated random effects |
| Sensitivity analysis | A sensitivity analysis including the studies from which IPD are not available will be performed to explore the robustness of findings observed in the IPD meta-analysis. |
| Plot | Forest plot. |
| Level of evidence | Rated by GRADE; not performed by the statistician. |

**Impact of age**

| | |
|---|---|
| Two-stage IPD meta-analysis | First stage:<br>- Outcome: MADRS after at least 4 weeks (continuous outcome).<br>- Covariate: age (continuous outcome).<br>- Estimation of the interaction term, between age and treatment group, and its variance in each study, in line with each reanalysis.<br>- The participants will be analysed according to their allocated treatment at baseline (ITT principle) and not the treatment they receive at the time of outcome assessment.<br>- Results: interaction term, standard deviation, number of subjects, per group.<br>Second stage:<br>- Outcome: interaction term (continuous outcome).<br>- Package meta, function metagen.<br>- Inverse variance method.<br>- Summary measure: interaction effect.<br>- Random effects will be used in case of heterogeneity defined as an $I^2$ index > 25% and/or significant heterogeneity ($p < 0.10$) detected using Cochran's Q test statistic.<br>- In case of random effects, restricted maximum likelihood estimation (REML) estimation will be used and confidence interval will be derived by the Hartung-Knapp Sidik-Jonkman (HKSJ) approach. |
| One-stage IPD meta-analysis | Outcome: MADRS after at least 4 weeks (continuous outcome).<br>$MADRS_{ij} = \phi_i + \beta 1_i z_{ij} + \theta_i treat_{ij} + \gamma_W (treat_{ij} * (z_{ij}-\bar{z}_i)) + \varepsilon_{ij} \quad \varepsilon_{ij} \sim N(0, \sigma_i^2)$<br>i = 1 to k trials<br>j participants<br>Esketamine group ($treat_{ij} = 1$), control group ($treat_{ij} = 0$)<br>$MADRS_{ij}$ : outcome<br>$\phi_i$ : control effect (intercept term)<br>$\theta_i$ : treatment effect<br>$\varepsilon_{ij}$ : residual error (error term)<br>$\gamma_W$ = within-study interaction term<br>$z_{ij}$ : participant-level covariate<br><br>Model should allow:<br>- One intercept for each study (stratification)<br>- One treatment effect for each study (random effects)<br>- The separation of within-study and across-study interaction (by centering covariates and including their mean or proportion)<br>- One within-study interaction effect for each study (random effects) |

| | |
|---|---|
| | - One residual variance for each study (stratification)<br>Uncorrelated random effects |
| Sensitivity analysis | A sensitivity analysis including the studies from which IPD are not available will be performed to explore the robustness of findings observed in the IPD meta-analysis. |
| Plot | Forest plot. |
| Level of evidence | Rated by GRADE; not performed by the statistician. |

**Impact of per capita gross national income**

Another sensitivity analysis will be performed to explore if efficacy varies according to the World Bank categorization into low, middle, and high income (https://data.worldbank.org/country) as initial evidence on antidepressants suggests that per capita gross national income is associated with trial results.

| Study | High income country | Low, lower middle, and upper middle income country | Missing data |
|---|---|---|---|
| 1 | *Number of participants* | *Number of participants* | *Number of participants* |
| 2 | *Number of participants* | *Number of participants* | *Number of participants* |
| 3 | *Number of participants* | *Number of participants* | *Number of participants* |
| … | *Number of participants* | *Number of participants* | *Number of participants* |

*Table 10: Description of the per capita gross national income in each initiation study*

| | |
|---|---|
| Two-stage IPD meta-analysis | First stage:<br>- Outcome: MADRS after at least 4 weeks (continuous outcome).<br>- Covariate: per capita gross national income (binary outcome: high income countries vs. the other).<br>- Estimation of the interaction term, between per capita gross national income and treatment group, and its variance in each study, in line with each reanalysis.<br>- The participants will be analysed according to their allocated treatment at baseline (ITT principle) and not the treatment they receive at the time of outcome assessment.<br>- Results: interaction term, standard deviation, number of subjects, per group.<br>Second stage:<br>- Outcome: interaction term (continuous outcome).<br>- Package meta, function metagen.<br>- Inverse variance method.<br>- Summary measure: interaction effect.<br>- Random effects will be used in case of heterogeneity defined as an $I^2$ index > 25% and/or significant heterogeneity ($p < 0.10$) detected using Cochran's Q test statistic.<br>- In case of random effects, restricted maximum likelihood estimation (REML) estimation will be used and confidence interval will be derived by the Hartung-Knapp Sidik-Jonkman (HKSJ) approach. |
| One-stage IPD meta-analysis | Outcome: MADRS after at least 4 weeks (continuous outcome).<br>$MADRS_{ij} = \phi_i + \beta 1_i z_{ij} + \theta_i treat_{ij} + \gamma_W (treat_{ij} * (z_{ij}-\bar{z}_i)) + \varepsilon_{ij} \quad \varepsilon_{ij} \sim N(0, \sigma_i^2)$<br>i = 1 to k trials<br>j participants<br>Esketamine group ($treat_{ij} = 1$), control group ($treat_{ij} = 0$)<br>$MADRS_{ij}$ : outcome<br>$\phi_i$ : control effect (intercept term)<br>$\theta_i$ : treatment effect<br>$\varepsilon_{ij}$ : residual error (error term)<br>$\gamma_W$ = within-study interaction term<br>$z_{ij}$ : participant-level covariate<br><br>Model should allow: |

| | |
|---|---|
| | - One intercept for each study (stratification)<br>- One treatment effect for each study (random effects)<br>- The separation of within-study and across-study interaction (by centering covariates and including their mean or proportion)<br>- One within-study interaction effect for each study (random effects)<br>- One residual variance for each study (stratification)<br>Uncorrelated random effects |
| Sensitivity analysis | A sensitivity analysis including the studies from which IPD are not available will be performed to explore the robustness of findings observed in the IPD meta-analysis. |
| Plot | Forest plot. |
| Level of evidence | Rated by GRADE; not performed by the statistician. |

##### 4.3.2 Continuation studies

Only randomised continuation trials will be included in the meta-analysis.

###### 4.3.2.1 Efficacy outcomes assessed at the end of the study

**Relapse**

| | |
|---|---|
| Type of outcome | Censored outcome, principal outcome for continuation trials. |
| Meta-analysis | Two-stage IPD meta-analysis for time-to-event outcome<br>- First stage:<br>o Methods: in line with each reanalysis. The participants will be analysed according to their allocated treatment at baseline (ITT principle) and not the treatment they receive at the time of outcome assessment.<br>o Results: hazard ratio and its variance.<br>- Second stage:<br>o Package meta, function metagen.<br>o Inverse variance method.<br>o Summary measure: hazard ratio.<br>o Random effects will be used in case of heterogeneity defined as an $I^2$ index > 25% and/or significant heterogeneity ($p < 0.10$) detected using Cochran's Q test statistic.<br>o In case of random effects, restricted maximum likelihood estimation (REML) estimation will be used and confidence interval will be derived by the Hartung-Knapp Sidik-Jonkman (HKSJ) approach. |
| Sensitivity analysis | A sensitivity analysis including the studies from which IPD are not available will be performed to explore the robustness of findings observed in the IPD meta-analysis. |
| Plot | Forest plot.<br>Funnel plot if applicable (> 10 studies). |
| Level of evidence | Rated by GRADE; not performed by the statistician. |

**Suicide/suicide attempt**

| | |
|---|---|
| Type of outcome | Binary outcome. |
| Meta-analysis | Two-stage IPD meta-analysis for binary outcome<br>- First stage: |

| | |
|---|---|
| | o Methods: in line with each reanalysis. The participants will be analysed according to their allocated treatment at baseline (ITT principle) and not the treatment they receive at the time of outcome assessment.<br>o Results: number of subjects with events, total number of subjects, per group.<br>- Second stage:<br>o Package meta, function metabin.<br>o Inverse variance method.<br>o Summary measure: relative risk.<br>o Random effects will be used in case of heterogeneity defined as an $I^2$ index > 25% and/or significant heterogeneity ($p < 0.10$) detected using Cochran's Q test statistic.<br>o In case of random effects, restricted maximum likelihood estimation (REML) estimation will be used and confidence interval will be derived by the Hartung-Knapp Sidik-Jonkman (HKSJ) approach. |
| Sensitivity analysis | A sensitivity analysis including the studies from which IPD are not available will be performed to explore the robustness of findings observed in the IPD meta-analysis. |
| Plot | Forest plot.<br>Funnel plot if applicable (> 10 studies). |
| Level of evidence | Rated by GRADE; not performed by the statistician. |

**Suicidal ideations, e.g. Beck Scale for Suicidal Ideation (BSS)**

| Type of outcome | Continuous outcome. |
|---|---|
| Meta-analysis | Two-stage IPD meta-analysis for continuous outcome<br>- First stage:<br>o Methods: in line with each reanalysis. The participants will be analysed according to their allocated treatment at baseline (ITT principle) and not the treatment they receive at the time of outcome assessment.<br>o Results: mean final score, standard deviation, number of subjects, per group.<br>- Second stage:<br>o Package meta, function metacont.<br>o Inverse variance method.<br>o Summary measure: mean difference.<br>o Random effects will be used in case of heterogeneity defined as an $I^2$ index > 25% and/or significant heterogeneity ($p < 0.10$) detected using Cochran's Q test statistic.<br>o In case of random effects, restricted maximum likelihood estimation (REML) estimation will be used and confidence interval will be derived by the Hartung-Knapp Sidik-Jonkman (HKSJ) approach. |
| Sensitivity analysis | A sensitivity analysis including the studies from which IPD are not available will be performed to explore the robustness of findings observed in the IPD meta-analysis. |
| Plot | Forest plot.<br>Funnel plot if applicable (> 10 studies). |
| Level of evidence | Rated by GRADE; not performed by the statistician. |

**MADRS**

| | |
|---|---|
| Type of outcome | Continuous outcome. |
| Meta-analysis | Two-stage IPD meta-analysis for continuous outcome<br>- First stage:<br>o Methods: in line with each reanalysis. The participants will be analysed according to their allocated treatment at baseline (ITT principle) and not the treatment they receive at the time of outcome assessment.<br>o Results: mean final score, standard deviation, number of subjects, per group.<br>- Second stage:<br>o Package meta, function metacont.<br>o Inverse variance method.<br>o Summary measure: mean difference.<br>o Random effects will be used in case of heterogeneity defined as an $I^2$ index > 25% and/or significant heterogeneity ($p < 0.10$) detected using Cochran's Q test statistic.<br>o In case of random effects, restricted maximum likelihood estimation (REML) estimation will be used and confidence interval will be derived by the Hartung-Knapp Sidik-Jonkman (HKSJ) approach.<br><br>Mean differences in MADRS observed (point estimates and confidence intervals) will be compared with 0 (absence of difference) and 6.5 points (the threshold defined *a priori* in initiation studies) on a forest plot (visual comparison). |
| Sensitivity analysis | A sensitivity analysis including the studies from which IPD are not available will be performed to explore the robustness of findings observed in the IPD meta-analysis. |
| Plot | Forest plot.<br>Funnel plot if applicable (> 10 studies). |
| Level of evidence | Rated by GRADE; not performed by the statistician. |

**Remission (score <8 if no binary)**

| | |
|---|---|
| Type of outcome | Binary outcome. |
| Meta-analysis | Two-stage IPD meta-analysis for binary outcome<br>- First stage:<br>o Methods: in line with each reanalysis. The participants will be analysed according to their allocated treatment at baseline (ITT principle) and not the treatment they receive at the time of outcome assessment.<br>o Results: number of subjects with events, total number of subjects, per group.<br>- Second stage:<br>o Package meta, function metabin.<br>o Inverse variance method.<br>o Summary measure: relative risk.<br>o Random effects will be used in case of heterogeneity defined as an $I^2$ index > 25% and/or significant heterogeneity ($p < 0.10$) detected using Cochran's Q test statistic. |

| | |
|---|---|
| | o In case of random effects, restricted maximum likelihood estimation (REML) estimation will be used and confidence interval will be derived by the Hartung-Knapp Sidik-Jonkman (HKSJ) approach. |
| Sensitivity analysis | A sensitivity analysis including the studies from which IPD are not available will be performed to explore the robustness of findings observed in the IPD meta-analysis. |
| Plot | Forest plot.<br>Funnel plot if applicable (> 10 studies). |
| Level of evidence | Rated by GRADE; not performed by the statistician. |

**Sheehan Disability Scale**

| Type of outcome | Continuous outcome. |
|---|---|
| Meta-analysis | Two-stage IPD meta-analysis for continuous outcome<br>- First stage:<br>o Methods: in line with each reanalysis. The participants will be analysed according to their allocated treatment at baseline (ITT principle) and not the treatment they receive at the time of outcome assessment.<br>o Results: mean final score, standard deviation, number of subjects, per group.<br>- Second stage:<br>o Package meta, function metacont.<br>o Inverse variance method.<br>o Summary measure: mean difference.<br>o Random effects will be used in case of heterogeneity defined as an $I^2$ index > 25% and/or significant heterogeneity ($p < 0.10$) detected using Cochran's Q test statistic.<br>o In case of random effects, restricted maximum likelihood estimation (REML) estimation will be used and confidence interval will be derived by the Hartung-Knapp Sidik-Jonkman (HKSJ) approach. |
| Sensitivity analysis | A sensitivity analysis including the studies from which IPD are not available will be performed to explore the robustness of findings observed in the IPD meta-analysis. |
| Plot | Forest plot.<br>Funnel plot if applicable (> 10 studies). |
| Level of evidence | Rated by GRADE; not performed by the statistician. |

**PHQ-9**

| Type of outcome | Continuous outcome. |
|---|---|
| Meta-analysis | Two-stage IPD meta-analysis for continuous outcome<br>- First stage:<br>o Methods: in line with each reanalysis. The participants will be analysed according to their allocated treatment at baseline (ITT principle) and not the treatment they receive at the time of outcome assessment.<br>o Results: mean final score, standard deviation, number of subjects, per group.<br>- Second stage:<br>o Package meta, function metacont. |

| | |
|---|---|
| | ○ Inverse variance method.<br>○ Summary measure: mean difference.<br>○ Random effects will be used in case of heterogeneity defined as an $I^2$ index > 25% and/or significant heterogeneity ($p < 0.10$) detected using Cochran's Q test statistic.<br>○ In case of random effects, restricted maximum likelihood estimation (REML) estimation will be used and confidence interval will be derived by the Hartung-Knapp Sidik-Jonkman (HKSJ) approach. |
| Sensitivity analysis | A sensitivity analysis including the studies from which IPD are not available will be performed to explore the robustness of findings observed in the IPD meta-analysis. |
| Plot | Forest plot.<br>Funnel plot if applicable (> 10 studies). |
| Level of evidence | Rated by GRADE; not performed by the statistician. |

###### 4.3.2.2 Safety outcomes assessed at the end of the study

**Serious adverse events**

| | |
|---|---|
| Type of outcome | Count outcome, i.e. number of events per participant. |
| Meta-analysis | Two-stage IPD meta-analysis for count outcome<br>- First stage:<br>○ Methods: in line with each reanalysis. The participants will be analysed according to the treatment they receive at the time of outcome assessment.<br>○ Results: number of events, person time at risk, per group.<br>- Second stage:<br>○ Package meta, function metainc.<br>○ Inverse variance method.<br>○ Summary measure: incidence rate ratio.<br>○ Random effects will be used in case of heterogeneity defined as an $I^2$ index > 25% and/or significant heterogeneity ($p < 0.10$) detected using Cochran's Q test statistic.<br>○ In case of random effects, restricted maximum likelihood estimation (REML) estimation will be used and confidence interval will be derived by the Hartung-Knapp Sidik-Jonkman (HKSJ) approach. |
| Sensitivity analysis | A sensitivity analysis including the studies from which IPD are not available will be performed to explore the robustness of findings observed in the IPD meta-analysis. |
| Plot | Forest plot.<br>Funnel plot if applicable (> 10 studies). |
| Level of evidence | Rated by GRADE; not performed by the statistician. |

**Dropout for any cause**

| | |
|---|---|
| Type of outcome | Binary outcome. |
| Meta-analysis | Two-stage IPD meta-analysis for binary outcome<br>- First stage:<br>○ Methods: in line with each reanalysis. The participants will be analysed according to the treatment they receive at the time of outcome assessment. |

| | |
|---|---|
| | ○ Results: number of subjects with events, total number of subjects, per group.<br>- Second stage:<br>○ Package meta, function metabin.<br>○ Inverse variance method.<br>○ Summary measure: relative risk.<br>○ Random effects will be used in case of heterogeneity defined as an $I^2$ index > 25% and/or significant heterogeneity ($p < 0.10$) detected using Cochran's Q test statistic.<br>○ In case of random effects, restricted maximum likelihood estimation (REML) estimation will be used and confidence interval will be derived by the Hartung-Knapp Sidik-Jonkman (HKSJ) approach. |
| Sensitivity analysis | A sensitivity analysis including the studies from which IPD are not available will be performed to explore the robustness of findings observed in the IPD meta-analysis. |
| Plot | Forest plot.<br>Funnel plot if applicable (> 10 studies). |
| Level of evidence | Rated by GRADE; not performed by the statistician. |

**Dropout due to adverse events**

| | |
|---|---|
| Type of outcome | Binary outcome. |
| Meta-analysis | Two-stage IPD meta-analysis for binary outcome<br>- First stage:<br>○ Methods: in line with each reanalysis. The participants will be analysed according to the treatment they receive at the time of outcome assessment.<br>○ Results: number of subjects with events, total number of subjects, per group.<br>- Second stage:<br>○ Package meta, function metabin.<br>○ Inverse variance method.<br>○ Summary measure: relative risk.<br>○ Random effects will be used in case of heterogeneity defined as an $I^2$ index > 25% and/or significant heterogeneity ($p < 0.10$) detected using Cochran's Q test statistic.<br>○ In case of random effects, restricted maximum likelihood estimation (REML) estimation will be used and confidence interval will be derived by the Hartung-Knapp Sidik-Jonkman (HKSJ) approach. |
| Sensitivity analysis | A sensitivity analysis including the studies from which IPD are not available will be performed to explore the robustness of findings observed in the IPD meta-analysis. |
| Plot | Forest plot.<br>Funnel plot if applicable (> 10 studies). |
| Level of evidence | Rated by GRADE; not performed by the statistician. |

**Adverse events**

| | |
|---|---|
| Type of outcome | Count outcome, i.e. number of events per participant. |
| Meta-analysis | Two-stage IPD meta-analysis for count outcome<br>- First stage: |

| | |
|---|---|
| | ○ Methods: in line with each reanalysis. The participants will be analysed according to the treatment they receive at the time of outcome assessment.<br>○ Results: number of events, person time at risk, per group.<br>- Second stage:<br>○ Package meta, function metainc.<br>○ Inverse variance method.<br>○ Summary measure: incidence rate ratio.<br>○ Random effects will be used in case of heterogeneity defined as an $I^2$ index > 25% and/or significant heterogeneity ($p < 0.10$) detected using Cochran's Q test statistic.<br>○ In case of random effects, restricted maximum likelihood estimation (REML) estimation will be used and confidence interval will be derived by the Hartung-Knapp Sidik-Jonkman (HKSJ) approach. |
| Sensitivity analysis | A sensitivity analysis including the studies from which IPD are not available will be performed to explore the robustness of findings observed in the IPD meta-analysis. |
| Plot | Forest plot.<br>Funnel plot if applicable (> 10 studies). |
| Level of evidence | Rated by GRADE; not performed by the statistician. |

**Blood pressure**

| | |
|---|---|
| Type of outcome | Continuous outcome. |
| Meta-analysis | Two-stage IPD meta-analysis for continuous outcome<br>- First stage:<br>○ Methods: in line with each reanalysis. The participants will be analysed according to the treatment they receive at the time of outcome assessment.<br>○ Results: mean final score, standard deviation, number of subjects, per group.<br>- Second stage:<br>○ Package meta, function metacont.<br>○ Inverse variance method.<br>○ Summary measure: mean difference.<br>○ Random effects will be used in case of heterogeneity defined as an $I^2$ index > 25% and/or significant heterogeneity ($p < 0.10$) detected using Cochran's Q test statistic.<br>○ In case of random effects, restricted maximum likelihood estimation (REML) estimation will be used and confidence interval will be derived by the Hartung-Knapp Sidik-Jonkman (HKSJ) approach. |
| Sensitivity analysis | A sensitivity analysis including the studies from which IPD are not available will be performed to explore the robustness of findings observed in the IPD meta-analysis. |
| Plot | Forest plot.<br>Funnel plot if applicable (> 10 studies). |
| Level of evidence | Rated by GRADE; not performed by the statistician. |

**Dissociation**

| | |
|---|---|
| Type of outcome | Binary outcome. |

| | |
|---|---|
| Meta-analysis | In case of dissociation is not explicitly reported, the AEs will be assessed in a blind manner (i.e. without knowledge of the treatment group) and discussed by a clinician.<br><br>Two-stage IPD meta-analysis for binary outcome<br>- First stage:<br>o Methods: in line with each reanalysis. The participants will be analysed according to the treatment they receive at the time of outcome assessment.<br>o Results: number of subjects with events, total number of subjects, per group.<br>- Second stage:<br>o Package meta, function metabin.<br>o Inverse variance method.<br>o Summary measure: relative risk.<br>o Random effects will be used in case of heterogeneity defined as an $I^2$ index > 25% and/or significant heterogeneity ($p < 0.10$) detected using Cochran's Q test statistic.<br>o In case of random effects, restricted maximum likelihood estimation (REML) estimation will be used and confidence interval will be derived by the Hartung-Knapp Sidik-Jonkman (HKSJ) approach. |
| Sensitivity analysis | A sensitivity analysis including the studies from which IPD are not available will be performed to explore the robustness of findings observed in the IPD meta-analysis. |
| Plot | Forest plot.<br>Funnel plot if applicable (> 10 studies). |
| Level of evidence | Rated by GRADE; not performed by the statistician. |

**Sedation**

| | |
|---|---|
| Type of outcome | Binary outcome. |
| Meta-analysis | In case of sedation is not explicitly reported, the AEs will be assessed in a blind manner (i.e. without knowledge of the treatment group) and discussed by a clinician.<br><br>Two-stage IPD meta-analysis for binary outcome<br>- First stage:<br>o Methods: in line with each reanalysis. The participants will be analysed according to the treatment they receive at the time of outcome assessment.<br>o Results: number of subjects with events, total number of subjects, per group.<br>- Second stage:<br>o Package meta, function metabin.<br>o Inverse variance method.<br>o Summary measure: relative risk.<br>o Random effects will be used in case of heterogeneity defined as an $I^2$ index > 25% and/or significant heterogeneity ($p < 0.10$) detected using Cochran's Q test statistic.<br>o In case of random effects, restricted maximum likelihood estimation (REML) estimation will be used and confidence |

| | |
|---|---|
| | interval will be derived by the Hartung-Knapp Sidik-Jonkman (HKSJ) approach. |
| Sensitivity analysis | A sensitivity analysis including the studies from which IPD are not available will be performed to explore the robustness of findings observed in the IPD meta-analysis. |
| Plot | Forest plot.<br>Funnel plot if applicable (> 10 studies). |
| Level of evidence | Rated by GRADE; not performed by the statistician. |

###### 4.3.2.3 Moderators of esketamine efficacy

Based on the individual data, the interactions will be assessed first with a two-stage approach. One-stage IPD meta-analysis may be used if necessary. The use of two-stage approach is a change from the initial protocol that was made to handle the risk for ecological bias inherent with the one-stage approach.

**Impact of resistance stage**

Each participant will be classified according to Thase and Rush resistance stages in accordance with their baseline characteristics. In case of missing data, the possibility to classify participants will be explored in a blind manner (i.e. without knowledge of the treatment group) with a clinician.

| | |
|---|---|
| Two-stage IPD meta-analysis | First stage:<br>- Outcome: MADRS at the end of the study (continuous outcome).<br>- Covariate: resistance stage (quantitative outcome, 1, 2, 3, 4, 5).<br>- Estimation of the interaction term, between resistance stage and treatment group, and its variance in each study, in line with each reanalysis.<br>- The participants will be analysed according to their allocated treatment at baseline (ITT principle) and not the treatment they receive at the time of outcome assessment.<br>- Results: interaction term, standard deviation, number of subjects, per group.<br>Second stage:<br>- Outcome: interaction term (continuous outcome).<br>- Package meta, function metagen.<br>- Inverse variance method.<br>- Summary measure: interaction effect.<br>- Random effects will be used in case of heterogeneity defined as an $I^2$ index > 25% and/or significant heterogeneity ($p < 0.10$) detected using Cochran's Q test statistic.<br>- In case of random effects, restricted maximum likelihood estimation (REML) estimation will be used and confidence interval will be derived by the Hartung-Knapp Sidik-Jonkman (HKSJ) approach. |
| One-stage IPD meta-analysis | Outcome: MADRS at the end of the study (continuous outcome).<br>$MADRS_{ij} = \phi_i + \beta 1_i z_{ij} + \theta_i treat_{ij} + \gamma_W (treat_{ij} * (z_{ij}-\bar{z}_i)) + \varepsilon_{ij} \quad \varepsilon_{ij} \sim N(0, \sigma_i^2)$<br>i = 1 to k trials<br>j participants<br>Esketamine group ($treat_{ij} = 1$), control group ($treat_{ij} = 0$)<br>$MADRS_{ij}$ : outcome<br>$\phi_i$ : control effect (intercept term) |

| | |
|---|---|
| | $\theta_i$ : treatment effect<br>$\varepsilon_{ij}$ : residual error (error term)<br>$\gamma_W$ = within-study interaction term<br>$z_{ij}$ : participant-level covariate<br><br>Model should allow:<br>- One intercept for each study (stratification)<br>- One treatment effect for each study (random effects)<br>- The separation of within-study and across-study interaction (by centering covariates and including their mean or proportion)<br>- One within-study interaction effect for each study (random effects)<br>- One residual variance for each study (stratification)<br>- Uncorrelated random effects |
| Sensitivity analysis | A sensitivity analysis including the studies from which IPD are not available will be performed to explore the robustness of findings observed in the IPD meta-analysis. |
| Plot | Forest plot. |
| Level of evidence | Rated by GRADE; not performed by the statistician. |

**Impact of age**

| | |
|---|---|
| Two-stage IPD meta-analysis | First stage:<br>- Outcome: MADRS at the end of the study (continuous outcome).<br>- Covariate: age (continuous outcome).<br>- Estimation of the interaction term, between age and treatment group, and its variance in each study, in line with each reanalysis.<br>- The participants will be analysed according to their allocated treatment at baseline (ITT principle) and not the treatment they receive at the time of outcome assessment.<br>- Results: interaction term, standard deviation, number of subjects, per group.<br>Second stage:<br>- Outcome: interaction term (continuous outcome).<br>- Package meta, function metagen.<br>- Inverse variance method.<br>- Summary measure: interaction effect.<br>- Random effects will be used in case of heterogeneity defined as an $I^2$ index > 25% and/or significant heterogeneity ($p < 0.10$) detected using Cochran's Q test statistic.<br>- In case of random effects, restricted maximum likelihood estimation (REML) estimation will be used and confidence interval will be derived by the Hartung-Knapp Sidik-Jonkman (HKSJ) approach. |
| One-stage IPD meta-analysis | Outcome: MADRS at the end of the study (continuous outcome).<br>$MADRS_{ij} = \phi_i + \beta 1_i z_{ij} + \theta_i treat_{ij} + \gamma_W (treat_{ij} * (z_{ij} - \bar{z}_i)) + \varepsilon_{ij} \quad \varepsilon_{ij} \sim N(0, \sigma_i^2)$<br>i = 1 to k trials<br>j participants<br>Esketamine group ($treat_{ij}$ = 1), control group ($treat_{ij}$ = 0)<br>$MADRS_{ij}$ : outcome<br>$\phi_i$ : control effect (intercept term)<br>$\theta_i$ : treatment effect<br>$\varepsilon_{ij}$ : residual error (error term)<br>$\gamma_W$ = within-study interaction term |

| | |
|---|---|
| | $z_{ij}$ : participant-level covariate<br><br>Model should allow:<br>- One intercept for each study (stratification)<br>- One treatment effect for each study (random effects)<br>- The separation of within-study and across-study interaction (by centering covariates and including their mean or proportion)<br>- One within-study interaction effect for each study (random effects)<br>- One residual variance for each study (stratification)<br>Uncorrelated random effects |
| Sensitivity analysis | A sensitivity analysis including the studies from which IPD are not available will be performed to explore the robustness of findings observed in the IPD meta-analysis. |
| Plot | Forest plot. |
| Level of evidence | Rated by GRADE; not performed by the statistician. |

**Impact of per capita gross national income**

Another sensitivity analysis will be performed to explore if efficacy varies according to the World Bank categorization into low, middle, and high income (https://data.worldbank.org/country) as initial evidence on antidepressants suggests that per capita gross national income is associated with trial results.

| Study | High income country | Low, lower middle, and upper middle income country | Missing data |
|---|---|---|---|
| 1 | *Number of participants* | *Number of participants* | *Number of participants* |
| 2 | *Number of participants* | *Number of participants* | *Number of participants* |
| 3 | *Number of participants* | *Number of participants* | *Number of participants* |
| … | *Number of participants* | *Number of participants* | *Number of participants* |

*Table 11: Description of the per capita gross national income in each continuation study*

| | |
|---|---|
| Two-stage IPD meta-analysis | First stage:<br>- Outcome: MADRS at the end of the study (continuous outcome).<br>- Covariate: per capita gross national income (binary outcome: high income countries vs. the other).<br>- Estimation of the interaction term, between per capita gross national income and treatment group, and its variance in each study, in line with each reanalysis.<br>- The participants will be analysed according to their allocated treatment at baseline (ITT principle) and not the treatment they receive at the time of outcome assessment.<br>- Results: interaction term, standard deviation, number of subjects, per group.<br>Second stage:<br>- Outcome: interaction term (continuous outcome).<br>- Package meta, function metagen.<br>- Inverse variance method.<br>- Summary measure: interaction effect.<br>- Random effects will be used in case of heterogeneity defined as an $I^2$ index > 25% and/or significant heterogeneity ($p < 0.10$) detected using Cochran's Q test statistic. |

| | |
|---|---|
| | - In case of random effects, restricted maximum likelihood estimation (REML) estimation will be used and confidence interval will be derived by the Hartung-Knapp Sidik-Jonkman (HKSJ) approach. |
| One-stage IPD meta-analysis | Outcome: MADRS at the end of the study (continuous outcome).<br>$MADRS_{ij} = \phi_i + \beta1_i\, z_{ij} + \theta_i\, treat_{ij} + \gamma_W\, (treat_{ij} * (z_{ij}-\bar{z}_i)) + \varepsilon_{ij} \quad \varepsilon_{ij} \sim N(0, \sigma_i^2)$<br>i = 1 to k trials<br>j participants<br>Esketamine group ($treat_{ij}$ = 1), control group ($treat_{ij}$ = 0)<br>$MADRS_{ij}$ : outcome<br>$\phi_i$ : control effect (intercept term)<br>$\theta_i$ : treatment effect<br>$\varepsilon_{ij}$ : residual error (error term)<br>$\gamma_W$ = within-study interaction term<br>$z_{ij}$ : participant-level covariate<br><br>Model should allow:<br>- One intercept for each study (stratification)<br>- One treatment effect for each study (random effects)<br>- The separation of within-study and across-study interaction (by centering covariates and including their mean or proportion)<br>- One within-study interaction effect for each study (random effects)<br>- One residual variance for each study (stratification)<br>Uncorrelated random effects |
| Sensitivity analysis | A sensitivity analysis including the studies from which IPD are not available will be performed to explore the robustness of findings observed in the IPD meta-analysis. |
| Plot | Forest plot. |
| Level of evidence | Rated by GRADE; not performed by the statistician. |
