## Supplementary material for "Efficacy and safety of esketamine for “treatment resistant depression”: registered report for a Systematic Review with an Individual Patient Data Meta-analysis of Randomized, Double-Blind, Placebo-Controlled Trials": Web appendix 3

**Statistical report (SR)**

| **Full study title** | Efficacy and safety of esketamine for “treatment resistant depression”: registered report for a Systematic Review with an Individual Participant Data Meta-analysis of Randomized, Double-Blind, Placebo-Controlled Trials |
| --- | --- |
| **Acronym** | ESK-T-DEP |
| **Prospero registry number** | CRD42021290721 |
| **OSF registration** | https://osf.io/xetvn |
| **Study protocol version** | V1.0 |
| **SR version** | V2 |

**Roles and responsibility**

| Author | Estelle Le Pabic |
| --- | --- |
| Coordinator | Florian NAUDET |

**SR revision history**

| Date | Document | Version | Description of and reason for change |
| --- | --- | --- | --- |
| 08.08.2024 | Statistical_report_240808_V1 | V1 | Creation of the document |
| 21.03.2024 | Statistical_report_250321_V1 | V2 | Additional analysis |

Table des matières

### 1 Methods

The main objective is to independently reappraise the efficacy of esketamine in “treatment resistant depression” (TRD) using an individual participant data meta-analysis methodology. The secondary objectives are to reappraise the safety of esketamine in the treatment of TRD and to explore moderating factors of esketamine efficacy and safety, including level of treatment resistance, patient age, site-specific effects, among others.

#### 1.1 Outcomes

The primary outcome will be the MADRS score (continuous outcome) assessed after at least 4 weeks (the end of treatment in the pivotal initiation trials / it is expected to be 4 weeks but may be longer depending on the studies selected for the meta-analysis). To take into account the possible rapid effect of esketamine, we will also study the MADRS score at week 1 as a secondary outcome.

All of the following secondary outcomes will be assessed at end of treatment as well as at the last follow-up visit post-treatment (expected to be 24 weeks) in initiation trials. These will be assessed at the end of the study in continuation trial (this is expected to be over 500 days).

Secondary outcomes will include efficacy outcomes, namely (a) suicide/suicide attempt (binary outcome); b) suicidal ideations, e.g. Beck Scale for Suicidal Ideation (BSS, continuous outcome); c) MADRS (continuous outcome, similar to the primary outcome but, here using the last follow-up visit post-treatment); d) remission (binary outcome); e) Sheehan Disability Scale (continuous outcome); f) PHQ-9 (continuous outcome); g) relapse (censored outcome) for continuation trials, and safety outcomes, namely a) serious adverse events (count outcome, i.e. number of events per patient); b) dropout for any cause (binary outcome); c) dropout due to adverse events (binary outcome); d) Adverse events (count outcome i.e. number of events per patient); e) Blood pressure (continuous repeatedly measured outcome); f) Dissociation (binary outcome) g) Sedation (binary outcome).

#### 1.2 Study participants

We were consider studies that include all patients with TRD meeting the inclusion criteria of esketamine’s development program for this indication. Following the definition used in this program, TRD refers to a depressive episode with inadequate response to at least two antidepressant trials of adequate doses and duration). This definition includes “higher” level of “resistance” according to Thase and Rush staging method of treatment resistant depression [20]. There was no age limit. In terms of strategy the systematic review search criteria was include all studies of esketamine for the primary treatment of depression. The IPD analysis was then be restricted to participants/patients with TRD within the included studies (i.e., have failed > 2 adequate antidepressant trials).

Included studies were studies that compare the use of intranasal esketamine (with the following approved doses: 56 mg or 84 mg or “flexible” doses) to a placebo. In the trials, placebo is described as a solution with a bittering agent added to simulate the taste of the esketamine solution. All randomised controlled trials (initiation and continuation trials) will be considered.

#### 1.3 Availability of data and materials

The datasets that analyzed during the current study are available in the YODA repository (<https://yoda.yale.edu/>). All prespecified analysis plans and the code to reproduce our analyses were shared on the [Open Science Framework](https://osf.io/).

This study, carried out under YODA Project 2021-4851, used data obtained from the Yale University Open Data Access Project, which has an agreement with JANSSEN RESEARCH & DEVELOPMENT, L.L.C.. The interpretation and reporting of research using this data are solely the responsibility of the authors and does not necessarily represent the official views of the Yale University Open Data Access Project or JANSSEN RESEARCH & DEVELOPMENT, L.L.C..

#### 1.4 Selected studies

Thirteen studies were selected. After applying the inclusion criteria, 6 initiation studies (NCT01998958, NCT02918318, NCT02417064, NCT02418585, NCT02422186 and NCT03434041) and 1 continuation study (NCT02493868). For study NCT01998958, only data at 1 week were selected, because patients were randomized after 1 week of treatment. The study were divided in two panel, A and B.

#### 1.5 Analysis strategy

For efficacy and safety outcomes a two-step approach will be adopted, the one-stage approach will be used to explore moderators of esketamine efficacy. All the analyses will be performed with R (R core Team, 2021).

A fixed effect meta-analysis, or a random effect meta-analysis (in case of heterogeneity defined as an I^2^ index > 25 % and/or detection of significant heterogeneity (p<0.10) detected using Cochran's Q test statistic), has been used to pool the different efficacy indexes (mean differences or relative risks or odds ratio or hazard ratios) that will be derived from these re-analyses. Initiation and continuation designs were analyzed separately. For the primary outcome, mean differences in MADRS scores observed (point estimates and confidence intervals were compared with 0 (absence of difference) and 6.5 points (the threshold defined a priori in initiation studies).

Sensitivity analyses were performed to deal with missing data. The multiple imputation by chained equations (MICE) method implemented in the mice package in R was used under the assumption of missing at random data and m=10 imputation were generated.

For the analysis of blood pressure, the one-satge approach was used with the realsiation of a linear mixed-effect model with treatment and measurement interaction (predose and at 40 minutes, 1 hour and 1.5 hours postdose) and treatment group and day effect as random effect and stratified by trial the intercept.

To explore moderators of esketamine efficay in one-stage approach, a mixed model or was specified with one intercept for each study (random effects), one treatment effect for each study, the separation of within-study and across-study interaction, one within-study interaction effect for each study (random effects) and one residual variance for each study (stratification).

### 2 Study description

Table 1: Studies included

| **Study** | **Initiation or continuation study** | **Reanalysis OSF link** |
| --- | --- | --- |
| NCT01998958 (2003, panel A) | Initiation study | [OSF_ ReAnalysis_2003.docx](https://osf.io/67ynq) |
| NCT01998958 (2003, panel B) | Initiation study | [OSF_ReAnalysis_2003.docx](https://osf.io/67ynq) |
| NCT02918318 (2005) | Initiation study | [OSF_ReAnalysis_2005.docx](https://osf.io/hg3x6) |
| NCT02417064 (3001) | Initiation study | [OSF_ReAnalysis_3001.docx](https://osf.io/skb42) |
| NCT02418585 (3002) | Initiation study | [OSF_ReAnalysis_3002xpt.docx](https://osf.io/z6uwc) |
| NCT02422186 (3005) | Initiation study | [OSF_ReAnalysis_3005.docx](https://osf.io/zw3dv) |
| NCT03434041 (3006) | Initiation study | [OSF_ReAnalysis_3006.docx](https://osf.io/zgu9t) |
| NCT02493868 (3003) | Continuation study | [OSF_ReAnalysis3003.docx](https://osf.io/zdfe7) |

Table 2: Studies assessed at the end of treatment

| **Study** | **Total** | **Esketamine** | **Placebo** |
| --- | --- | --- | --- |
| NCT02918318 (2005) | 161 | 81 | 80 |
| NCT02417064 (3001) | 342 | 229 | 113 |
| NCT02418585 (3002) | 232 | 118 | 114 |
| NCT02422186 (3005) | 138 | 72 | 66 |
| NCT03434041 (3006) | 250 | 124 | 126 |
| **Overall population** | **1123** | **624** | **499** |

Table 3: Studies assessed at 1 week

| **Study** | **Total** | **Esketamine** | **Placebo** |
| --- | --- | --- | --- |
| NCT01998958 (2003, panel A) | 55 | 23 | 32 |
| NCT01998958 (2003, panel B) | 30 | 9 | 21 |
| NCT02918318 (2005) | 161 | 81 | 80 |
| NCT02417064 (3001) | 342 | 229 | 113 |
| NCT02418585 (3002) | 232 | 118 | 114 |
| NCT02422186 (3005) | 138 | 72 | 66 |
| NCT03434041 (3006) | 250 | 124 | 126 |
| **Overall population** | **1208** | **656** | **552** |

Table 4: Studies assessed at the follow-up visit

| **Study** | **Total** | **Esketamine** | **Placebo** |
| --- | --- | --- | --- |
| NCT02918318 (2005) | 161 | 81 | 80 |
| NCT03434041 (3006) | 250 | 124 | 126 |
| **Overall population** | **411** | **205** | **206** |

Table 5: Treatment during follow-up phase

| **Study** | **Total** |
| --- | --- |
| NCT02918318 (2005) | No intranasal study medication were administered during follow-up phase. The oral antidepressant medication were to be continued in this phase unless determined as not clinically appropriate |
| NCT03434041 (3006) | No intranasal study medication were administered during follow-up phase. The decision to continue the oral antidepressant in this phase were at the discretion of the investigator. |

The total duration of a posttreatment follow-up (FU) phase in study NCT03434041 (3006) was 8-week. For study NCT02918318 (2005), responders (subject who have ≥50% reduction from baseline in MADRS total score) at the end of the double-blind (DB) induction phase were eligible to proceed to the posttreatment phase; thode who do bot (ie, nonresponders) were proceed to the 4-week follow-up phase, the 8-week posttreatment were selected for to match 4-week follow-up (4-week DB + 4-week posttreatment).

Table 6: Continuation study

| **Study** | **Total** | **Esketamine** | **Placebo** |
| --- | --- | --- | --- |
| NCT02493868 (3003) | 297 | 152 | 145 |

Table 7: Esketamine dose and study duration (double-blinded)

| **Study** | **Esketamine dose*** | **Study duration (blinded)** |
| --- | --- | --- |
| NCT01998958 (2003, panel A) | 28, 56 and 84 mg | 8 days |
| NCT01998958 (2003, panel B) | 14, 56 mg | 8 days |
| NCT02918318 (2005) | 28, 56 and 84 mg | 28 days |
| NCT02417064 (3001) | 56 and 84 mg | 28 days |
| NCT02418585 (3002) | Flexible (56 or 84 mg) | 28 days |
| NCT02422186 (3005) | Flexible (28, 56 or 84 mg) | 28 days |
| NCT03434041 (3006) | Flexible (56 or 84 mg) | 28 days |
| NCT02493868 (3003 : continuation study) | Flexible (56 or 84 mg) | Time to event |

* For the re-analyses, all doses were considered; however, for the meta-analyses, only doses of 56 mg, 84 mg, or flexible dosing were included.

Table 8: Description patient (mean age and sex) and major deviations in double-blinded phase

| **Study** | **Mean age ± standard deviation** | **Sex, n (%)** |
| --- | --- | --- |
| NCT01998958 (2003, panel A) | 42.9 ± 10.1 | M: 29 (43.3) ; W: 38 (56.7) |
| NCT01998958 (2003, panel B) | 42.2 ± 8.0 | M: 24 (58.5) ; W: 17 (41.5) |
| NCT02918318 (2005) | 43.4 ± 10.4 | M: 106 (52.5) ; W: 96 (47.5) |
| NCT02417064 (3001) | 46.7 ± 11.2 | M: 101 (29.5) ; W: 241 (70.5) |
| NCT02418585 (3002) | 46.2 ± 11.8 | M: 90 (38.8) ; W: 142 (61.2) |
| NCT02422186 (3005) | 70.3 ± 4.5 | M: 52 (37.7) ; W: 86 (62.3 |
| NCT03434041 (3006) | 36.4 ± 12.4 | M: 137 (54.8) ; W: 113 (45.2) |
| NCT02493868 (3003 : continuation study) : Responders | 47.9 ± 10.5 | M: 42 (34.7) ; W: 79 (65.3) |
| NCT02493868 (3003 : continuation study) : Remiters | 46.9 ± 11.6 | M: 58 (33) ; W: 118 (67) |

M: Male; W: Women

Table 9: Major deviations

| **Study** | **Details of observed major deviations** |
| --- | --- |
| NCT01998958 (2003, panel A) | There were 5 major deviations: 1 patient entered but did not satisfy criteria and 4 patients had major deviations categorized as 'other,' with no further details provided. |
| NCT01998958 (2003, panel B) | There were 2 major deviations: 2 patients had major deviations categorized as 'other,' with no further details provided. |
| NCT02918318 (2005) | There were 25 major deviations: 5 patients received disallowed concomitant treatments, 10 patients received the wrong treatment or an incorrect dose, and 9 patients had deviations categorized as 'other' (including 1 patient with 2 deviations). No further details were provided. |
| NCT02417064 (3001) | There were 62 major deviations: 28 patients entered but did not satisfy criteria (of those, one had 2 deviations), 9 patients received disallowed concomitant treatments, 1 patient received wrong treatment or incorrect dose, 2 patients developed withdrawal criteria but not withdrawn, and 21 patients had deviations categorized as 'other'. No further details were provided. |
| NCT02418585 (3002) | There were 14 major deviations: 3 patients received disallowed concomitant treatments, 3 patients received the wrong treatment or an incorrect dose and 8 patients had deviations categorized as 'other'. No further details were provided. |
| NCT02422186 (3005) | There were 22 major deviations: 6 patients received the wrong treatment or an incorrect dose, 6 entered but did not satisfy criteria and 10 had deviations categorized as 'other'. No further details were provided. |
| NCT03434041 (3006) | There were 21 major deviations: 10 patients received disallowed concomitant treatments, 2 patients received the wrong treatment or an incorrect dose, 2 patients entered but did not satisfy criteria and 7 patients had deviations categorized as 'other'. No further details were provided. |
| NCT02493868 (3003 : continuation study) | There were 5 major deviations: 2 patients developed withdrawal criteria but not withdrawn, 1 patient received the wrong treatment or an incorrect dose, 1 patient received disallowed concomitant treatments and 1 patient had deviations categorized as 'other'. No further details were provided. |

### 3 Results

#### 3.1 Initiation studies

##### 3.1.1 Efficacy outcomes

###### 3.1.1.1 Meta-analysis of primary outcome

Table 10: Missing data per study for primary outcome

| **Study** | **Total** | **Esketamine** | **Placebo** |
| --- | --- | --- | --- |
| NCT02918318 (2005) | 16* | 8 | 8 |
| NCT02417064 (3001) | 24 | 19 | 5 |
| NCT02418585 (3002) | 21 | 13 | 8 |
| NCT02422186 (3005) | 15 | 9 | 6 |
| NCT03434041 (3006) | 35 | 15 | 19 |

*The 41 patients in the esketamine 28mg group were not included in the analysis and are not included in the calculation of the number of missing data.

**MADRS (Montgomery-Asberg Depression Rating Scale) after at least 4 weeks**

Figure 1: Forest plot, MADRS after at least 4 weeks outcome

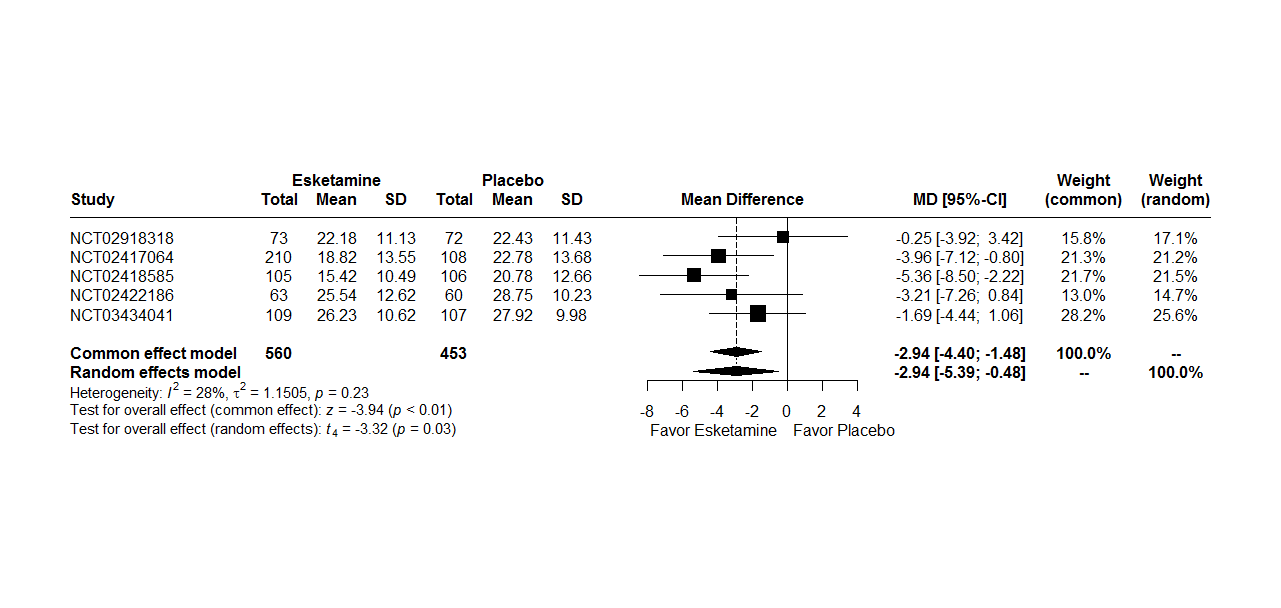

*MD : Mean Difference*

*CI : Confidence Interval*

A random effects model were used because there are heterogeneity between studies (I^2^ = 28%, t^2^ = 1.15, p = 0.23). The two-stage meta-analysis indicates a significant effect between MADRS at 4 weeks and treatment group with a mean difference of -2.94 [-4.40; -1.48], p < 0.01 in common effect model and a mean difference of -2.94 [-5.39; -0.48], p = 0.03 in a random effects model.

Figure 2: Forest plot, MADRS after at least 4 weeks outcome and line reference at -6.5 (6.5 points is the threshold defined a priori in initiation studies)

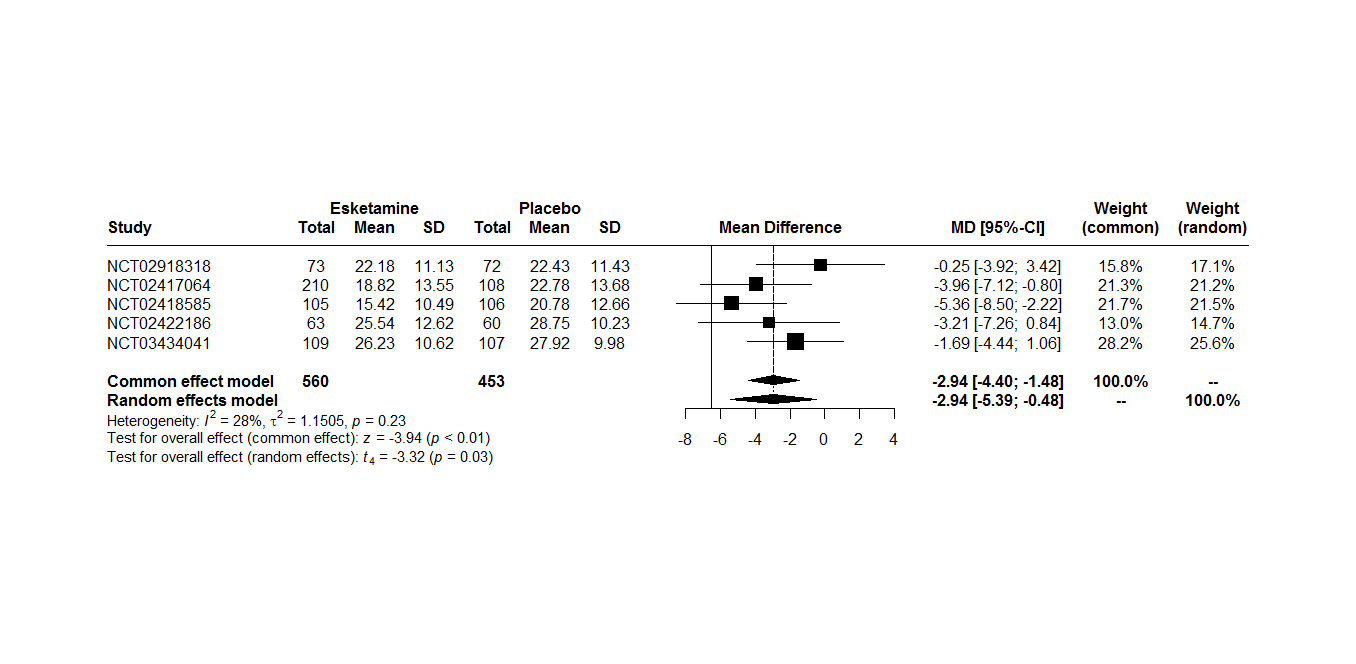

*MD : Mean Difference*

*CI : Confidence Interval*

**MADRS after at least 4 weeks with imputation missing data**

Figure 3: Forest plot, MADRS after at least 4 weeks outcome with imputation missing data

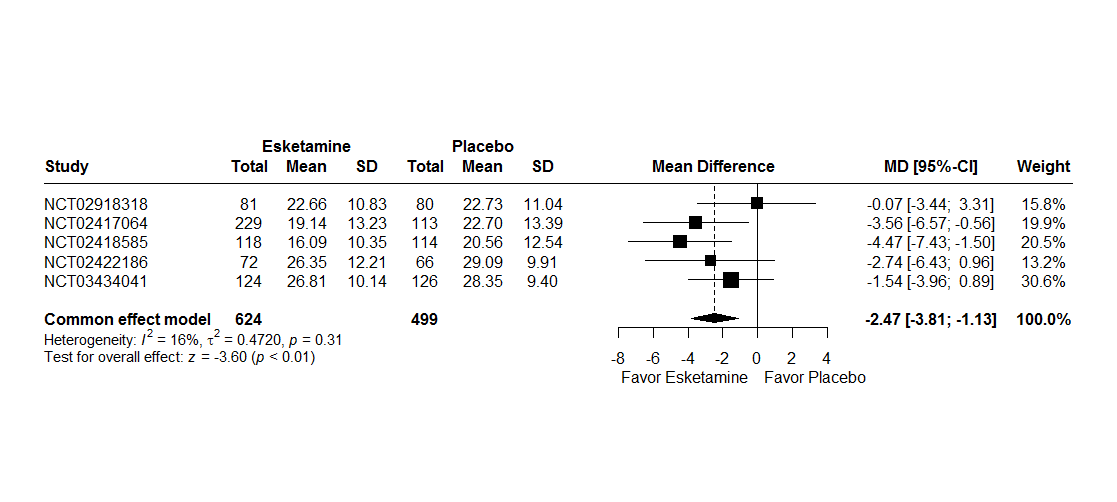

*MD : Mean Difference*

*CI : Confidence Interval*

A random effects model were not used because there are no heterogeneity between studies (I^2^ = 16%, t^2^ = 0.47, p = 0.31). The two-stage meta-analysis indicates a significant effect between MADRS at 4 weeks with imputation data and treatment group with a beta of -2.47 [-3.81; -1.13], p < 0.01 in common effect model.

Figure 4: Forest plot, MADRS after at least 4 weeks outcome with imputation missing data and line reference at -6.5 (6.5 points is the threshold defined a priori in initiation studies)

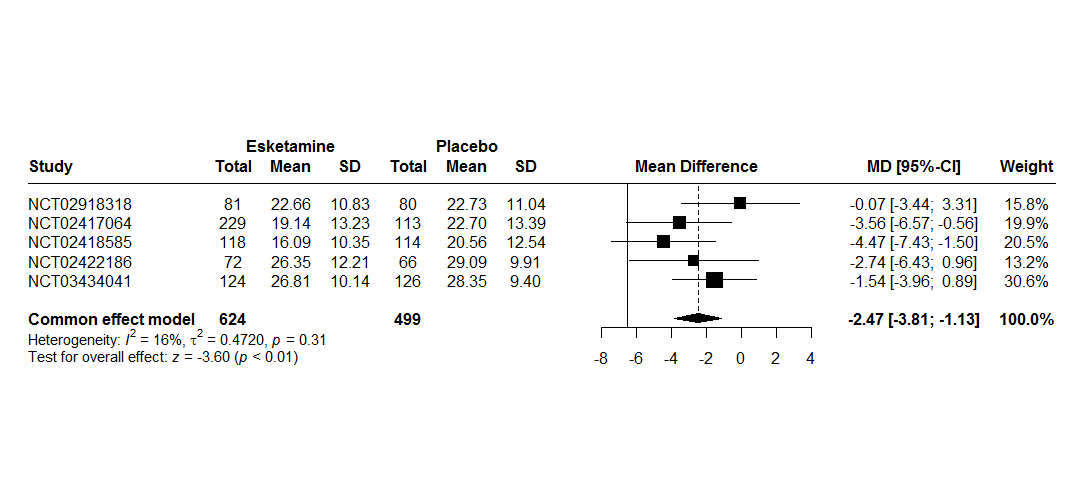

*MD : Mean Difference*

*CI : Confidence Interval*

###### 3.1.1.2 Meta-analysis of secondary outcomes

###### 3.1.1.2.1 Outcome assessed at week 1

**MADRS after at least 1 week**

Figure 5: Forest plot, MADRS after at least 1 week outcome

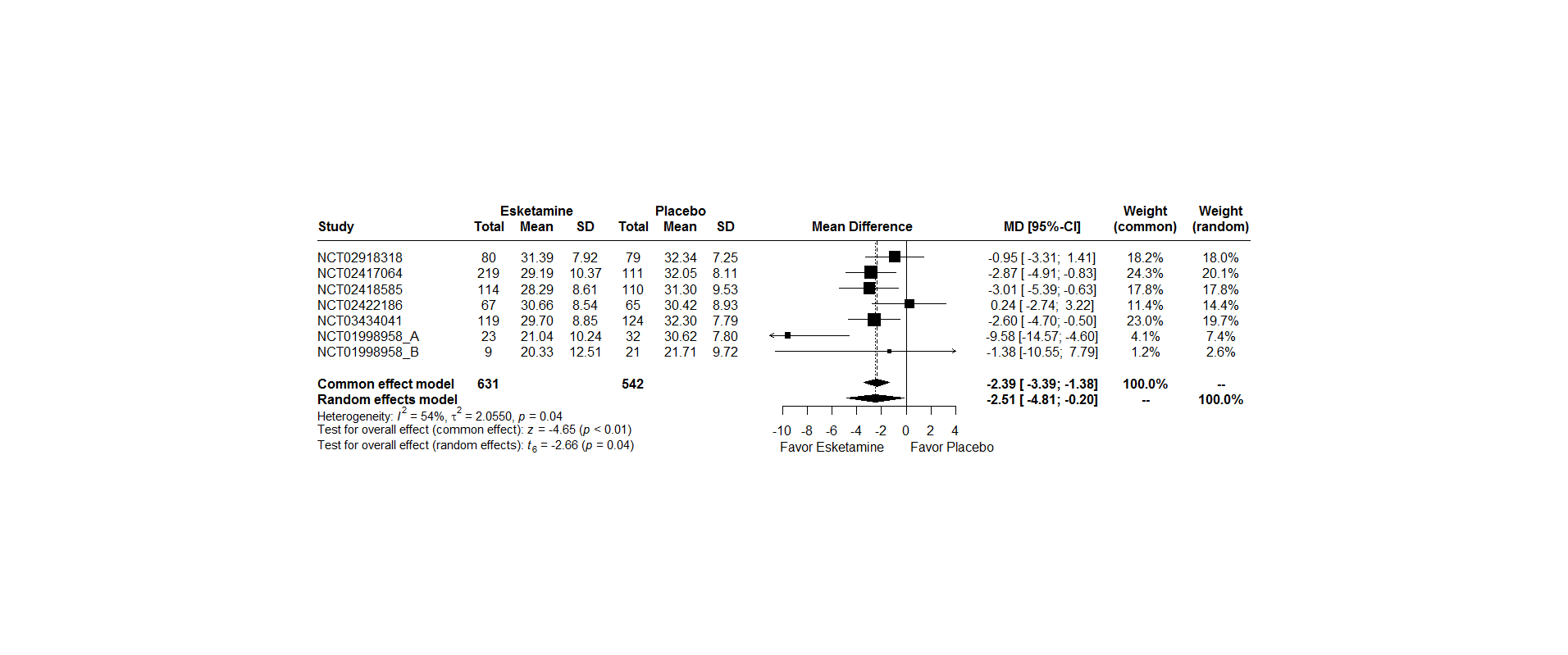

*MD : Mean Difference*

*CI : Confidence Interval*

A random effects model were used because there are heterogeneity between studies (I^2^ = 54%, t^2^ = 2.06, p = 0.04). The two-stage meta-analysis indicates a significant effect between MADRS at 1 week and treatment group with a beta of –2.39 [-3.39; -1.38], p < 0.01 in common effect model and a beta of -2.51 [-4.81; -0.20], p = 0.04 in a random effects model.

For studies NCT01998958, panel A and NCT01998958, panel B, the number of patients in the esketamine group is lower because patients randomized in the esketamine 28 mg group for panel A and esketamine 14 mg for panel B were excluded from the analysis (see section 1.2).

**MADRS after at least 1 weeks with imputation missing data**

Figure 6: Forest plot, MADRS after at least 1 week outcome with imputation missing data

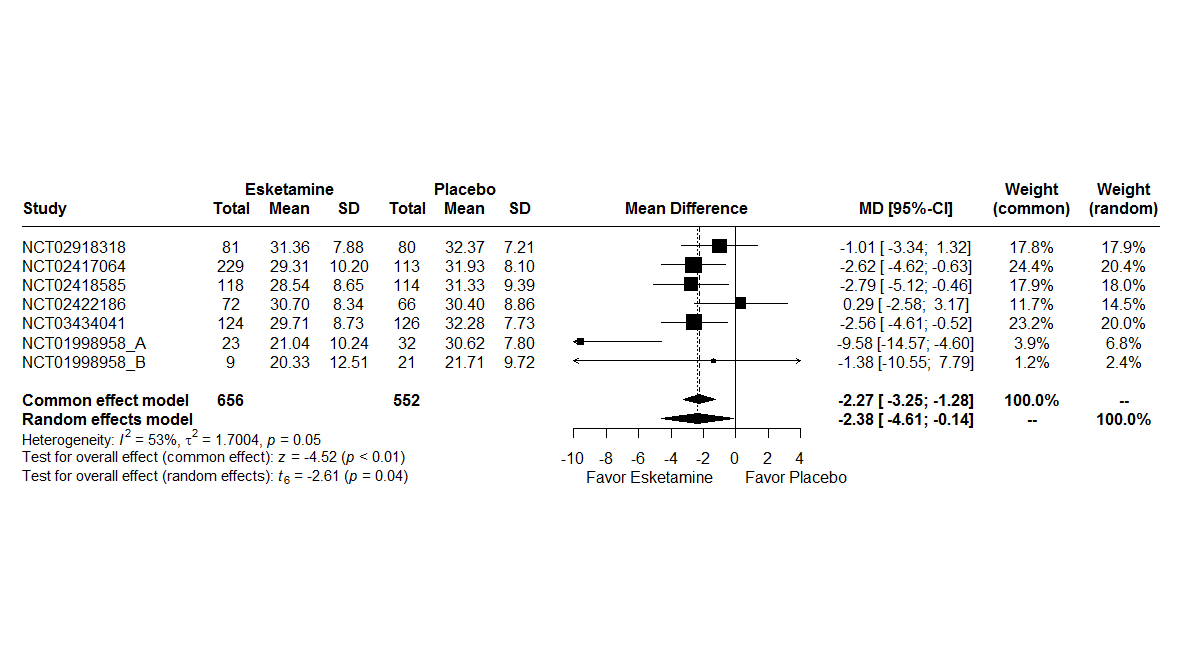

*MD : Mean Difference*

*CI : Confidence Interval*

A random effects model were used because there are heterogeneity between studies (I^2^ = 53%, t^2^ = 1.70, p = 0.05). The two-stage meta-analysis indicates a significant effect between MADRS at 1 week with imputation missing data and treatment group with a beta of –2.27 [-3.25; -1.28], p < 0.01 in common effect model and a beta of -2.38 [-4.61; -0.15], p =0.04 in a random effects model.

###### 3.1.1.2.2 Outcomes assessed at the end of treatment

**Suicide/suicide attempt**

To identify suicide attempts or suicides, the "Actual Attempt" and "Suicide" items of the Columbia-Suicide Severity Rating Scale (C-SSRS) part Suicidal behavior were selected, along with adverse events reported as "Suicide attempt", "Suicide", “Attempted suicide”, “Suicidal behavior”, “Completed suicide”. The Peto method was used to take account of rare events.

Figure 7: Forest plot, suicide/suicide attempt outcome

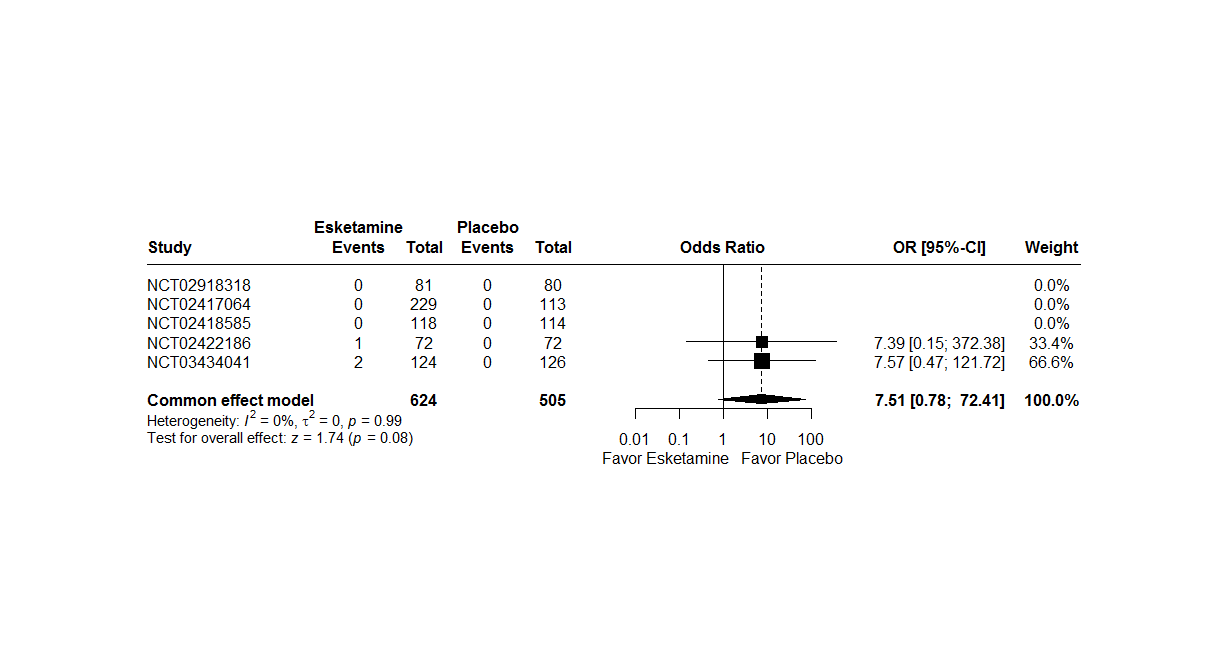

*RR : Relative Risk*

*CI : Confidence Interval*

A random effects model were not used because there not are heterogeneity between studies (I^2^ = 0%, t^2^ = 0, p = 0.99). The two-stage meta-analysis indicates a no significant effect between suicide/suicide attempt and treatment group with a RR of 7.51 [0.78; 72.41], p = 0.08 in common effects.

Table 11: Description Suicide/Suicide attempt

| NCT02422186 | Suicidal and self-injurious behaviour | 14 days after randomization | No serious event, the patient continue the treatment Esketamine until the end of the double blind phase (day 28) |
| --- | --- | --- | --- |
| NCT03434041 | Completed suicide | 4 days after randomization | Patient treated with Esketamine. He has died. More details in table 12 |
| NCT03434041 | Suicide attempt | 2 day after randomization | Patient treated with Esketamine. The double-blind phase and treatment is discountinued |

Table 12: Death

| Study | **Cause of death** | **Time of study** | **Arm** | **On esketamine treatment at time of death** | **Time since last treatment** | **Treatment-related death** |
| --- | --- | --- | --- | --- | --- | --- |
| 3002 | Road traffic accident and multiple injuries | Treatment (serious adverse event on day 16) | Esketamine | No | Clinical study report : Died 55 days after study start (40 days after last dose of treatment) | Clinical study report : according to the investigator, death unrelated to treatment |
| 3006 | Completed suicide | Treatment | Esketamine | Yes | Clinical study report : On study day 4 | Clinical study report : The investigator assessed the event of completed suicide as possibly to blinded study medication. In the subsequent evaluation, the sponsor took into account such factors as underlying severe depressive condition with worsening symptoms since the start of a major depressive episode several pharmacological treatments, the prior, and medication plan changes in the last month; based on the totality of evidence, the sponsor assessed the event as not related to esketamine treatment |

**Suicidal ideations**

To identify suicidal ideations, the Columbia-Suicide Severity Rating Scale (C-SSRS) part Suicidal ideation were selected, along with adverse events reported as "Suicide ideation", “Passive suicidal ideation”, “Suicidal intention”, “Depression suicidal”.

For C-SSRS, the suicidal ideation was defined if the sum of 5 items is greater than 3. The answer to questions 3, 4, 5 were available if answer question 2 was “yes”. The Peto method was used to take account of rare events.

Figure 8: Forest plot, suicidal ideations outcome

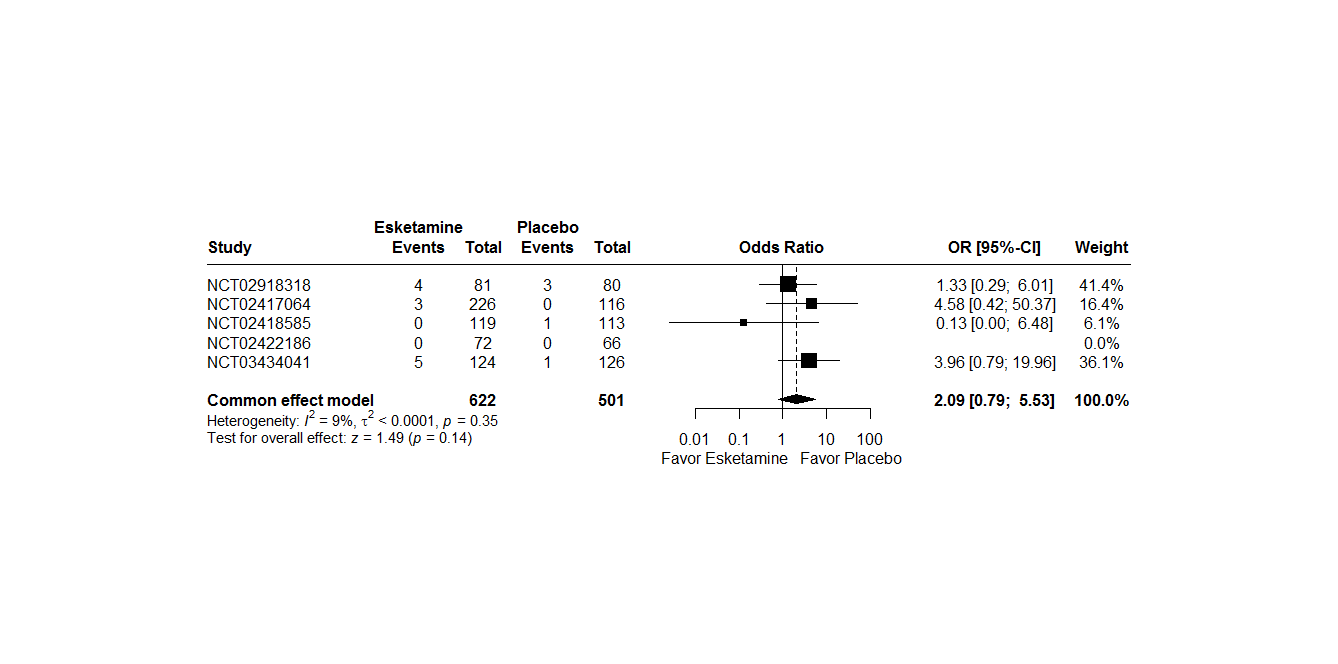

*RR : Relative Risk*

*CI : Confidence Interval*

Two studies don’t have events in two groups.

A random effects model were not used because there not are heterogeneity between studies (I^2^ = 9%, t^2^ <0.01, p = 0.35). The two-stage meta-analysis indicates a no significant effect between suicidal ideation and treatment group with a RR of 2.09 [0.79; 5.53], p = 0.14l in common effects.

**Remission**

For all studies, remission is defined as MADRS score ≤ 12.

Figure 9: Forest plot, remission outcome

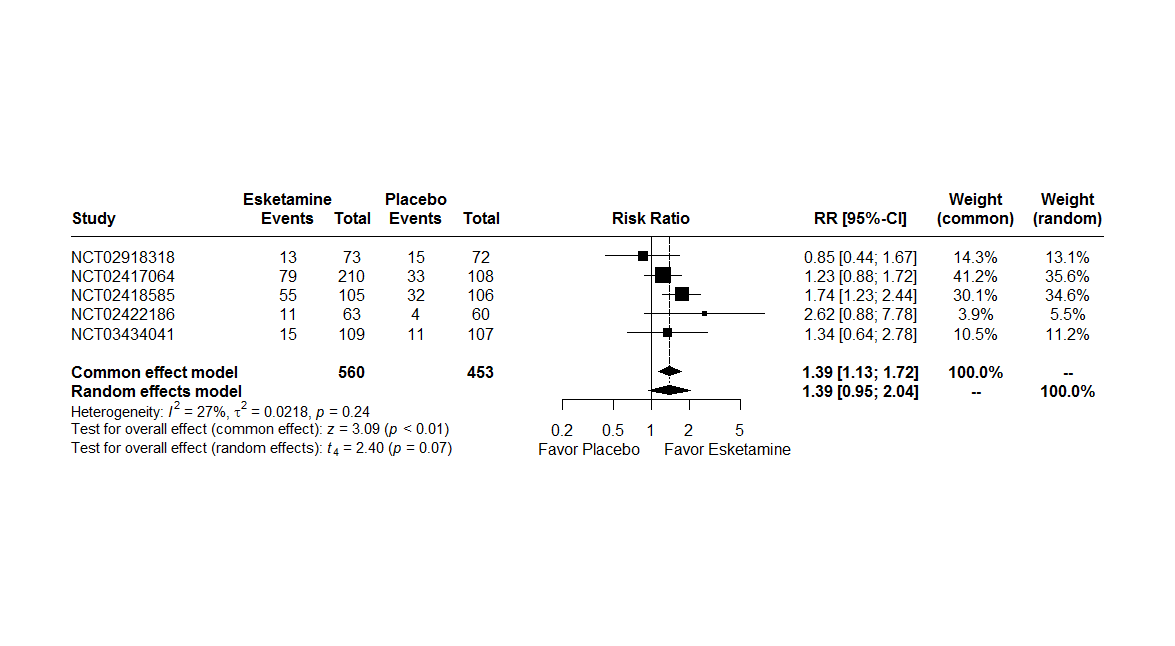

*RR : Relative Risk*

*CI : Confidence Interval*

A random effects model were used because there are heterogeneity between studies (I^2^ = 27%, t^2^ = 0.02, p = 0.24). The two-stage meta-analysis indicates a significant effect between remission and treatment group with a RR of 1.39 [1.13; 1.72], p < 0.01 in common effect model and a RR of 1.39 [0.95; 2.04], p = 0.07 in a random effects model.

**Remission with imputation missing data**

For all studies, remission is defined as MADRS score ≤ 12.

Figure 10: Forest plot, remission outcome with imputation missing data.

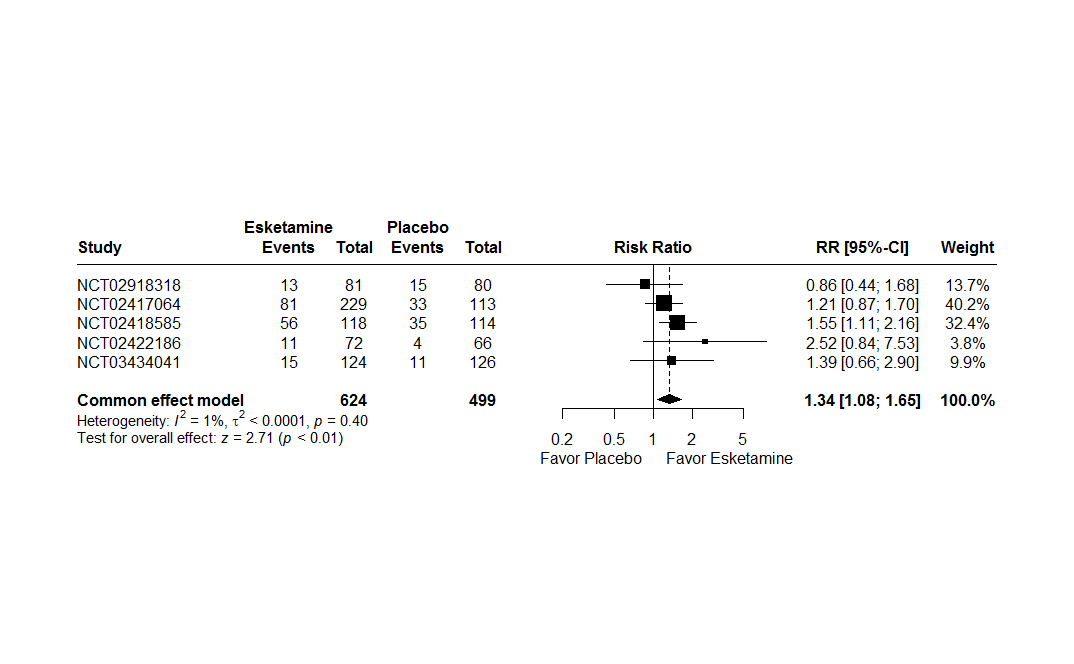

*RR : Relative Risk*

*CI : Confidence Interval*

A random effects model were not used because there are heterogeneity between studies (I^2^ = 1%, t^2^ = <0.01, p = 0.40). The two-stage meta-analysis indicates a significant effect between remission and treatment group with a RR of 1.34 [1.08; 1.65], p < 0.01 in common effect model.

**Sheehan Disability Scale**

The Sheehan Disability Scale (SDS) is a self-rated, 3-item questionnaire that uses from 0 (not at all) to 10 (extremely) to assess impairment in the family life/home responsibilities, social life, work/school domains. The three items are summed to obtain a total score (range: 0–30), with higher scores indicating greater functional impairment. If “I have not worked / studied” checked and/or work/school missing, first analysis the average of the family life/home responsibilities and social life items was imputed to work/school for calculated total score and second analysis the total score was considered missing.

First analysis:

Figure 11: Forest plot, Sheehan Disability Scale outcome

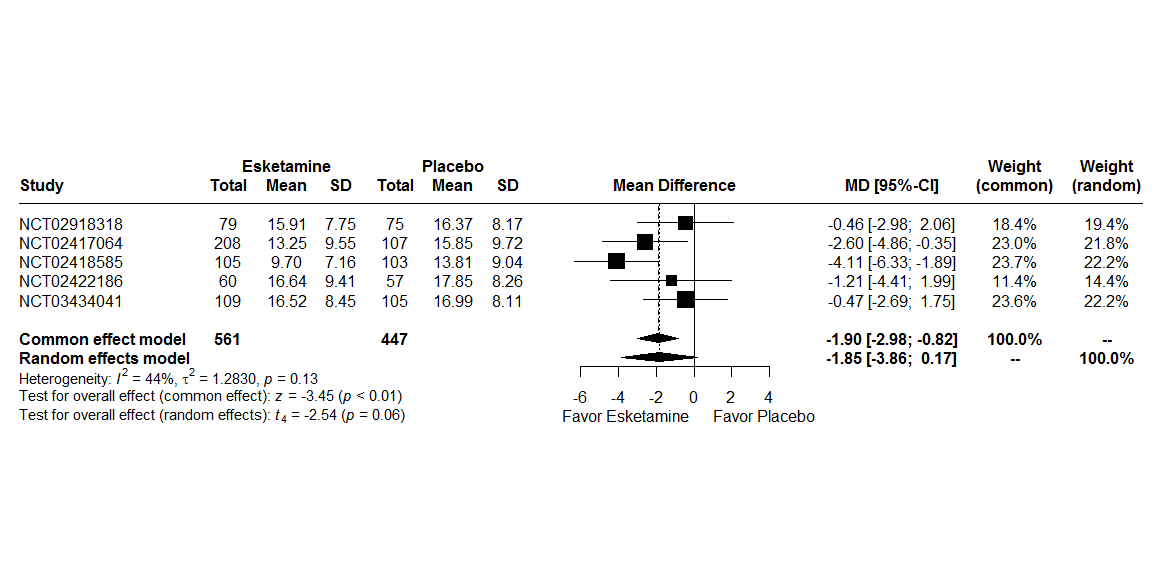

*MD : Mean Difference*

*CI : Confidence Interval*

A random effects model were used because there are heterogeneity between studies (I^2^ = 44%, t^2^ = 1.28, p = 0.13). The two-stage meta-analysis indicates a significant effect between SDS and treatment group with a beta of –1.90 [-2.98; -0.82], p < 0.01 in common effect model and a beta of -1.85 [-3.86; 0.17], p = 0.06 in a random effects model.

Second analysis:

Figure 12: Forest plot, Sheehan Disability Scale outcome

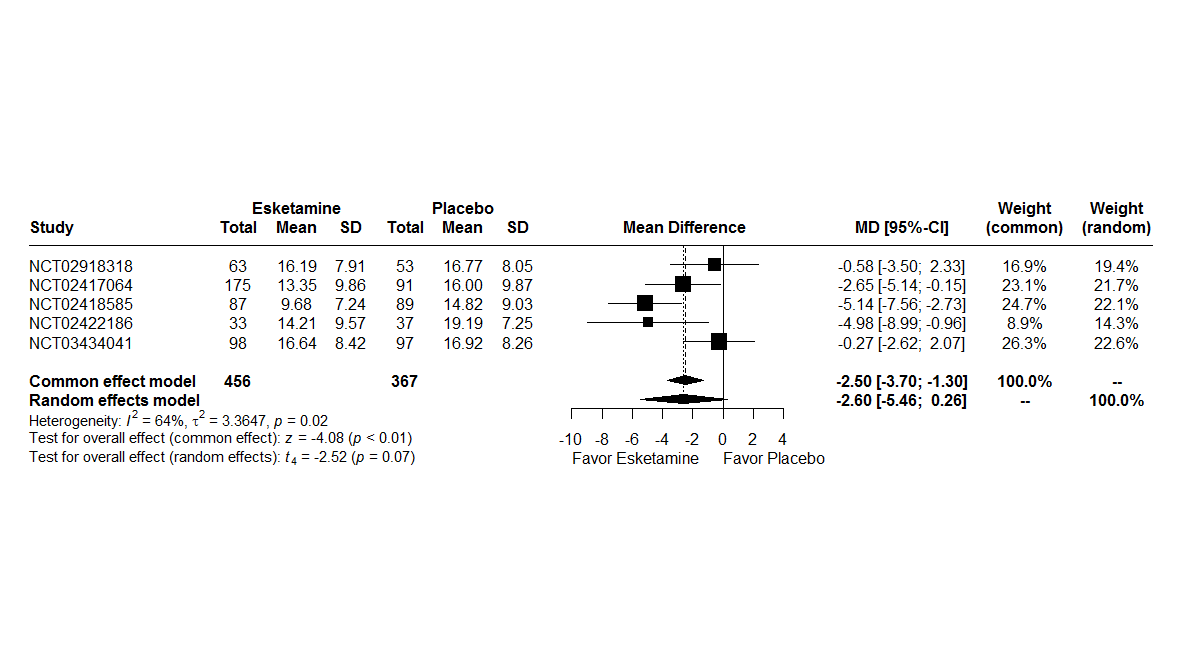

*MD : Mean Difference*

*CI : Confidence Interval*

A random effects model were used because there are heterogeneity between studies (I^2^ = 64%, t^2^ = 3.36, p = 0.02). The two-stage meta-analysis indicates a significant effect between SDS and treatment group with a beta of –2.50 [-3.70; -1.30], p < 0.01 in common effect model and no significant effect with a beta of -2.60 [-5.46; 0.26], p = 0.07 in a random effects model.

**Sheehan Disability Scale with imputation missing data**

First analysis:

Figure 13: Forest plot, Sheehan Disability Scale outcome with imputation missing data

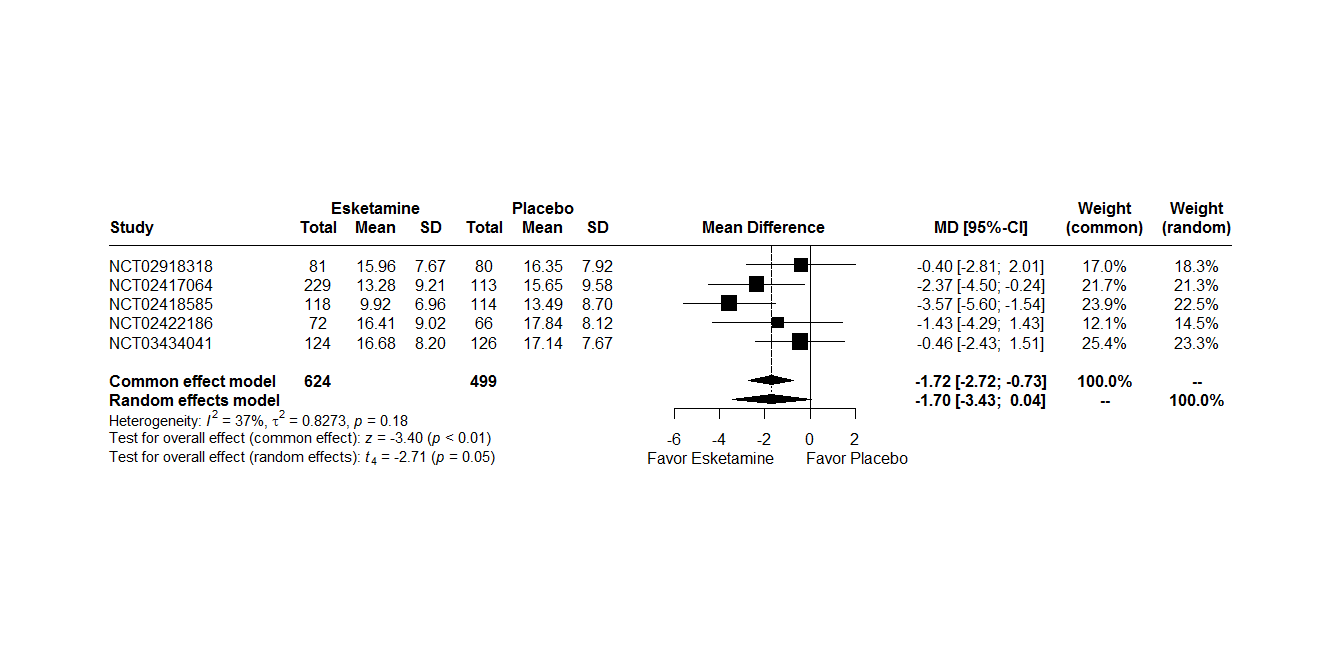

*MD : Mean Difference*

*CI : Confidence Interval*

A random effects model were used because there are heterogeneity between studies (I^2^ = 37%, t^2^ = 0.83, p = 0.18). The two-stage meta-analysis indicates a significant effect between SDS and treatment group with a beta of –1.72 [-2.72; -0.73], p < 0.01 in common effect model and no significant effect with a beta of -1.70 [-3.43; -0.04], p = 0.05 in a random effect model.

Second analysis:

Figure 14: Forest plot, Sheehan Disability Scale outcome with imputation missing data

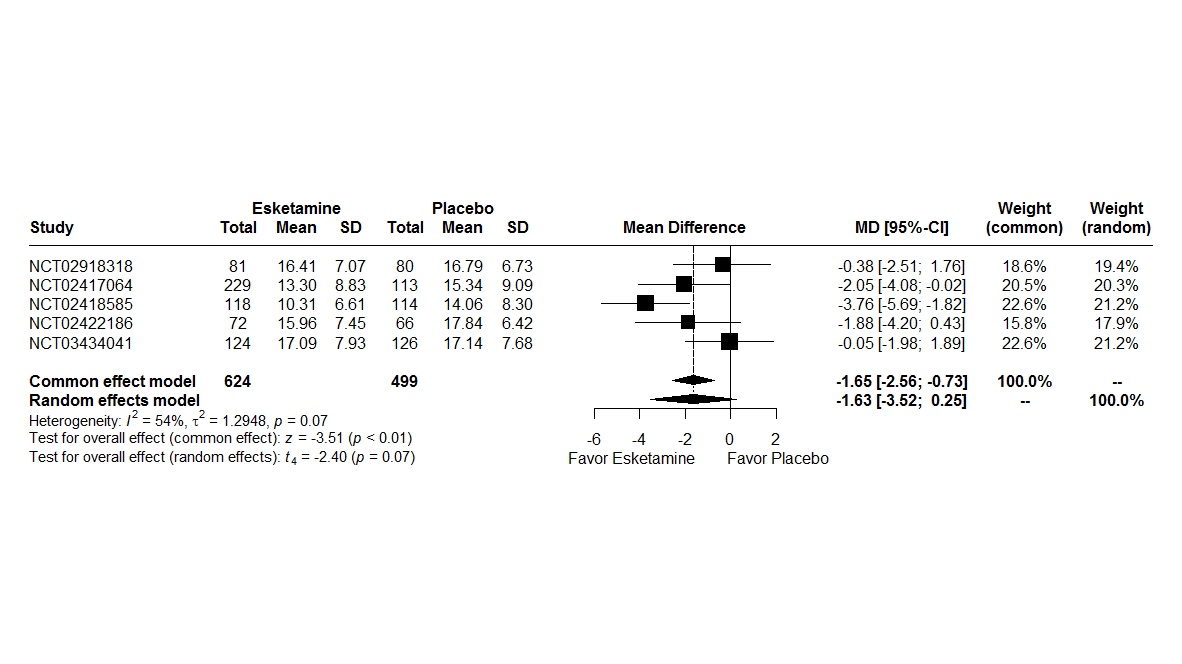

*MD : Mean Difference*

*CI : Confidence Interval*

A random effects model were used because there are heterogeneity between studies (I^2^ = 54%, t^2^ = 1.29, p = 0.07). The two-stage meta-analysis indicates a significant effect between SDS and treatment group with a beta of –1.65 [-2.56; -0.73], p < 0.01 in common effect model and no significant effect with a beta of -1.63 [-3.52; 0.25], p = 0.07 in a random effect model.

**PHQ-9**

The Patient Health Questionnaire – 9-item (PHQ-9), I a 9-item, subject-reported outcome measure that was used to assess depressive symptoms. The scale scores each of the 9 symptom domains of the DSM-5 MDD criteria, and it has been used both as a screening tool and a measure of response to treatment for depression. Each item is rated on a 4 point scale (0=not at all, 1=several days, 2=more than half the days and 3=nearly every day). The subject’s item responses are summed to provide a total score (range of 0 to 27) with higher scores indicating greater severity of depressive symptoms.

Figure 15: Forest plot, PHQ-9 outcome

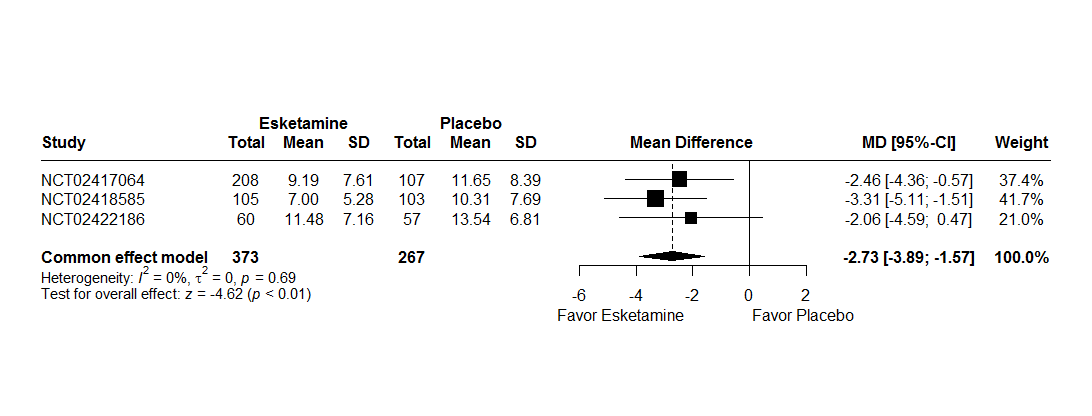

*MD : Mean Difference*

*CI : Confidence Interval*

Three studies were realized PHQ-9. A random effects model were 4 used because there are 4 heterogeneity between studies (I^2^ = 0%, t^2^ = 0, p = 0.69). The two-stage meta-analysis indicates a no significant effect between PHQ-9 and treatment group with a beta of –2.73 [-3.89; -1.57], p < 0.01 in common effect model.

**PHQ-9 with imputation missing data**

Figure 16: Forest plot, PHQ-9 outcome with imputation missing data

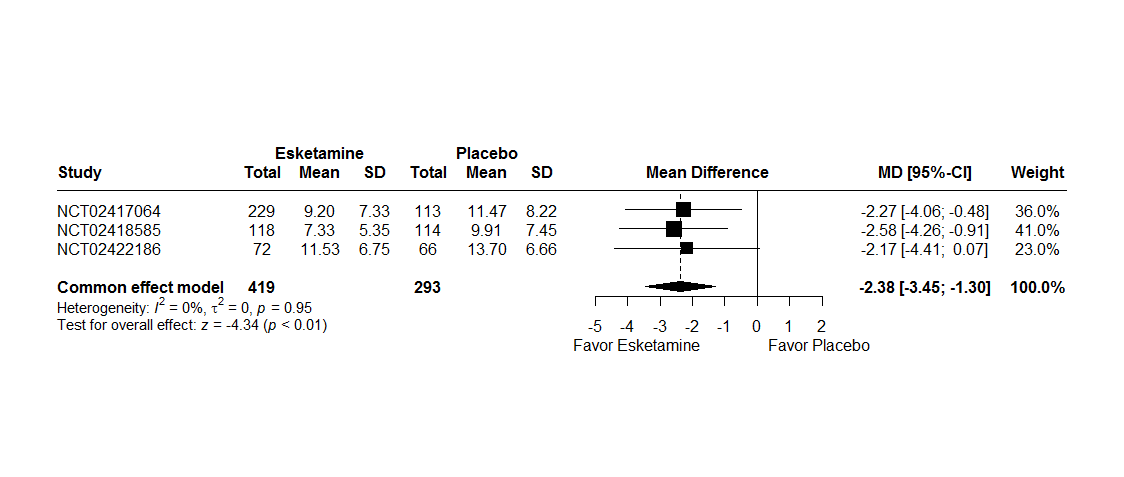

Three studies were realized PHQ-9. A random effects model were not used because there are not heterogeneity between studies (I^2^ = 0%, t^2^ = 0, p = 0.95). The two-stage meta-analysis indicates a significant effect between PHQ-9 and treatment group with a beta of –2.38 [-3.45; -1.30], p < 0.01 in common effect model.

###### 3.1.1.2.3 Outcomes assessed at the last follow-up visit post-treatment

For analysis at the last follow-up visit post-treatment, two studies were analysed.

Reasons for no analysis other studies:

- NCT02422186 (3005) : follow-up phase included patient with received at least 1 dose of intranasal study medication in the double-blind phase and either withdrew early before the end of the double-blind induction phase, or chose not to participate in the other (ESKETINTRD3004) safety study
- NCT02418585 (3002): follow-up phase included all subjects who were not eligible or who chose to not participate in the maintenance of effect study NCT02493868 (3003).
- NCT02417064 (3001): follow-up included subjects who receive at least 1 dose of intranasal study medication in the double-blind induction phase, but do not enter the subsequent maintenance clinical study NCT02493868 (3003).

**Suicide/suicide attempt**

To identify suicide attempts or suicides, the "Actual Attempt" and "Suicide" items of the Columbia-Suicide Severity Rating Scale (C-SSRS) part Suicidal behavior were selected, along with adverse events reported as "Suicide attempt", "Suicide", “Attempted suicide”, “Suicidal behavior”, “Completed suicide”. The Peto method was used to take account of rare events.

Figure 17: Forest plot, suicide/suicide attempt outcome during the follow-up period.

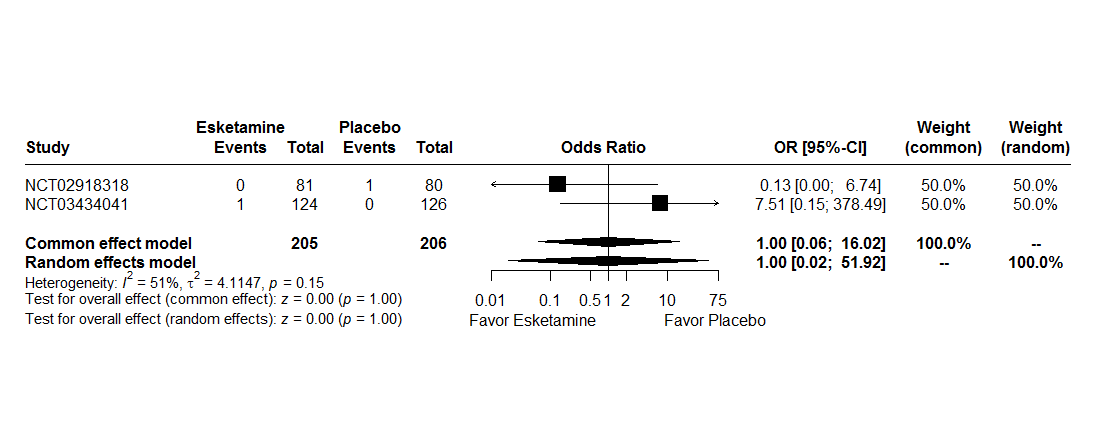

*RR : Relative Risk*

*CI : Confidence Interval*

A random effects model were not used because there are heterogeneity between studies (I^2^ = 51%, t^2^ = 4.11, p = 0.15). The two-stage meta-analysis indicates a no significant effect between Suicide/suicide attempt and treatment group with a RR of 1.00 [0.06; 16.02], p = 1.00 in common effect model and a beta of 1.00 [0.02; 51.92], p = 1.00 in a random effects model.

Table 13: Description suicide attempt at the last follow-up visit post-treatment

| **Study** | **Description** | **Treatment during event** |
| --- | --- | --- |
| NCT02918318 | Two episodes the same day | No treatment |
| NCT03434041 | The same subject who attempted suicide in the double-blind phase | Oral antidepressant (Setraline) |

**Suicidal ideations**

To identify suicidal ideations, the Columbia-Suicide Severity Rating Scale (C-SSRS) part Suicidal ideation were selected, along with adverse events reported as "Suicide ideation", “Passive suicidal ideation”, “Suicidal intention”, “Depression suicidal”.

For C-SSRS, the suicidal ideation was defined if the sum of 5 items is greater than 3. The answer to questions 3, 4, 5 were available if answer question 2 was “yes”. The Peto method was used to take account of rare events.

Figure 18: Forest plot, suicidal ideations outcome during the follow-up period

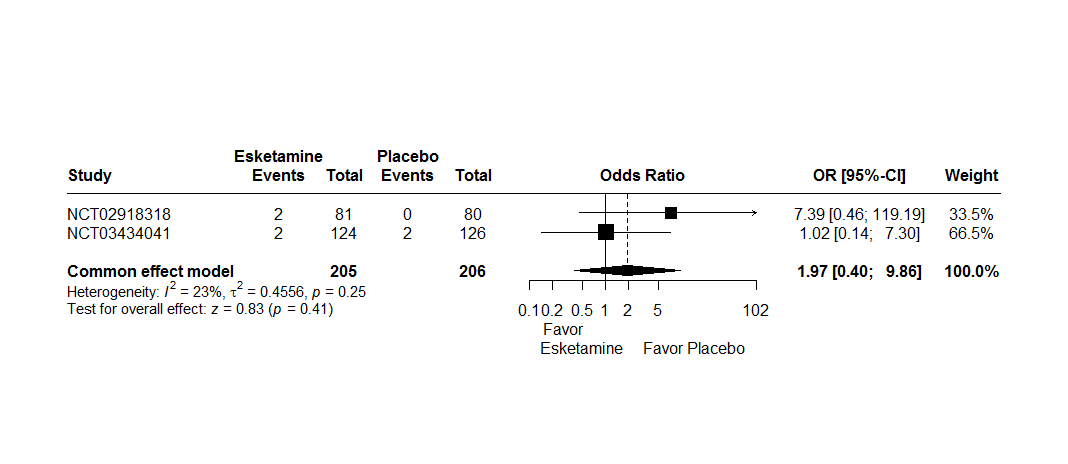

*RR : Relative Risk*

*CI : Confidence Interval*

A random effects model were not used because there not are heterogeneity between studies (I^2^ = 23%, t^2^ = 0.46, p = 0.25). The two-stage meta-analysis indicates a no significant effect between suicidal ideation and treatment group with a RR of 1.97 [0.40; 9.86], p = 0.41 in common effects.

Table 14: Description suicide attempt at the last follow-up visit post-treatment

| **Study** | **Study medication in double-blind phase** | **Description** | **Study medication during event** |
| --- | --- | --- | --- |
| NCT02918318 | Esketamine | C-SSR > 3 | No study medication |
| NCT02918318 | Esketamine | C-SSR > 3 | No study medication |
| NCT03434041 | Esketamine | C-SSR > 3 | No study medication |
| NCT03434041 | Placebo | C-SSR > 3 | No study medication |
| NCT03434041 | Placebo | Adverse event | No study medication |
| NCT03434041 | Esketamine | Adverse event | No study medication |

**MADRS**

The total duration of a posttreatment follow-up (FU) phase in study NCT03434041 (3006) was 8-week. For study NCT02918318 (2005), responders (subject who have ≥50% reduction from baseline in MADRS total score) at the end of the double-blind (DB) induction phase were eligible to proceed to the posttreatment phase; thode who do bot (ie, nonresponders) were proceed to the 4-week follow-up phase, the 8-week posttreatment were selected for to match 4-week follow-up (4-week DB + 4-week posttreatment).

MADRS score was selected if patient was realized the last visit of follow-up provided in the protocol.

Figure 19: Forest plot, MADRS outcome at the last follow-up visit post-treatment

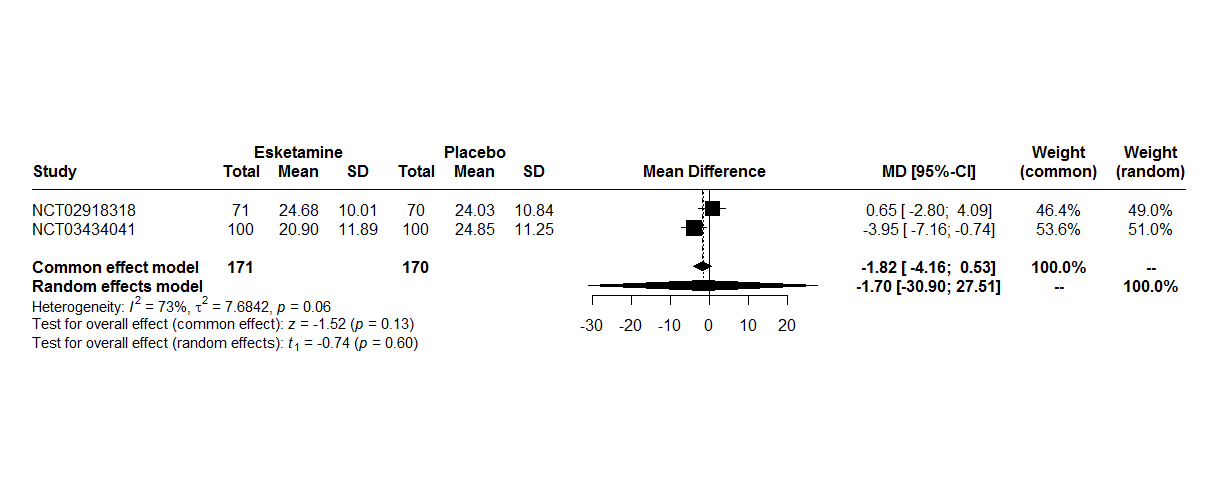

*MD : Mean Difference*

*CI : Confidence Interval*

A random effects model were used because there are heterogeneity between studies (I^2^ = 73%, t^2^ = 7.68, p = 0.06). The two-stage meta-analysis indicates a no significant effect between MADRS at follow-up phase and treatment group with a beta of -1.82 [-4.16; 0.53], p = 0.13 in common effect model and a beta of -1.70 [-30.90; 27.51], p = 0.60 in a random effects model.

**MADRS with imputation missing data**

Figure 20: Forest plot, MADRS outcome with imputation missing data at the last follow-up visit post-treatment

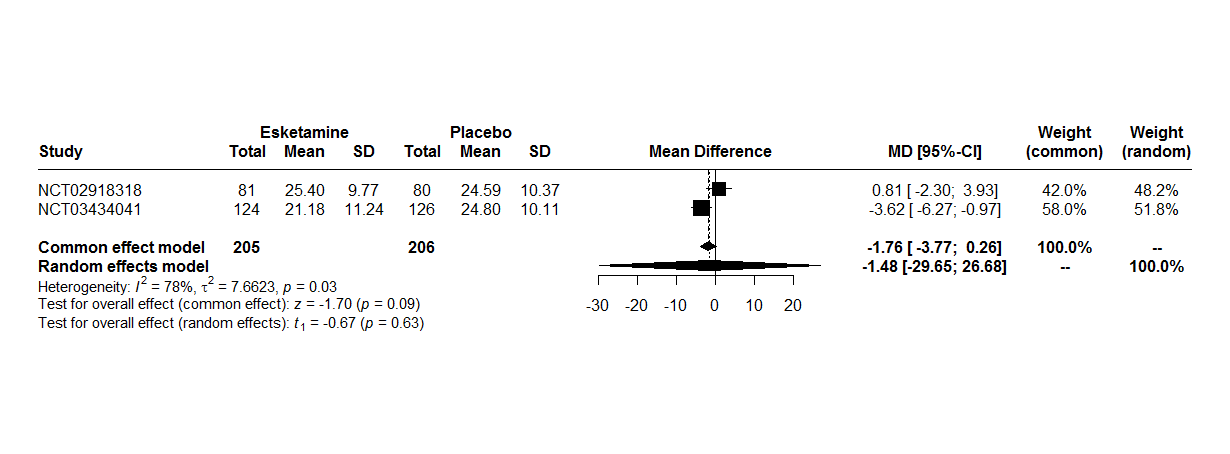

*MD : Mean Difference*

*CI : Confidence Interval*

A random effects model were used because there are heterogeneity between studies (I^2^ = 78%, t^2^ = 7.66, p = 0.03). The two-stage meta-analysis indicates a no significant effect between MADRS at follow-up phase and treatment group with a beta of -1.76 [-3.77; 0.26], p = 0.09 in common effect model and a beta of -1.48 [-29.65; 26.68], p = 0.63 in a random effects model.

**Remission**

For all studies, remission is defined as MADRS score ≤ 12.

Figure 21: Forest plot, remission outcome at the last follow-up visit post-treatment

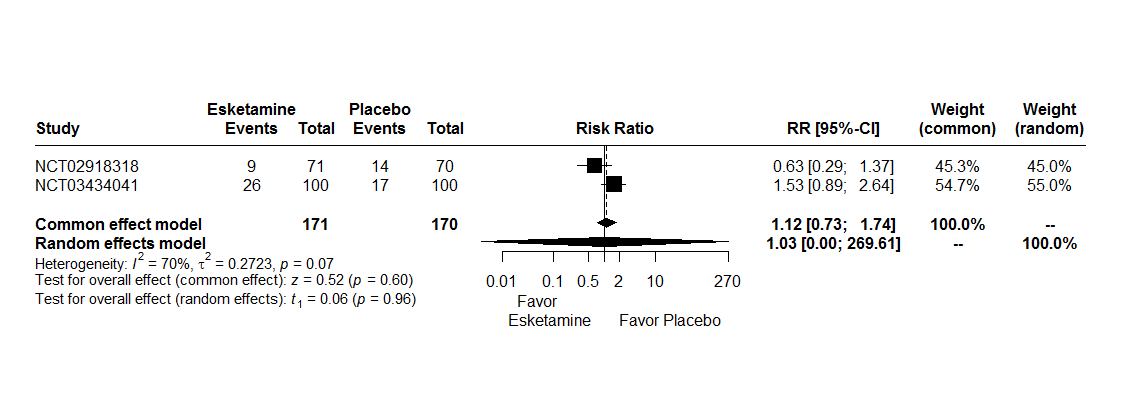

*RR : Relative Risk*

*CI : Confidence Interval*

A random effects model were used because there are heterogeneity between studies (I^2^ = 70%, t^2^ = 0.27, p = 0.07). The two-stage meta-analysis indicates a no significant effect between remission and treatment group with a RR of 1.12 [0.73; 1.74], p = 0.60 in common effect model and a RR of 1.03 [0.00; 269.61], p = 0.96 in a random effects model.

**Remission with imputation missing data**

For all studies, remission is defined as MADRS score ≤ 12.

Figure 22: Forest plot, remission with imputation missing data outcome at the last follow-up visit post-treatment

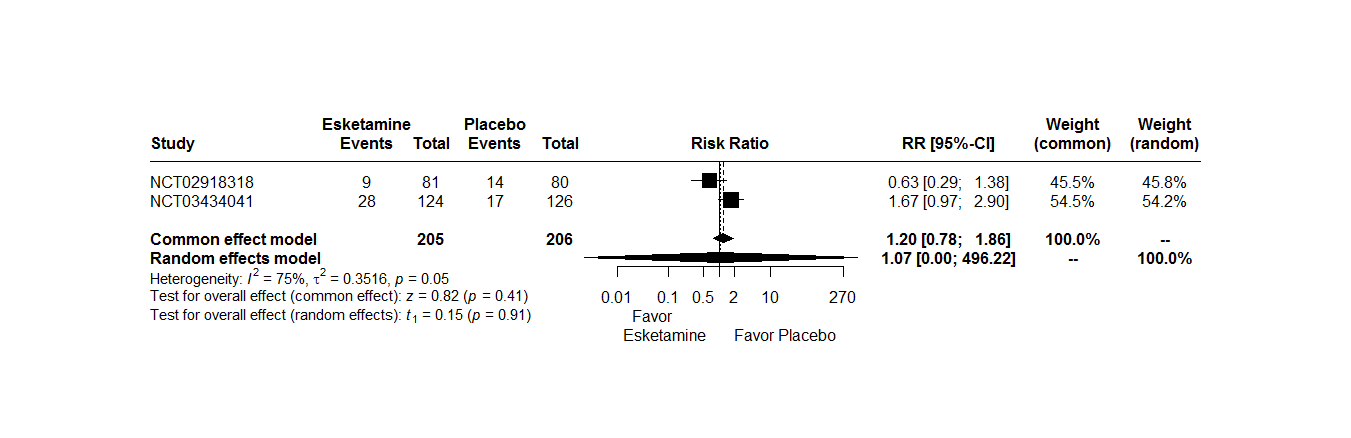

Interpretation:

A random effects model were used because there are heterogeneity between studies (I^2^ = 75%, t^2^ = 0.35, p=0.05). The two-stage meta-analysis indicates a no significant effect between remission and treatment group with a RR of 1.20 [0.78; 1.86], p=0.41 in common effect model and a RR of 1.07 [0.00; 496.22], p=0.91 in a random effects model.

**Sheehan Disability Scale**

The Sheehan Disability Scale (SDS) is a self-rated, 3-item questionnaire that uses from 0 (not at all) to 10 (extremely) to assess impairment in the family life/home responsibilities, social life, work/school domains. The three items are summed to obtain a total score (range: 0–30), with higher scores indicating greater functional impairment. If “I have not worked / studied” checked and/or work/school missing, first analysis the average of the family life/home responsibilities and social life items was imputed to work/school for calculated total score and second analysis the total score was considered missing.

Only study NCT03434041 (3006) have a data to SDS at follow-up phase. No forest-plot performed.

First analysis:

|  | Esketamine | Placebo |
| --- | --- | --- |
| n | 95 | 95 |
| Mean | 13.01 | 14.58 |
| Standard deviation | 8.73 | 8.63 |

Result of ANOVA:

|  | **Estimate [Confidence Interval 95%]** | **P-value** |
| --- | --- | --- |
| Esketamine vs. Placebo | -1.57 [-4.05 ; 0.92] | 0.2146 |

Second analysis:

|  | Esketamine | Placebo |
| --- | --- | --- |
| n | 91 | 85 |
| Mean | 13.55 | 14.89 |
| Standard deviation | 8.82 | 8.61 |

Result of ANOVA:

|  | **Estimate [Confidence Interval 95%]** | **P-value** |
| --- | --- | --- |
| Esketamine vs. Placebo | -1.34 [-3.93 ; 1.26] | 0.3103 |

**Sheehan Disability Scale with imputation missing data**

Only study NCT03434041 (3006) have a data to SDS at follow-up phase. No forest-plot performed.

First analysis:

|  | Esketamine | Placebo |
| --- | --- | --- |
| n | 124 | 126 |
| Mean | 13.16 | 14.74 |
| Standard deviation | 8.43 | 7.96 |

Result of ANOVA:

|  | **Estimate [Confidence Interval 95%]** | **P-value** |
| --- | --- | --- |
| Esketamine vs. Placebo | -1.58 [-3.62 ; 0.46] | 0.1288 |

Second analysis:

|  | Esketamine | Placebo |
| --- | --- | --- |
| n | 124 | 126 |
| Mean | 13.61 | 15.13 |
| Standard deviation | 8.15 | 7.94 |

Result of ANOVA:

|  | **Estimate [Confidence Interval 95%]** | **P-value** |
| --- | --- | --- |
| Esketamine vs. Placebo | -1.52 [-3.52 ; 0.49] | 0.1371 |

**PHQ-9**

No study has PHQ-9 data at the follow-up phase.

##### 3.1.2 Safety outcomes

###### 3.1.2.1 Outcomes assessed at the end of treatment

**Serious adverse events**

Figure 23: Forest plot, serious adverse events outcome

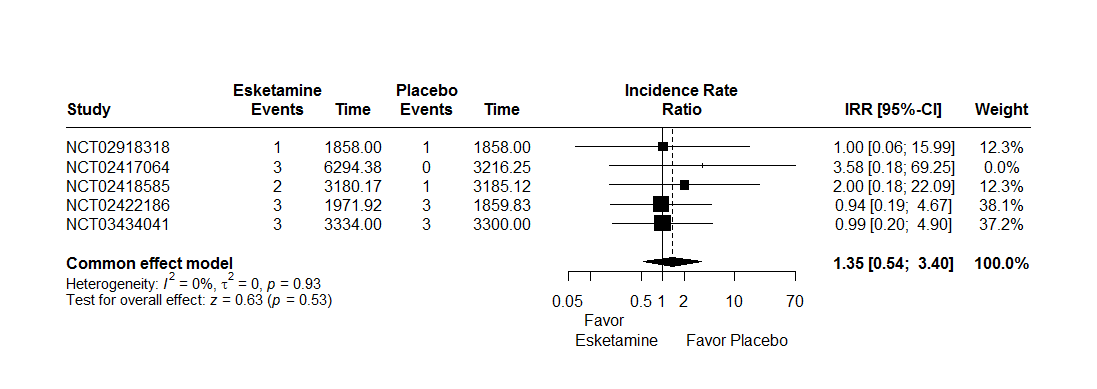

*IRR : Incidence Rate Ratio*

*CI : Confidence Interval*

A random effects model were not used because there are not heterogeneity between studies (I^2^ = 0%, t^2^ = 0, p = 0.93). The two-stage meta-analysis indicates a no significant effect between serious adverse events and treatment group with an IRR of 1.35 [0.54; 3.40], p = 0.53 in common effect model.

Table 15: Description serious adverse events

| **Study** | **AELLT** | **Group** |
| --- | --- | --- |
| 3006 | Recurrent depressive disorder | PLACEBO |
| 3006 | Recurrent depressive disorder | ESKETAMINE |
| 3006 | Recurrent depressive disorder | PLACEBO |
| 3006 | Completed suicide | ESKETAMINE |
| 3006 | Suicide attempt | ESKETAMINE |
| 3006 | Recurrent depressive disorder | PLACEBO |
| 3005 | Feeling of despair | PLACEBO |
| 3005 | Gait disturbance | PLACEBO |
| 3005 | Broken hip | ESKETAMINE |
| 3005 | Blood pressure increased | ESKETAMINE |
| 3005 | Anxiety disorder | ESKETAMINE |
| 3005 | Dizziness | PLACEBO |
| 3002 | Benign paroxysmal positional vertigo | PLACEBO |
| 3002 | Multiple injuries | ESKETAMINE |
| 3002 | Road traffic accident | ESKETAMINE |
| 3001 | Depression worsened | ESKETAMINE |
| 3001 | Depression aggravated | ESKETAMINE |
| 3001 | Headache | ESKETAMINE |
| 2005 | Suicidal ideation | PLACEBO |
| 2005 | Suicidal ideation | ESKETAMINE |

**Dropout for any cause**

Dropout for any cause is considered if patient checked at least one of the following criteria:

- Adverse event
- Death
- Lack of efficacy
- Lost to follow-up
- Non-compliance with switched oral antidepressant therapy
- Other
- Withdrawal by subject
- Investigator decision
- Protocol violation

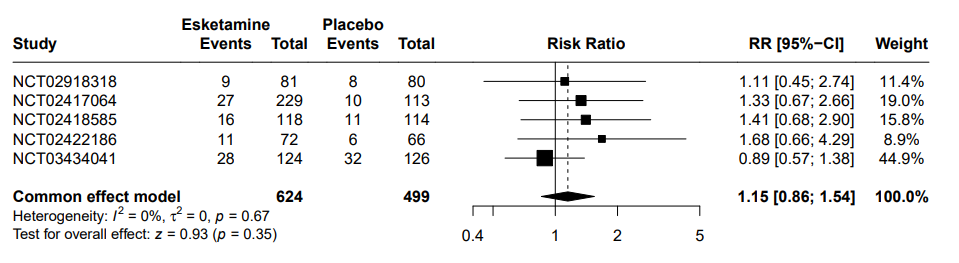
Figure 24: Forest plot, dropout due to any cause outcome

*RR : Relative Risk*

*CI : Confidence Interval*

A random effects model were not used because there are not heterogeneity between studies (I^2^ = 0%, t^2^ = 0, p = 0.67). The two-stage meta-analysis indicates a no significant effect between dropout for any cause and treatment group with an RR of 1.15 [0.86; 1.54], p = 0.35 in common effect model.

**Dropout due to adverse events**

Figure
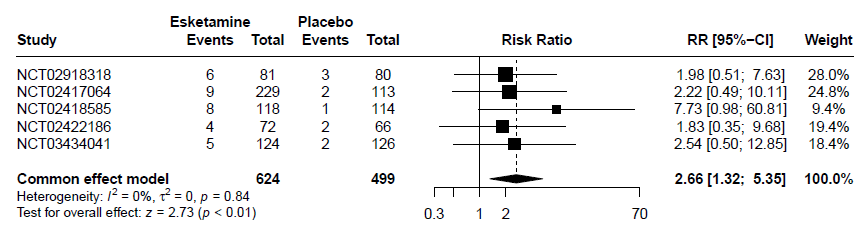
 25: Forest plot, dropout due to adverse events outcome

*RR : Relative Risk*

*CI : Confidence Interval*

A random effects model were not used because there are not heterogeneity between studies (I^2^ = 0%, t^2^ = 0, p = 0.84). The two-stage meta-analysis indicates a significant effect between dropout due to adverse events and treatment group with an RR of 2.66 [1.32; 5.35], p < 0.01 in common effect model.

**Adverse events**

Figure 26: Forest plot, adverse events outcome

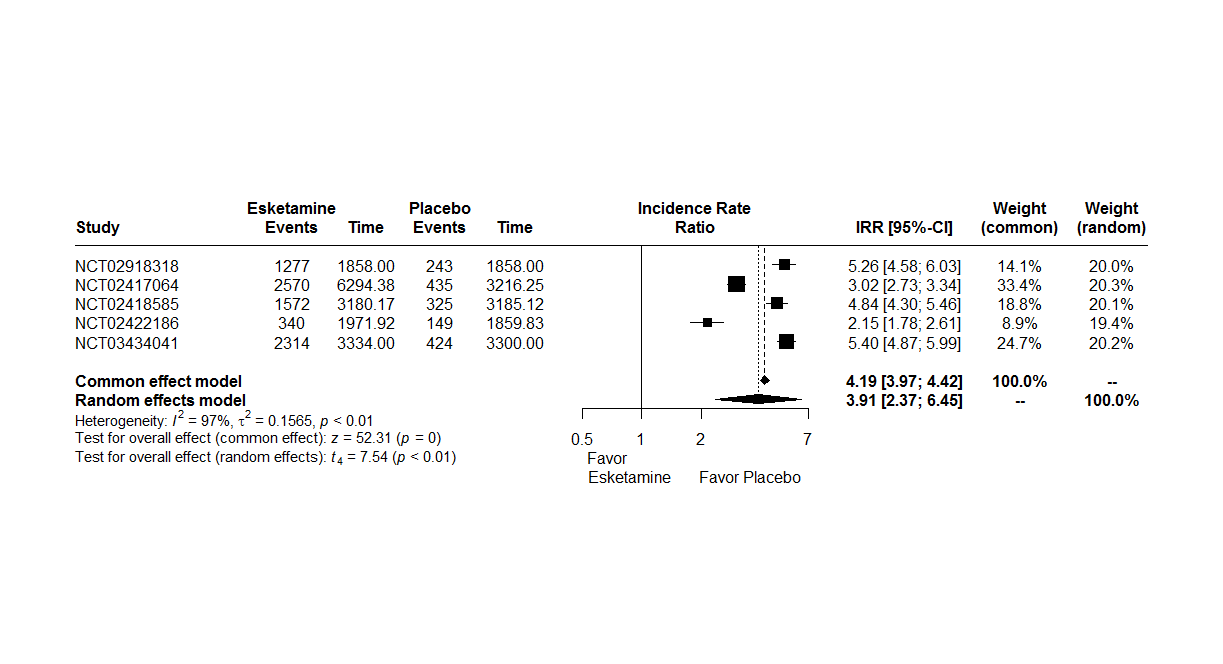

*IRR : Incidence Rate Ratio*

*CI : Confidence Interval*

A random effects model were used because there are heterogeneity between studies (I^2^ = 97%, t^2^ = 0.15, p < 0.01). The two-stage meta-analysis indicates a significant effect between adverse events and treatment group with an IRR of 3.91 [2.37; 6.45], p < 0.01 in random effects model.

**Blood pressure**

The analysis of blood pressure was realized with one-stage individual participant’s data meta-analysis. The blood pressure was measured at predose and at 40 minutes, 1 hour and 1.5 hours postdose and repeated during the double-blind induction phase.

A linear mixed model was realized with blood pressure as response variable, treatment and measurement interaction (predose and at 40 minutes, 1 hour and 1.5 hours postdose) and treatment group and day effect as random effect and stratified by trial the intercept.

Diastolic blood pressure per day and per time

Figure 27: Evolution of diastolic blood pressure by measurement time and per day according to group

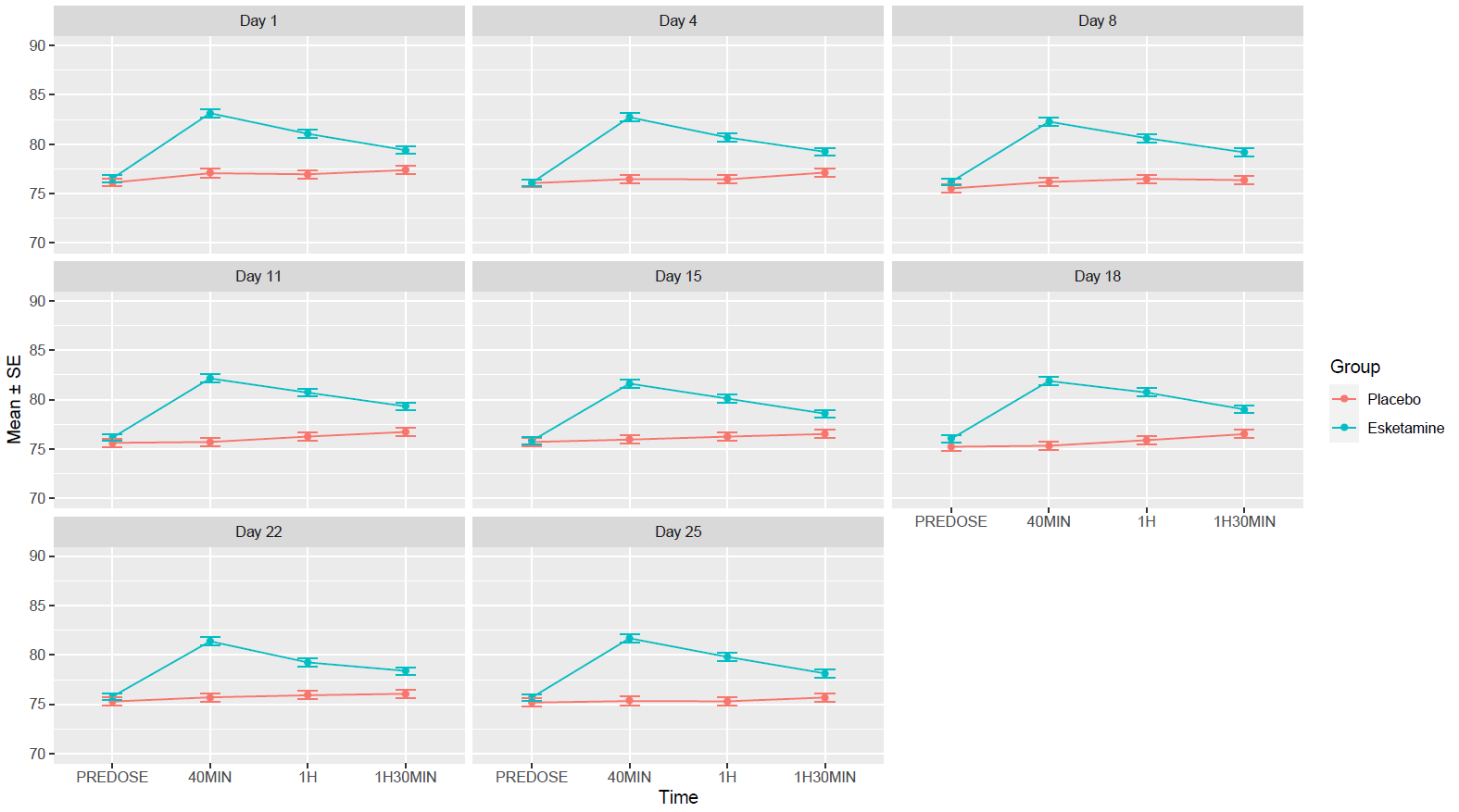

Table 16 : Evolution of diastolic blood pressure by measurement time and per day according to group, mean difference [Confidence Interval(CI) 95%]

| **Day** | **Time** | **Mean difference [CI95%]** |
| --- | --- | --- |
| Day 1 |  |  |
|  | PREDOSE | 0.41 [-0.61 ; 1.45] |
|  | 40 minutes | 6.09 [4.92 ; 7.26] |
|  | 1 hour | 4.10 [2.94 ; 5.27] |
|  | 1 hour and 30 minutes | 2.02 [0.95 ; 3.09] |
| Day 4 |  |  |
|  | PREDOSE | 0.04 [-1.00 ; 1.08] |
|  | 40 minutes | 6.26 [5.06 ; 7.47] |
|  | 1 hour | 4.25 [3.08 ; 5.43] |
|  | 1 hour and 30 minutes | 2.11 [0.98 ; 3.24] |
| Day 8 |  |  |
|  | PREDOSE | 0.66 [-0.44 ; -1.76] |
|  | 40 minutes | 6.11 [4.89 ; 7.33] |
|  | 1 hour | 4.12 [2.96 ; 5.29] |
|  | 1 hour and 30 minutes | 2.81 [1.68 ; 3.94] |
| Day 11 |  |  |
|  | PREDOSE | 0.52 [-0.54 ; 1.58] |
|  | 40 minutes | 6.44 [5.26 ; 7.62] |
|  | 1 hour | 4.45 [3.28 ; 5.61] |
|  | 1 hour and 30 minutes | 2.60 [1.47 ; 3.74] |
| Day 15 |  |  |
|  | PREDOSE | 0.12 [-0.97 ; 1.20] |
|  | 40 minutes | 5.67 [4.50 ; 6.84] |
|  | 1 hour | 3.86 [2.67 ; 5.03] |
|  | 1 hour and 30 minutes | 2.06 [0.95 ; 3.18] |
| Day 18 |  |  |
|  | PREDOSE | 0.80 [-0.29 ; 1.89] |
|  | 40 minutes | 6.56 [5.39 ; 7.73] |
|  | 1 hour | 4.83 [3.66 ; 6.00] |
|  | 1 hour and 30 minutes | 2.50 [1.38 ; 3.63] |
| Day 22 |  |  |
|  | PREDOSE | 0.49 [-0.56 ; 1.55] |
|  | 40 minutes | 5.68 [4.51 ; 6.84] |
|  | 1 hour | 3.33 [2.18 ; 4.48] |
|  | 1 hour and 30 minutes | 2.32 [1.22 ; 3.43] |
| Day 25 |  |  |
|  | PREDOSE | 0.51 [-0.57 ; 1.59] |
|  | 40 minutes | 6.36 [5.16 ; 7.56] |
|  | 1 hour | 4.50 [3.34 ; 5.67] |
|  | 1 hour and 30 minutes | 2.45 [1.34 ; 3.55] |

|  | Pvalue |
| --- | --- |
| Interaction treatment*measure | < 0.0001 |

The interaction between treatment and measurement was significant at the 5% level. The effect of measurement time on diastolic blood pressure differed between groups.

Diastolic blood pressure per time

For each patient, the mean of D4 to D25 was taken for each time.

Figure 28: Evolution of diastolic blood pressure by measurement time and per day according to group

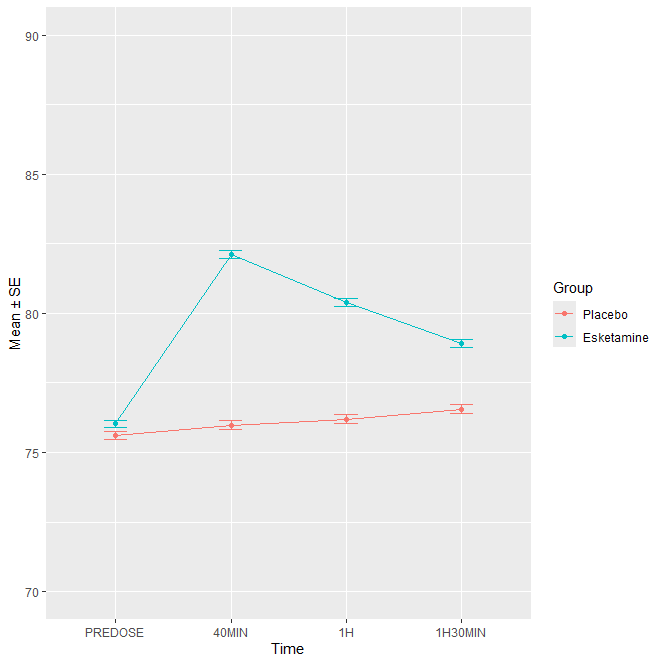

|  | Pvalue |
| --- | --- |
| Interaction treatment*time | < 0.0001 |

The interaction between treatment and measurement was significant at the 5% level. The effect of measurement time on diastolic blood pressure differed between groups.

Least-squares means were computed using the lsmeans function, with Tukey adjustment for multiple comparisons.

|  | Comparison | Estimate | Standard error | Pvalue |
| --- | --- | --- | --- | --- |
| Time : Predose | Esketamine vs Placebo | 0.52 | 0.57 | 0.3644 |
| Time : 40 minutes | Esketamine vs Placebo | 6.14 | 0.57 | < 0.0001 |
| Time : 1 hour | Esketamine vs Placebo | 4.22 | 0.57 | < 0.0001 |
| Time : 1 hour 30 | Esketamine vs Placebo | 2.41 | 0.57 | < 0.0001 |

Systolic blood pressure per day and per time

Figure 29: Evolution of systolic blood pressure by measurement time and per day according to group

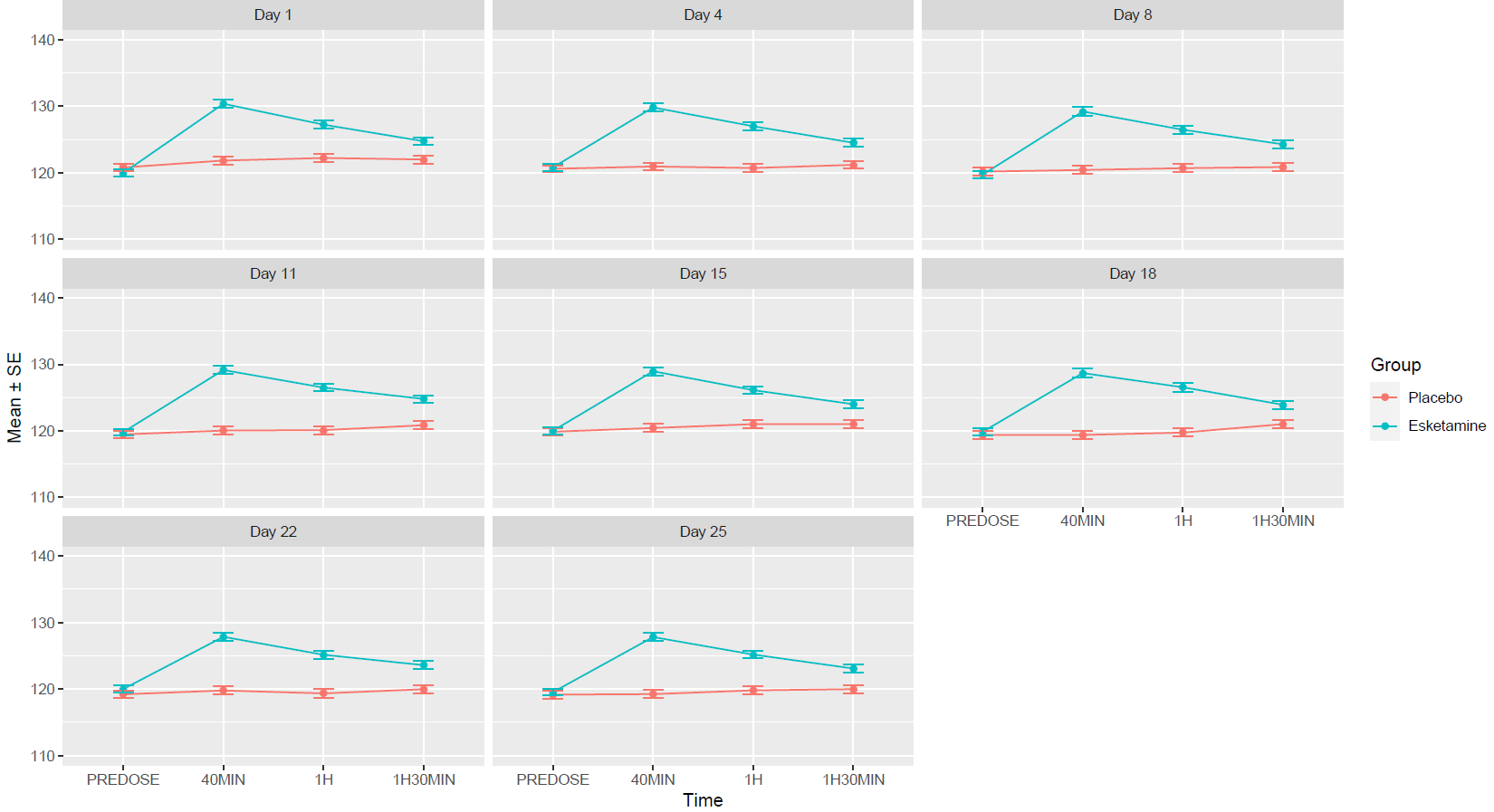

Table 17: Evolution of systolic blood pressure by measurement time and per day according to group, mean difference [Confidence Interval (CI) 95%]

| **Day** | **Time** | **Mean difference [CI95%]** |
| --- | --- | --- |
| Day 1 |  |  |
|  | PREDOSE | -0.83 [-2.31 ; 0.64] |
|  | 40 minutes | 8.54 [6.78 ; 10.30] |
|  | 1 hour | 5.01 [3.29 ; 6.74] |
|  | 1 hour and 30 minutes | 2.74 [1.14 ; 4.33] |
| Day 4 |  |  |
|  | PREDOSE | 0.22 [-1.25 ; 1.68] |
|  | 40 minutes | 8.88 [7.15 ; 10.60] |
|  | 1 hour | 6.28 [4.59 ; 7.96] |
|  | 1 hour and 30 minutes | 3.32 [1.69 ; 4.95] |
| Day 8 |  |  |
|  | PREDOSE | -0.46 [-2.04 ; 1.11] |
|  | 40 minutes | 8.80 [7.00 ; 10.59] |
|  | 1 hour | 5.78 [4.09 ; 7.47] |
|  | 1 hour and 30 minutes | 3.41 [1.75 ; 5.07] |
| Day 11 |  |  |
|  | PREDOSE | 0.34 [-1.13 ; 1.82] |
|  | 40 minutes | 9.10 [7.39 ; 10.82] |
|  | 1 hour | 6.40 [4.72 ; 8.07] |
|  | 1 hour and 30 minutes | 3.94 [2.30 ; 5.60] |
| Day 15 |  |  |
|  | PREDOSE | 0.14 [-1.38 ; 1.67] |
|  | 40 minutes | 8.53 [6.81 ; 10.26] |
|  | 1 hour | 5.12 [3.37 ; 6.86] |
|  | 1 hour and 30 minutes | 3.00 [1.34 ; 4.66] |
| Day 18 |  |  |
|  | PREDOSE | 0.46 [-1.03 ; 1.95] |
|  | 40 minutes | 9.30 [7.59 ; 11.02] |
|  | 1 hour | 6.82 [5.06 ; 8.58] |
|  | 1 hour and 30 minutes | 2.91 [1.22 ; 4.60] |
| Day 22 |  |  |
|  | PREDOSE | 0.80 [-0.70 ; 2.29] |
|  | 40 minutes | 8.07 [6.35 ; 9.80] |
|  | 1 hour | 5.81 [4.11 ; 7.51] |
|  | 1 hour and 30 minutes | 3.62 [1.97 ; 5.27] |
| Day 25 |  |  |
|  | PREDOSE | 0.38 [-1.11 ; 1.87] |
|  | 40 minutes | 8.58 [6.89 ; 10.27] |
|  | 1 hour | 5.38 [3.72 ; 7.05] |
|  | 1 hour and 30 minutes | 3.10 [1.51 ; 4.69] |

|  | Pvalue |
| --- | --- |
| Interaction treatment*measure | < 0.0001 |

The interaction between treatment and measurement was significant at the 5% level. The effect of measurement time on systolic blood pressure differed between groups.

Systolic blood pressure per time

For each patient, the mean of D4 to D25 was taken for each time.

Figure 30: Evolution of diastolic blood pressure by measurement time and per day according to group

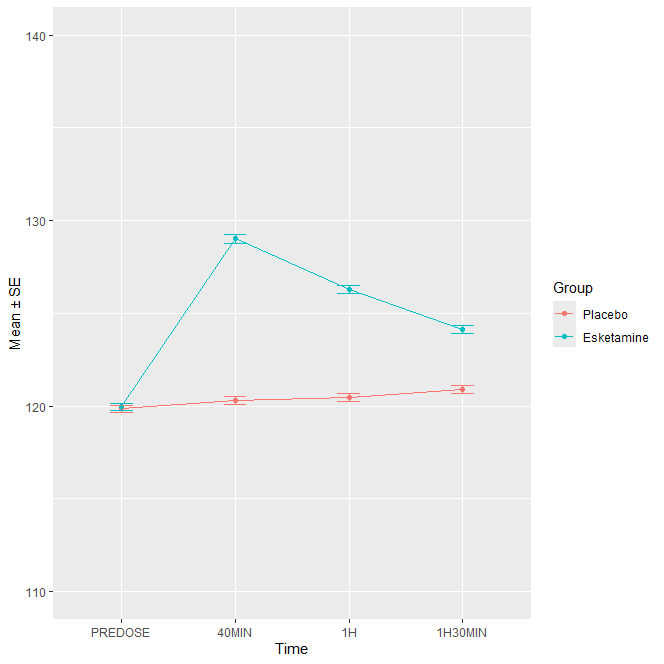

|  | Pvalue |
| --- | --- |
| Interaction treatment*time | < 0.0001 |

The interaction between treatment and measurement was significant at the 5% level. The effect of measurement time on systolic blood pressure differed between groups.

Least-squares means were computed using the lsmeans function, with Tukey adjustment for multiple comparisons.

|  | Comparison | Estimate | Standard error | Pvalue |
| --- | --- | --- | --- | --- |
| Time : Predose | Esketamine vs Placebo | 0.39 | 0.72 | 0.5881 |
| Time : 40 minutes | Esketamine vs Placebo | 8.96 | 0.72 | < 0.0001 |
| Time : 1 hour | Esketamine vs Placebo | 6.09 | 0.72 | < 0.0001 |
| Time : 1 hour 30 | Esketamine vs Placebo | 3.58 | 0.72 | < 0.0001 |

**Dissociation**

The dissociation criteria was analyzed using the Clinician-Administered Dissociative States Scale (CADSS). The CADSS consists of 23 subjective items. Participant’s responses are coded on a 5-point scale (0=not at all through to 4=extremely). To obtain the total CADSS score you need to sums all the items. The score vary from 0 to 92.

The CADSS was performed predose and at 40 minutes and 1.5 hours postdose. Predose no select. The maximum of the two values (40 minutes and 1.5 hours postdose) was selected for each day of double-blind induction phase, then the maximum value of phase was selected. Dissociation was defined if the CADSS value was ≥ 1.

Figure 31: Forest plot, dissociation outcome

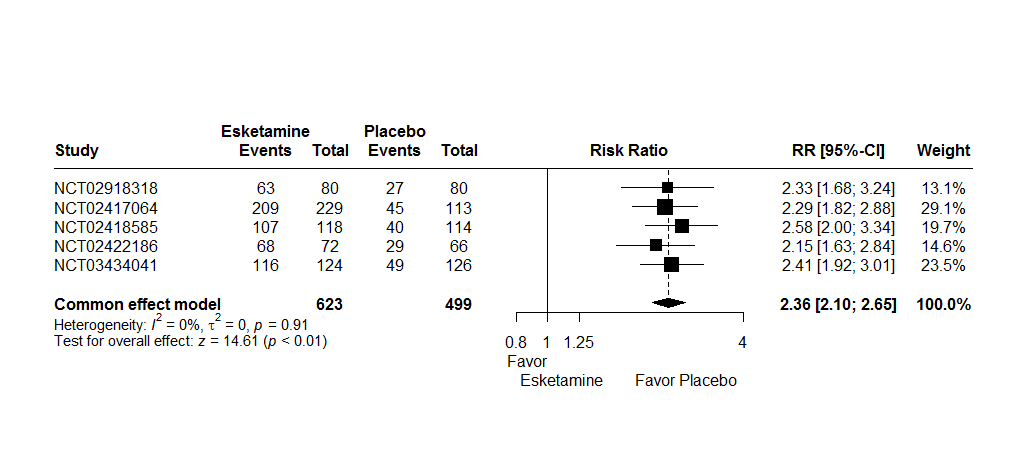

*RR : Relative Risk*

*CI : Confidence Interval*

A random effects model were not used because there are not heterogeneity between studies (I^2^ = 0%, t^2^ = 0, p = 0.91). The two-stage meta-analysis indicates a significant effect between dissociation and treatment group with an RR of 2.36 [2.10; 2.65], p < 0.01 in common effect model.

**Sedation**

The sedation criteria was analyzed using the Modified Observer’s Assessment of Alertness/Sedation (MOAA/S). The MOAA/S scores range from 0=no response to painful stimulus to 5=readily responds to name spoken in normal tone.

On each intranasal doing day, the MOAA/S was performed at every 15 minutes from predose to t=+1.5 hours postdose. Predose no select. The minimum of the values was selected for each day, then the minimal value for the treatment period was selected. Sedation was defined if the MOAA/S value was < 5.

Figure 32: Forest plot, sedation outcome

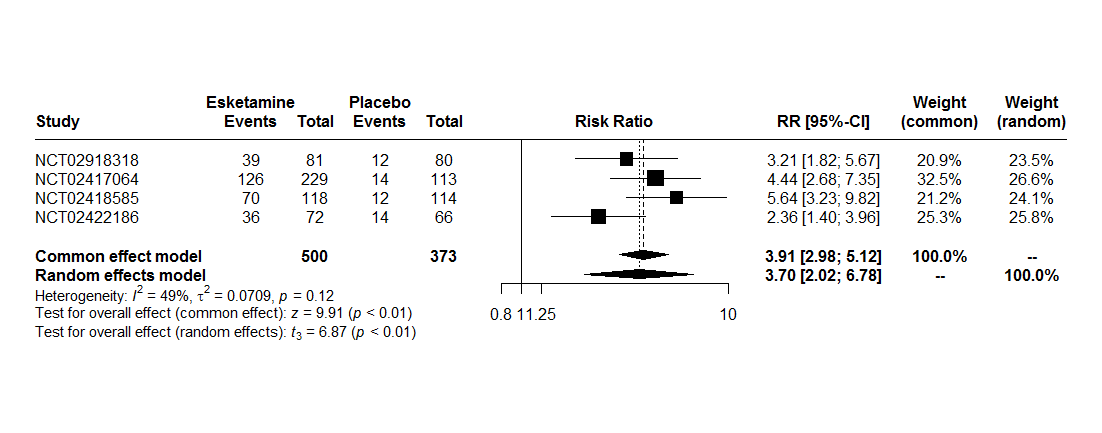
*RR : Relative Risk*

*CI : Confidence Interval*

A random effects model were used because there are heterogeneity between studies (I^2^ = 49%, t^2^ = 0.07, p = 0.12). The two-stage meta-analysis indicates a significant effect between dissociation and treatment group with an RR of 3.91 [2.98; 5.12], p < 0.01 in common effect model and an RR of 3.70 [2.02; 6.78], p < 0.01 in random effects model.

###### 3.1.2.2 Outcomes assessed at the last follow-up visit post-treatment

**Serious adverse events**

Figure 33: Forest plot, serious adverse events outcome at the follow-up visit

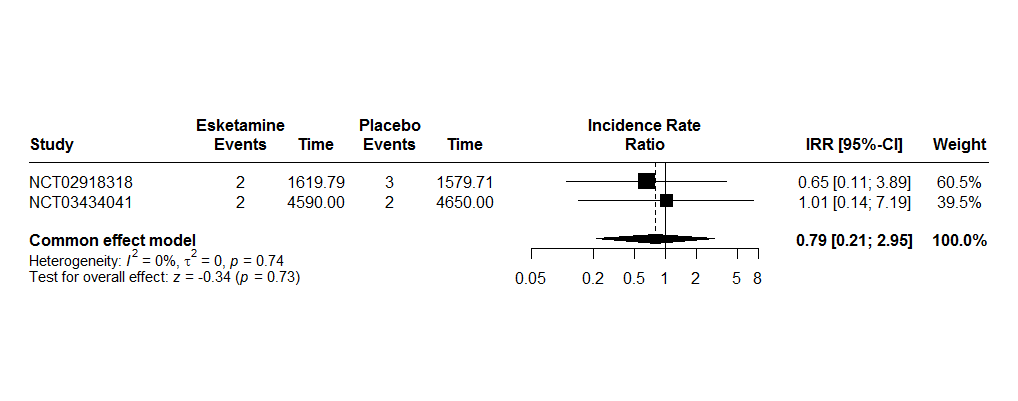

*IRR : Incidence Rate Ratio*

*CI : Confidence Interval*

A random effects model were not used because there are not heterogeneity between studies (I^2^ = 0%, t^2^ = 0, p = 0.74). The two-stage meta-analysis indicates a no significant effect between serious adverse events and treatment group with an IRR of 0.79 [0.21; 2.95], p = 0.73 in common effect model.

**Dropout for any cause**

Dropout for any cause is considered if patient checked at least one of the following criteria:

- Adverse event
- Death
- Lack of efficacy
- Lost to follow-up
- Non-compliance with switched oral antidepressant therapy
- Other
- Withdrawal by subject
- Investigator decision
- Protocol violation

Figure 34: Forest plot, dropout for any cause outcome during the follow-up period

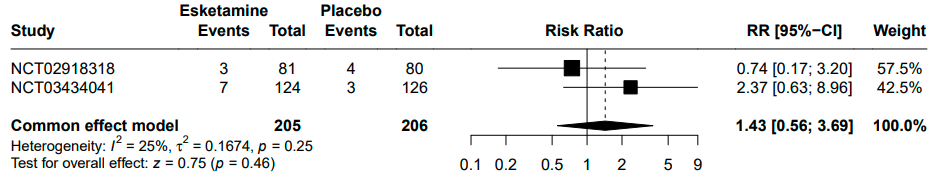

*RR : Relative Risk*

*CI : Confidence Interval*

A random effects model were not used because there are not heterogeneity between studies (I^2^ = 25%, t^2^ = 0.17, p = 0.25). The two-stage meta-analysis indicates a no significant effect between dropout for any cause and treatment group with an RR of 1.43 [0.56; 3.69], p = 0.46 in common effect model.

**Dropout due to adverse events**

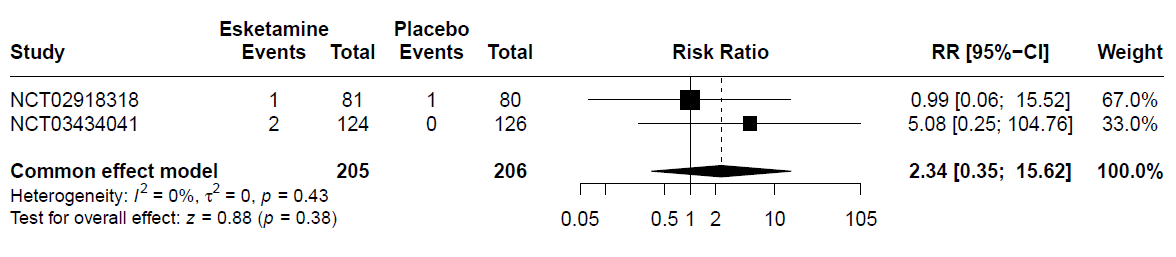
Figure 35: Forest plot, dropout due to adverse events outcome during the follow-up period

*RR : Relative Risk*

*CI : Confidence Interval*

A random effects model were not used because there are not heterogeneity between studies (I^2^ = 0%, t^2^ = 0, p = 0.43). The two-stage meta-analysis indicates a no significant effect between dropout due to adverse events and treatment group with an RR of 2.34 [0.35; 15.62], p = 0.38 in common effect model.

**Adverse events**

Figure 36: Forest plot, adverse events outcome at the follow-up visit

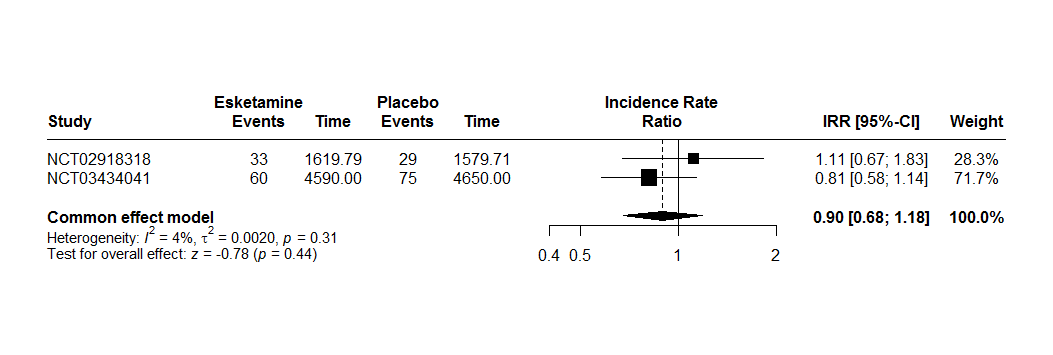
*IRR : Incidence Rate Ratio*

*CI : Confidence Interval*

A random effects model were not used because there are no heterogeneity between studies (I^2^ = 4%, t^2^ < 0.01, p = 0.31). The two-stage meta-analysis indicates a no significant effect between adverse events and treatment group with an IRR of 0.90 [0.68; 1.18], p = 0.44 in common effects.

**Blood pressure**

Only study NCT03434041 (3006) had available follow-up data for blood pressure.

Diastolic blood pressure

Figure 37: Evolution of diastolic blood pressure by week 2, 5 and 8 at the follow-up visit

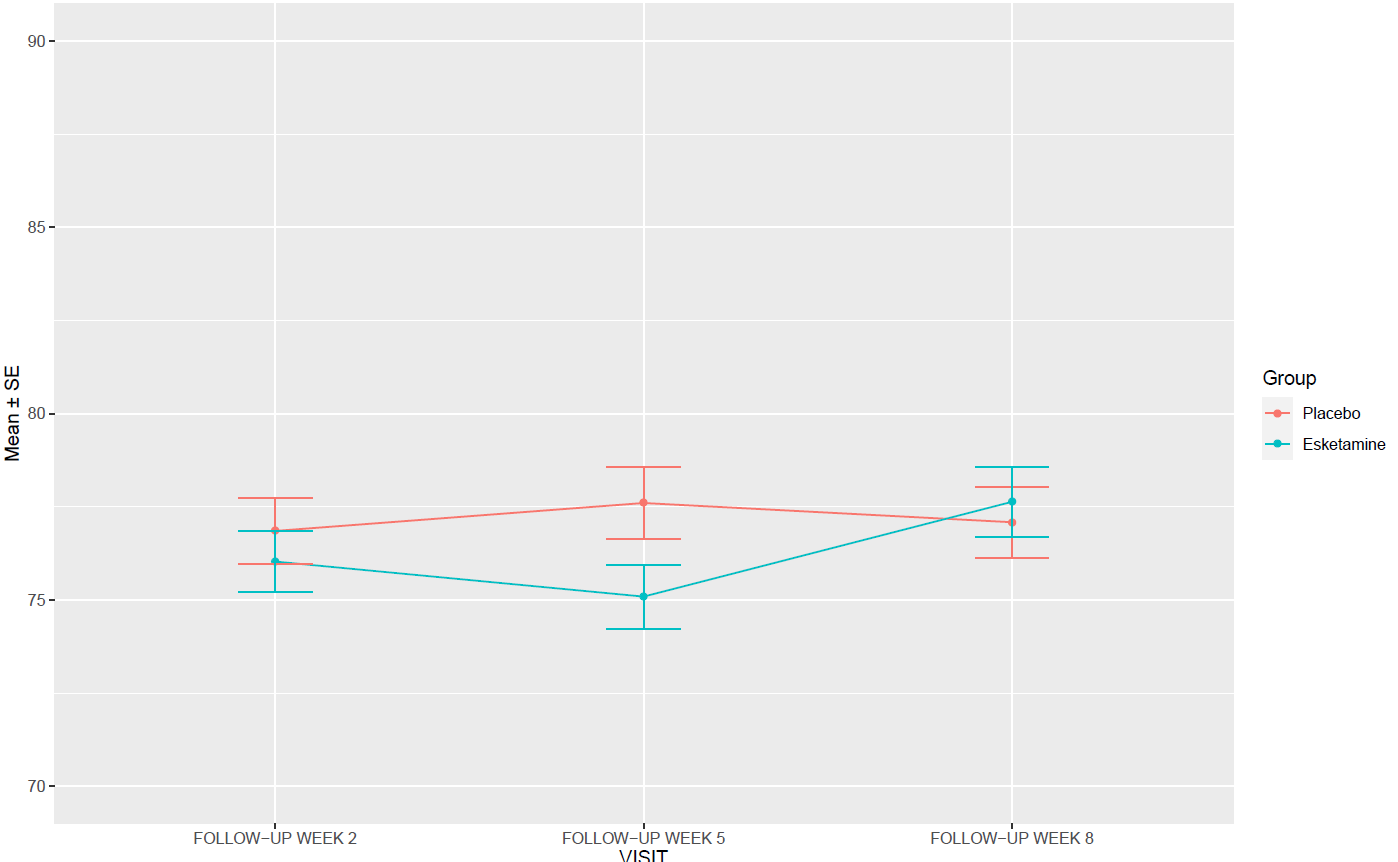

Table 18: Evolution of diastolic blood pressure by measurement time and per day according to group, mean difference [Confidence Interval(CI) 95%]

| **Day** | **Mean difference [CI95%]** |
| --- | --- |
| Follow-up week 2 | -0.82 [-3.20 ; 1.56] |
| Follow-up week 5 | -2.51 [-5.06 ; 0.04] |
| Follow-up week 8 | 0.55 [-2.06 ; 3.16] |

|  | Pvalue |
| --- | --- |
| Interaction treatment*follow-up visit | 0.2454 |

The interaction between treatment and follow-up visit was no significant at the 5% level.

Systolic blood pressure

Figure 38: Evolution of systolic blood pressure by week 2, 5 and 8 at the follow-up visit

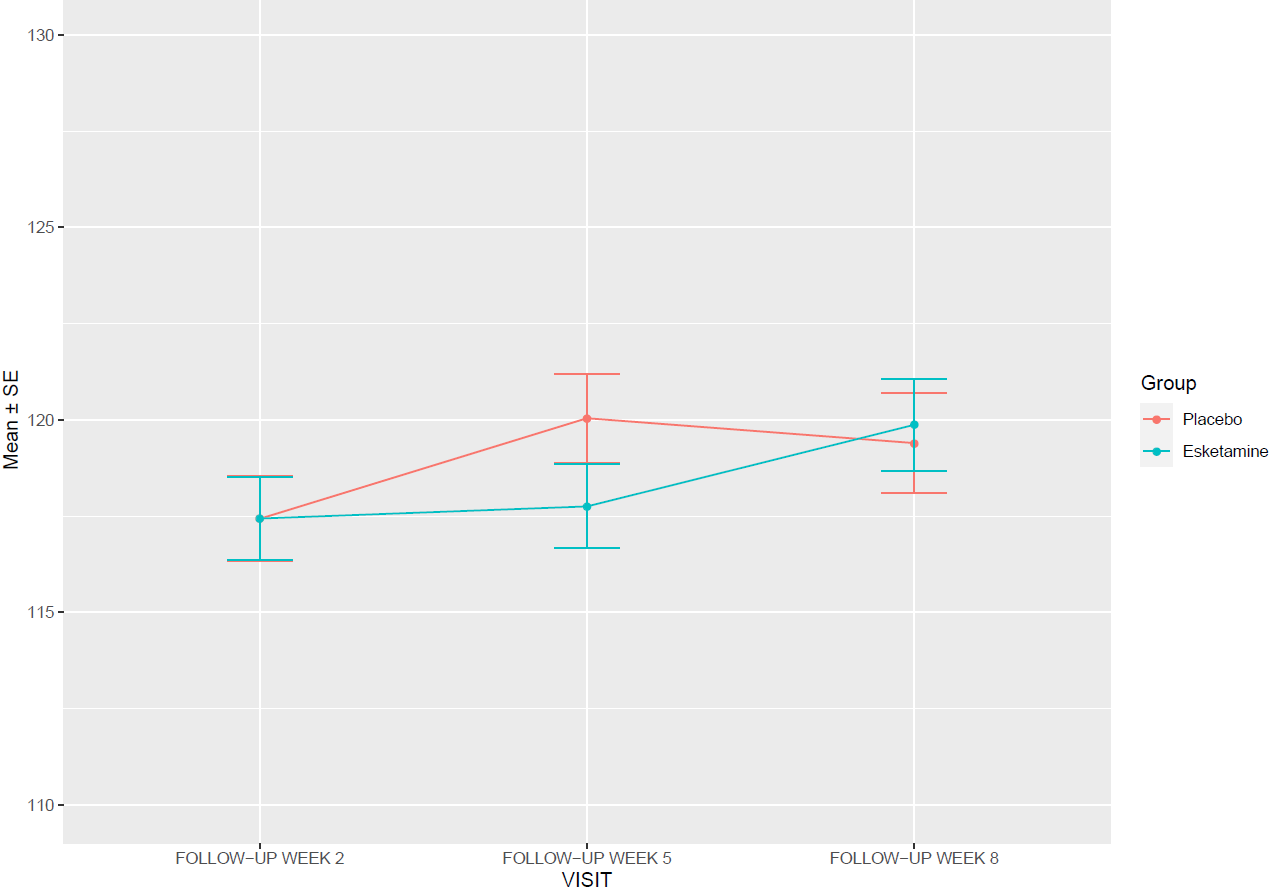

Table 19: Evolution of systolic blood pressure by measurement time and per day according to group, mean difference [Confidence Intervals (CI) 95%]

| **Day** | **Mean difference [CI95%]** |
| --- | --- |
| Follow-up week 2 | 0.01 [-3.02 ; 3.04] |
| Follow-up week 5 | -2.29 [-5.41 ; 0.83] |
| Follow-up week 8 | 0.47 [-2.98 ; 3.92] |

|  | Pvalue |
| --- | --- |
| Interaction treatment*visit | 0.4471 |

The interaction between treatment and visit was no significant at the 5% level.

##### 3.1.3 Moderators of esketamine efficacy

###### 3.1.3.1 Impact of resistance stage

Each participant was to be classified into the Thase and Rush resistance stages according to their basic characteristics (Table 7). However, some of patients have received treatments that do not fit neatly into the established categories of classification. A modified classification was applied (Table 8) to better capture these nuances. We used classification, but the pure number of attempts, irrespective of class was used (one class: stage I, two classes: stage II, three classes: stage III, four or more: stage IV.

Table 20: Definition classification Thase and Rush

| **Stage** | **Definition** |
| --- | --- |
| Stage I | Failure of 1 adequate trial of 1 major class of antidepressants |
| Stage II | Failure of 2 adequate trial of 2 major class of antidepressants |
| Stage III | Stage II resistance + failure of an adequate trial of a tricyclic agent |
| Stage IV | Stage III resistance + failure of an adequate trial of a monoamine oxidase inhibitor |
| Stage V | Stage IV resistance + a course of bilateral electroconvulsive therapy |

Table 21: Coding of classification Thase and Rush with the combined treatments

| **Traitement** | **STAGES** |
| --- | --- |
| MAOI | Stage 1 |
| MAOI + TRICYCLIC | Stage 2 |
| OTHER | Stage 1 |
| OTHER + MAOIS | Stage 2 |
| OTHER + TRICYCLIC | Stage 2 |
| SNRIS | Stage 1 |
| SNRIS + MAOI | Stage 2 |
| SNRIS + OTHER | Stage 2 |
| SNRIS + OTHER + MAOI + TRICYCLIC | Stage 4 |
| SNRIS + OTHER + MAOI | Stage 3 |
| SNRIS + OTHER + TRICYCLIC | Stage 3 |
| SNRIS + SSRIS | Stage 2 |
| SNRIS + SSRIS + MAOI | Stage 3 |
| SNRIS + SSRIS + OTHER | Stage 2 |
| SNRIS + SSRIS + OTHER + MAOI | Stage 4 |
| SNRIS + SSRIS + OTHER + MAOI + TRICYCLIC | Stage 4 |
| SNRIS + SSRIS + OTHER + TRICYCLIC | Stage 3 |
| SNRIS + SSRIS + TRICYCLIC | Stage 3 |
| SNRIS + TRICYCLIC | Stage 2 |
| SSRIS | Stage 1 |
| SSRIS + MAOI | Stage 2 |
| SSRIS + MAOI + TRICYCLIC | Stage 3 |
| SSRIS + OTHER | Stage 2 |
| SSRIS + OTHER + MAOI + TRICYCLIC | Stage 4 |
| SSRIS + OTHER + MAOI | Stage 3 |
| SSRIS + OTHER + TRICYCLIC | Stage 3 |
| SSRIS + TRICYCLIC | Stage 2 |
| TRICYCLIC | Stage 1 |

###### 3.1.3.1.1 Two-stage

**MADRS after at least 4 weeks**

Table 22: Distribution of the number of patients in each stage by study

|  | Number patients : Stage 1 | Number patients : Stage 2 | Number patients : Stage 3 | Number patients : Stage 4 |
| --- | --- | --- | --- | --- |
| NCT02918318 (2005) | 90 | 69 | 2 | 0 |
| NCT02417064 (3001) | 81 | 245 | 15 | 1 |
| NCT02418585 (3002) | 55 | 169 | 7 | 1 |
| NCT02422186 (3005) | 39 | 92 | 6 | 1 |
| NCT03434041 (3006) | 105 | 143 | 2 | 0 |
| All studies | 370 | 718 | 32 | 3 |

Figure 39: Forest plot, impact of resistance stage

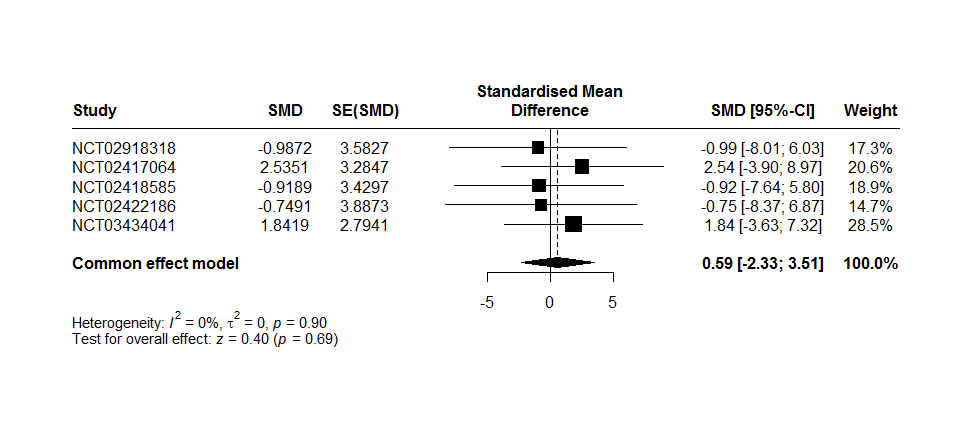

*SMD : The effect size estimates*

*SE(SMD) : Standard errors of the treatment effect estimates*

A random effects model were not used because there are no heterogeneity between studies (I2 = 0%, t^2^ = 0, p = 0.90). The two-stage meta-analysis indicates a no significant effect with MARDS at 4 weeks and interaction of Impact of resistance stage and treatment group with an SMD of 0.59 [-2.33; 3.51], p = 0.69 in common effects.

**MADRS after at least 4 weeks with imputation missing data**

Figure 40: Forest plot, impact of resistance stage with imputation missing data

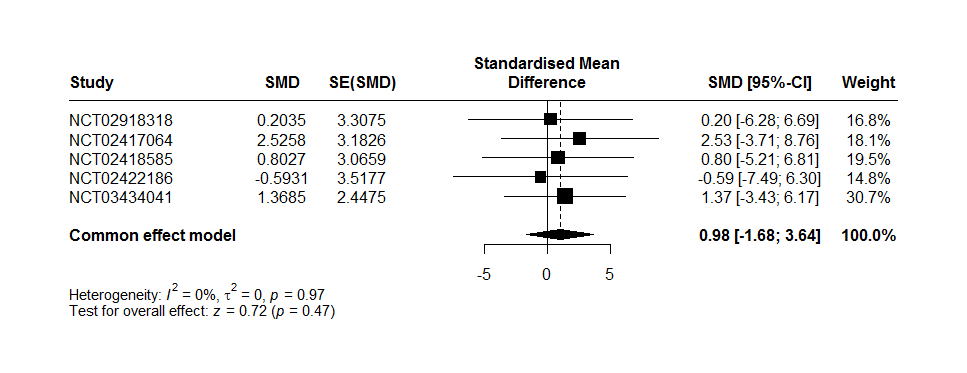

*SMD : The effect size estimates*

*SE(SMD) : Standard errors of the treatment effect estimates*.

A random effects model were not used because there are no heterogeneity between studies (I2 = 0%, t^2^ = 0, p = 0.97). The two-stage meta-analysis indicates a no significant effect with MARDS at 4 weeks with imputation data and interaction of Impact of resistance stage and treatment group with an SMD of 0.98 [-1.68; 3.64], p = 0.47 in common effects.

###### 3.1.3.1.2 One-stage

**MADRS after at least 4 weeks**

Table 23: Description MARDRS at 4 weeks by impact of resistance stage and group

| **Stage** | **Group** | **N** | **Mean ± standard deviation** |
| --- | --- | --- | --- |
| Stage 1 |  |  |  |
|  | Placebo | 147 | 23.38 ± 12.00 |
|  | Esketamine | 187 | 20.96 ± 12.87 |
| Stage 2 |  |  |  |
|  | Placebo | 296 | 24.72 ± 12.33 |
|  | Esketamine | 352 | 20.41 ± 12.43 |
| Stage 3 |  |  |  |
|  | Placebo | 9 | 22.44 ± 11.89 |
|  | Esketamine | 20 | 25.90 ± 13.62 |
| Stage 4 |  |  |  |
|  | Placebo | 1 | 33.00 |
|  | Esketamine | 1 | 38.00 |

|  | Pvalue |
| --- | --- |
| Interaction impact of resistance stage and treatment group | 0.8014 |

The one-stage meta-analysis indicates a no significant effect with MARDS at 4 weeks and interaction of Impact of resistance stage and treatment group at the 5% level.

**MADRS after at least 4 weeks with imputation missing data**

Table 24: Description MARDRS at 4 weeks with imputation missing data by impact of resistance stage and group

| **Stage** | **Group** | **N** | **Mean ± standard deviation** |
| --- | --- | --- | --- |
| Stage 1 |  |  |  |
|  | Placebo | 162 | 23.64 ± 11.68 |
|  | Esketamine | 208 | 21.41 ± 12.55 |
| Stage 2 |  |  |  |
|  | Placebo | 324 | 24.91 ± 12.06 |
|  | Esketamine | 394 | 21.04 ± 12.14 |
| Stage 3 |  |  |  |
|  | Placebo | 11 | 24.15 ± 11.48 |
|  | Esketamine | 21 | 26.50 ± 13.55 |
| Stage 4 |  |  |  |
|  | Placebo | 2 | 26.55 ± 9.12 |
|  | Esketamine | 1 | 38.00 |

|  | Pvalue |
| --- | --- |
| Interaction impact of resistance stage and treatment group | 0.5956 |

The one-stage meta-analysis indicates a no significant effect with MARDS at 4 weeks and interaction of Impact of resistance stage and treatment group at the 5% level.

###### 3.1.3.2 Sensitivity analysis impact of resistance stage

A sensitivity analysis was performed excluding patients with MAOI.

###### 3.1.3.2.1 Two-stage

**MADRS after at least 4 weeks**

Table 25: Distribution of the number of patients in each stage by study

|  | Number patients : Stage 1 | Number patients : Stage 2 | Number patients : Stage 3 |
| --- | --- | --- | --- |
| NCT02918318 (2005) | 90 | 69 | 2 |
| NCT02417064 (3001) | 81 | 243 | 11 |
| NCT02418585 (3002) | 54 | 168 | 5 |
| NCT02422186 (3005) | 39 | 92 | 4 |
| NCT03434041 (3006) | 105 | 143 | 1 |
| All studies | 369 | 715 | 23 |

Figure 41: Forest plot, impact of resistance stage

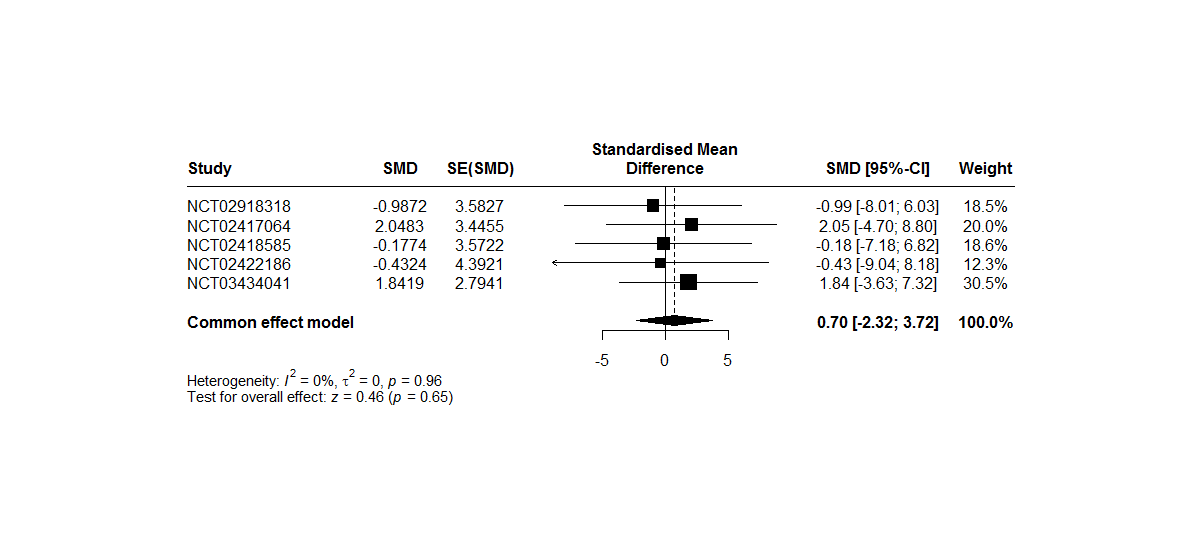

*SMD : The effect size estimates*

*SE(SMD) : Standard errors of the treatment effect estimates*

A random effects model were not used because there are no heterogeneity between studies (I2 = 0%, t^2^ = 0, p = 0.96). The two-stage meta-analysis indicates a no significant effect with MARDS at 4 weeks and interaction of impact of resistance stage and treatment group with an SMD of 0.70 [-2.32; 3.72], p = 0.65 in common effects.

**MADRS after at least 4 weeks with imputation missing data**

Figure 42: Forest plot, impact of resistance stage with imputation missing data

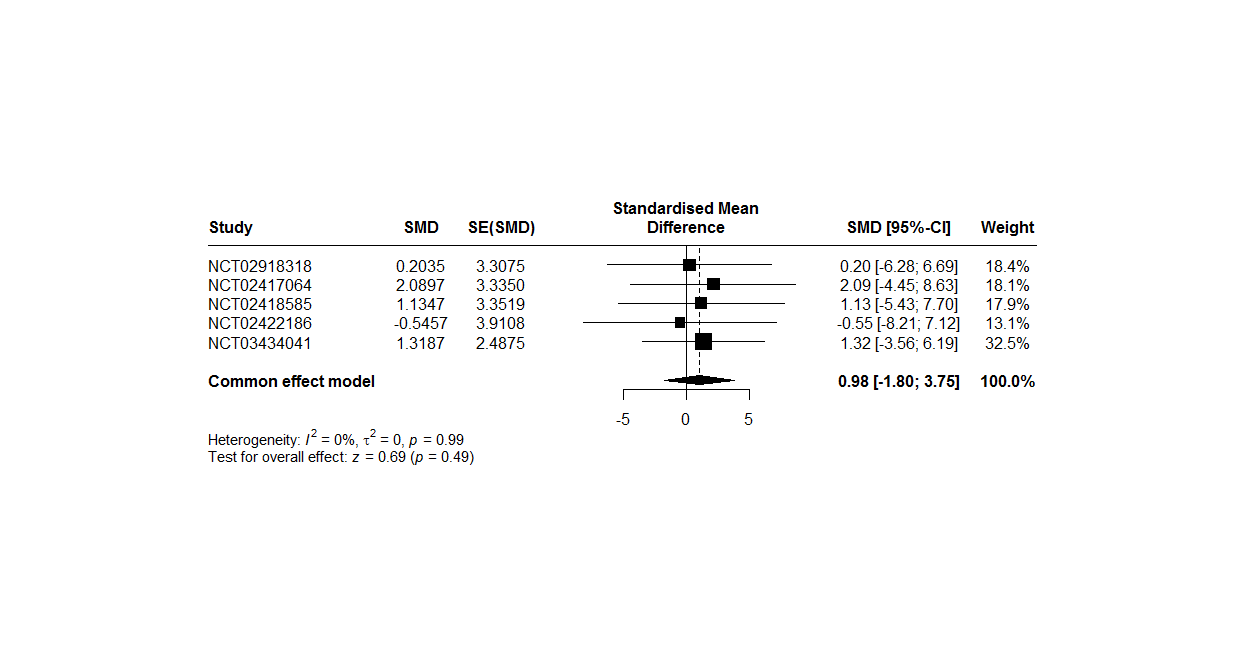

*SMD : The effect size estimates*

*SE(SMD) : Standard errors of the treatment effect estimates*.

A random effects model were not used because there are no heterogeneity between studies (I2 = 0%, t^2^ = 0, p = 0.99). The two-stage meta-analysis indicates a no significant effect with MARDS at 4 weeks with imputation data and interaction of Impact of resistance stage and treatment group with an SMD of 0.98 [-1.80; 3.75], p = 0.49 in common effects.

###### 3.1.3.2.2 One-stage

**MADRS after at least 4 weeks**

Table 26: Description MARDRS at 4 weeks by impact of resistance stage and group

| **Stage** | **Group** | **N** | **Mean ± standard deviation** |
| --- | --- | --- | --- |
| Stage 1 |  |  |  |
|  | Placebo | 146 | 23.40 ± 12.04 |
|  | Esketamine | 187 | 20.96 ± 12.87 |
| Stage 2 |  |  |  |
|  | Placebo | 296 | 24.72 ± 12.33 |
|  | Esketamine | 350 | 20.38 ± 12.44 |
| Stage 3 |  |  |  |
|  | Placebo | 7 | 19.71 ± 11.31 |
|  | Esketamine | 14 | 28.07 ± 12.98 |

|  | Pvalue |
| --- | --- |
| Interaction impact of resistance stage and treatment group | 0.8576 |

The one-stage meta-analysis indicates a no significant effect with MARDS at 4 weeks and interaction of Impact of resistance stage and treatment group at the 5% level.

**MADRS after at least 4 weeks with imputation missing data**

Table 27: Description MARDRS at 4 weeks with imputation missing data by impact of resistance stage and group

| **Stage** | **Group** | **N** | **Mean ± standard deviation** |
| --- | --- | --- | --- |
| Stage 1 |  |  |  |
|  | Placebo | 161 | 23.66 ± 11.71 |
|  | Esketamine | 208 | 21.41 ± 12.55 |
| Stage 2 |  |  |  |
|  | Placebo | 324 | 24.91 ± 12.06 |
|  | Esketamine | 391 | 21.02 ± 12.17 |
| Stage 3 |  |  |  |
|  | Placebo | 8 | 21.81 ± 12.04 |
|  | Esketamine | 15 | 28.76 ± 12.79 |

|  | Pvalue |
| --- | --- |
| Interaction impact of resistance stage and treatment group | 0.6958 |

The one-stage meta-analysis indicates a no significant effect with MARDS at 4 weeks and interaction of Impact of resistance stage and treatment group at the 5% level.

###### 3.1.3.3 Impact of age

###### 3.1.3.3.1 Two-stage

**MADRS after at least 4 weeks**

Table 28: Number subject per trial per group

| **Number NCT trial** | **Group** | **N** |
| --- | --- | --- |
| NCT02918318 | Placebo | 72 |
|  | Esketamine | 73 |
| NCT02417064 | Placebo | 108 |
|  | Esketamine | 210 |
| NCT02418585 | Placebo | 106 |
|  | Esketamine | 105 |
| NCT02422186 | Placebo | 60 |
|  | Esketamine | 63 |
| NCT03434041 | Placebo | 107 |
|  | Esketamine | 109 |

Figure 43: Forest plot, impact of age

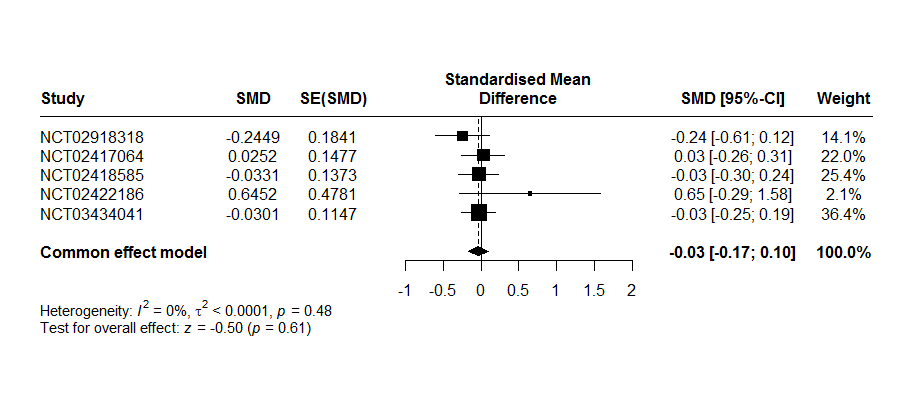
*SMD : The effect size estimates*

*SE(SMD) : Standard errors of the treatment effect estimates.*

A random effects model were not used because there are no heterogeneity between studies (I^2^ = 0%, t^2^ < 0.01, p = 0.48). The two-stage meta-analysis indicates a no significant effect with MARDS at 4 weeks and interaction of impact of age and treatment group with an SMD of -0.03 [-0.17; 0.10], p = 0.61 in common effects.

**MADRS after at least 4 weeks with imputation missing data**

Table 29: Number subject per trial per group

| **Number NCT trial** | **Group** | **N** |
| --- | --- | --- |
| NCT02918318 (2005) | Placebo | 80 |
|  | Esketamine | 81 |
| NCT02417064 (3001) | Placebo | 113 |
|  | Esketamine | 229 |
| NCT02418585 (3002) | Placebo | 114 |
|  | Esketamine | 118 |
| NCT02422186 (3005) | Placebo | 66 |
|  | Esketamine | 72 |
| NCT03434041 (3006) | Placebo | 126 |
|  | Esketamine | 124 |

Figure 44: Forest plot, impact of age with imputation missing data for MADRS

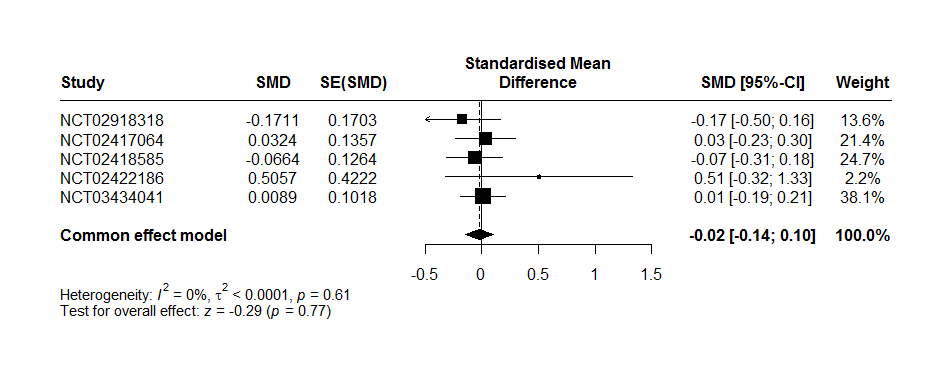

*SMD : The effect size estimates*

*SE(SMD) : Standard errors of the treatment effect estimates.*

A random effects model were not used because there are no heterogeneity between studies (I^2^ = 0%, t^2^ <0.01, 0, p = 0.61). The two-stage meta-analysis indicates a no significant effect with MARDS at 4 weeks with imputation data and interaction of impact of age and treatment group with an SMD of -0.02 [-0.14; 0.10], p = 0.77 in common effects.

###### 3.1.3.3.2 One-stage

**MADRS after at least 4 weeks**

|  | Pvalue |
| --- | --- |
| Interaction impact of age and treatment group | 0.4456 |

The one-stage meta-analysis indicates a no significant effect with MARDS at 4 weeks and interaction of Impact of age and treatment group at the 5% level.

Table 30: Description MARDRS at 4 weeks by age and group

| **Stage** | **Group** | **N** | **Mean ± standard deviation** |
| --- | --- | --- | --- |
| <40 |  |  |  |
|  | Placebo | 137 | 24.50 ± 11.34 |
|  | Esketamine | 202 | 21.65 ± 12.28 |
| [40-50] |  |  |  |
|  | Placebo | 122 | 22.04 ± 12.78 |
|  | Esketamine | 132 | 19.52 ± 12.40 |
| ]50-60] |  |  |  |
|  | Placebo | 100 | 24.13 ± 13.27 |
|  | Esketamine | 128 | 18.75 ± 12.41 |
| ]60-70] |  |  |  |
|  | Placebo | 77 | 26.42 ± 10.70 |
|  | Esketamine | 69 | 21.20 ± 13.50 |
| >70 |  |  |  |
|  | Placebo | 17 | 29.29 ± 12.49 |
|  | Esketamine | 29 | 29.21 ± 11.99 |

**MADRS after at least 4 weeks with imputation missing data**

|  | Pvalue |
| --- | --- |
| Interaction impact of age and treatment group | 0.6052 |

The one-stage meta-analysis indicates a no significant effect with MARDS at 4 weeks and interaction of Impact of age and treatment group at the 5% level.

Table 31: Description MARDRS at 4 weeks with imputation missing data by age and group

| **Stage** | **Group** | **N** | **Mean ± standard deviation** |
| --- | --- | --- | --- |
| <40 |  |  |  |
|  | Placebo | 163 | 24.98 ± 10.92 |
|  | Esketamine | 224 | 21.98 ± 11.93 |
| [40-50] |  |  |  |
|  | Placebo | 129 | 22.01 ± 12.61 |
|  | Esketamine | 146 | 20.14 ± 12.30 |
| ]50-60] |  |  |  |
|  | Placebo | 104 | 24.12 ± 13.07 |
|  | Esketamine | 141 | 19.37 ± 12.12 |
| ]60-70] |  |  |  |
|  | Placebo | 83 | 26.45 ± 10.37 |
|  | Esketamine | 78 | 21.85 ± 13.06 |
| >70 |  |  |  |
|  | Placebo | 20 | 30.21 ± 11.73 |
|  | Esketamine | 35 | 29.71 ± 11.40 |

###### 3.1.3.4 Impact of per capita gross national income

Table 32: Country classification

|  | **High income country** | **Low, lower middle, and upper middle income country** |
| --- | --- | --- |
| China |  | **X** |
| USA | **X** |  |
| Africa |  | **X** |
| Europe | **X** |  |
| North America | **X** |  |
| South America |  | **X** |
| Czech Republic | **X** |  |
| Germany | **X** |  |
| Spain | **X** |  |
| Poland | **X** |  |
| Belgium | **X** |  |
| Brazil |  | **X** |
| Canada | **X** |  |
| Estonia | **X** |  |
| France | **X** |  |
| Hungary | **X** |  |
| Ital | **X** |  |
| Mexico |  | **X** |
| Slovakia | **X** |  |
| Sweden | **X** |  |
| Turkey |  | **X** |
| Japan | **X** |  |

Table 33: Description of the per capita gross national income in each initiation study

| **Study** | **High income country** | **Low, lower middle, and upper middle income country** |
| --- | --- | --- |
| NCT02918318 (2005) | 161 | 0 |
| NCT02417064 (3001) | 240 | 102 |
| NCT02418585 (3002) | 232 | 0 |
| NCT02422186 (3005) | 130 | 8 |
| NCT03434041 (3006) | 28 | 222 |

###### 3.1.3.4.1 Two-stage

Three country were evaluated to two-stage analysis because two studies had no low, lower middle or upper middle income countries.

**MADRS after at least 4 weeks**

Figure 45: Forest plot, impact of per capita gross national income

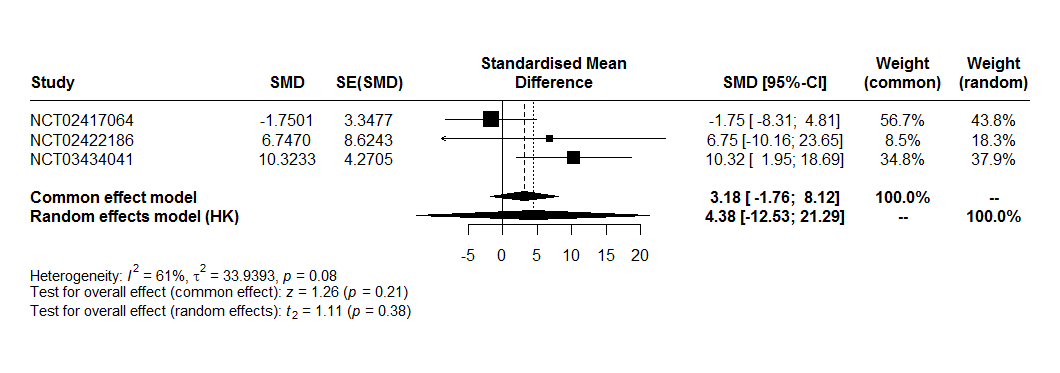
*SMD : The effect size estimates*

*SE(SMD) : Standard errors of the treatment effect estimates*

Interpretation:

A random effects model were used because there are heterogeneity between studies (I^2^ = 61%, t^2^ = 33.94, p = 0.08). The two-stage meta-analysis indicates a no significant effect with MARDS at 4 weeks and interaction of impact of per capital gross national income and treatment group with an SMD of 3.218 [-1.76; 8.12], p = 0.21 in common effect and an SMD of 4.39 [-12.53; 21.29], p = 0.38 in random effects.

**MADRS after at least 4 weeks with imputation missing data**

Figure 46: Forest plot, impact of per capita gross national income with imputation missing data for MADRS

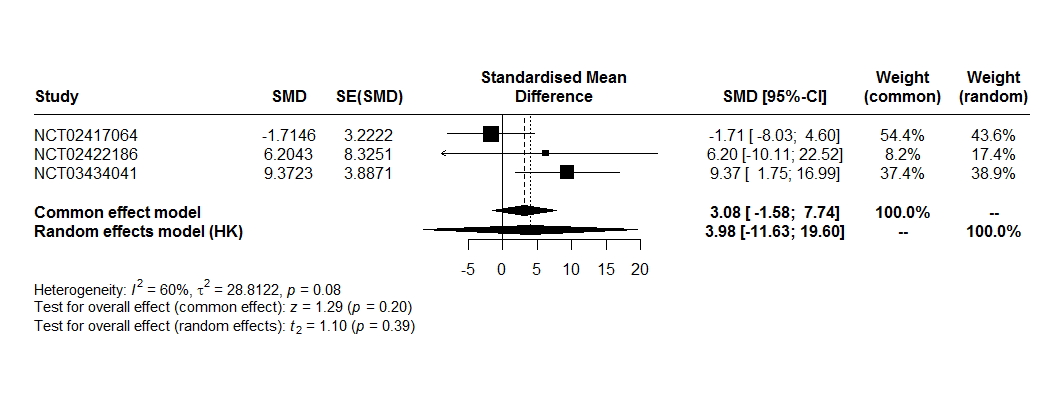
*SMD : The effect size estimates*

*SE(SMD) : Standard errors of the treatment effect estimates.*

A random effects model were used because there are heterogeneity between studies (I^2^ = 60%, t^2^=28.81, p = 0.08). The two-stage meta-analysis indicates a no significant effect with MARDS at 4 weeks with imputation data and interaction of impact of per capital gross national income and treatment group with an SMD of 3.08 [-1.58; 7.74], p=0.08 in common effect and an SMD of 3.98 [-11.63; 19.60], p=0.39 in random effects.

###### 3.1.3.4.2 One-stage

**MADRS after at least 4 weeks**

Table 34: Description MARDRS at 4 weeks by impact of capita gross national income

| **Stage** | **Group** | **N** | **Mean ± standard deviation** |
| --- | --- | --- | --- |
| **High income country** |  |  |  |
|  | Placebo | 321 | 24.08 ± 12.22 |
|  | Esketamine | 395 | 20.68 ± 12.67 |
| **Low, lower middle, and upper middle income country** |  |  |  |
|  | Placebo | 132 | 24.70 ± 12.19 |
|  | Esketamine | 165 | 21.16 ± 12.62 |

|  | Pvalue |
| --- | --- |
| Interaction impact of capita gross national income and treatment group | 0.3079 |

The one-stage meta-analysis indicates a no significant effect with MARDS at 4 weeks and interaction of impact of per capital gross national income and treatment group at the 5% level.

**MADRS after at least 4 weeks with imputation missing data**

Table 35: Description MARDRS at 4 weeks with imputation missing data by impact of capita gross national income

| **Stage** | **Group** | **N** | **Mean ± standard deviation** |
| --- | --- | --- | --- |
| **High income country** |  |  |  |
|  | Placebo | 349 | 24.07 ± 11.97 |
|  | Esketamine | 442 | 21.14 ± 12.36 |
| **Low, lower middle, and upper middle income country** |  |  |  |
|  | Placebo | 150 | 25.46 ± 11.71 |
|  | Esketamine | 182 | 21.94 ± 12.35 |

|  | Pvalue |
| --- | --- |
| Interaction impact of capita gross national income and treatment group | 0.3557 |

The one-stage meta-analysis indicates a no significant effect with MARDS at 4 weeks with imputation missing data and interaction of impact of per capital gross national income and treatment group at the 5% level.

#### 3.2 Continuation studies

##### 3.2.1 Efficacy outcomes assessed at the end of the study

Only one continuation study.

Patients randomized in the maintenance phase were included in the meta-analysis. A total of 303 patients were randomized, but 6 were excluded due to sponsor audit results. The analysis was carried out on 297 patients (176 in a stable remission and 121 in a stable response).

The end of study corresponds to the end of maintenance phase.

For all outcome, the definitions were the same that initiations studies.

**Relapse**

| **Treatment** | **HR [95%CI]** | **Standard error** |
| --- | --- | --- |
| Placebo | 1 |  |
| Esketamine | 0.38 [0.26, 0.57] | 0.20 |

Relapse for patient in stable remission:

| **Treatment** | **HR [95%CI]** | **Standard error** |
| --- | --- | --- |
| Placebo | 1 |  |
| Esketamine | 0.46 [0.28, 0.77] | 0.26 |

Relapse for patient in stable response:

| **Treatment** | **HR [95%CI]** | **Standard error** |
| --- | --- | --- |
| Placebo | 1 |  |
| Esketamine | 0.30 [0.16, 0.55] | 0.31 |

**Suicide/suicide attempt**

| **Treatment** | **Number event** | **Number total of subjects** |
| --- | --- | --- |
| Placebo | 0 | 145 |
| Esketamine | 2 | 152 |

Table 36: Describe Suicide/Suice attempt

| Patient 1 | Stable response | Esketamine group | C-SSRS part Suicidal behavior were selected. Time relapse = 48 days |
| --- | --- | --- | --- |
| Patient 2 | Stable response | Esketamine group | C-SSRS part Suicidal behavior were selected. Time relapse = 262.96 days |

**Suicidal ideations**

| **Treatment** | **Number event** | **Number total of subjects** |
| --- | --- | --- |
| Placebo | 0 | 145 |
| Esketamine | 1 | 152 |

**MADRS**

Selection of last MADRS available in maintenance phase by patient.

|  | ESKETAMINE | Placebo |
| --- | --- | --- |
| n | 151 | 145 |
| Mean | 12.89 | 19.07 |
| Standard deviation | 11.66 | 12.61 |

**Remission**

| **Treatment** | **Number of event** | **Number total of subjects** |
| --- | --- | --- |
| Placebo | 51 | 145 |
| Esketamine | 86 | 151 |

**Sheehan Disability Scale**

|  | ESKETAMINE | Placebo |
| --- | --- | --- |
| n | 150 | 144 |
| Mean | 7.32 | 11.04 |
| Standard deviation | 7.28 | 8.58 |

**PHQ-9**

|  | ESKETAMINE | Placebo |
| --- | --- | --- |
| n | 150 | 144 |
| Mean | 5.73 | 8.12 |
| Standard deviation | 5.47 | 6.52 |

##### 3.2.2 Safety outcomes assessed at the end of the study

**Serious adverse events**

|  | ESKETAMINE | Placebo |
| --- | --- | --- |
| Number of events | 5 | 1 |
| Person time at risk (days) | 24145.58 | 16123.33 |
| Incidence rate [CI95%] per 1000/person day | 0.2071 [0.2013 ; 0.2128] | 0.0620 [0.0562 ; 0.0677] |

Result poisson regression. An overdispersion was observed, and a quasi-Poisson model was performed.

| Estimate (reference = Placebo) | Standard error | P-value |
| --- | --- | --- |
| 1.206 | 2.06 | 0.5590 |

Incidence Risk Ratio and 95% Confidence interval = 3.34 [0.42; 26.21]

**Dropout for any cause**

|  | ESKETAMINE | Placebo |
| --- | --- | --- |
| Number of events | 12 | 11 |
| Number total of subjects | 152 | 145 |

Odds ratio and 95% confidence interval (reference: Placebo) = 1.04 [0.44; 2.48]

Relative risk and 95% confidence interval (reference: Placebo) = 1.04 [0.47; 2.28]

**Dropout due to adverse events**

|  | ESKETAMINE | Placebo |
| --- | --- | --- |
| Number of events | 1 | 2 |
| Number total of subjects | 152 | 145 |

Odds ratio and 95% confidence interval (reference: Placebo) = 0.47 [0.02; 4.99]

Relative risk and 95% confidence interval (reference: Placebo) = 0.48 [0.04; 5.20]

**Adverse events**

|  | ESKETAMINE | Placebo |
| --- | --- | --- |
| Number of events | 3263 | 400 |
| Person time at risk (days) | 24145.58 | 16123.33 |
| Incidence rate [CI95%] per 1000/person day | 135.14 [134.99 ; 135.29] | 24.81 [24.73 ; 24.89] |

Result poisson regression. An overdispersion was observed, and a quasi-Poisson model was performed.

| Estimate (reference = Placebo) | Standard error | P-value |
| --- | --- | --- |
| 1.695 | 0.26 | < 0.0001 |

Incidence Risk Ratio and 95% Confidence interval = 5.45 [4.20; 7.06]

**Blood pressure**

The blood pressure was measured at predose and at 40 minutes, 1 hour and 1.5 hours postdose and repeated during the maintenance phase.

A linear mixed model was realized with blood pressure as response variable, treatment and measurement interaction (predose and at 40 minutes, 1 hour and 1.5 hours postdose) and day as random effect.

Diastolic blood pressure

|  | Pvalue |
| --- | --- |
| Interaction treatment*measure | < 0.0001 |

The interaction between treatment and measurement was significant at the 5% level. The effect of measurement time on diastolic blood pressure differed between groups.

Systolic blood pressure

|  | Pvalue |
| --- | --- |
| Interaction treatment*measure | < 0.0001 |

The interaction between treatment and measurement was significant at the 5% level. The effect of measurement time on systolic blood pressure differed between groups.

**Dissociation**

|  | ESKETAMINE | Placebo |
| --- | --- | --- |
| Number of events | 118 | 27 |
| Number total of subjects | 151 | 143 |

Odds ratio and 95% confidence interval (reference: Placebo) = 15.36 [8.83; 27.63]

Relative risk and 95% confidence interval (reference: Placebo) = 4.14 [2.92; 5.87]

**Sedation**

|  | ESKETAMINE | Placebo |
| --- | --- | --- |
| Number of events | 66 | 22 |
| Number total of subjects | 152 | 145 |

Odds ratio and 95% confidence interval (reference: Placebo) = 4.29 [2.49; 7.61]

Relative risk and 95% confidence interval (reference: Placebo) = 2.86 [1.87; 4.38]

##### 3.2.3 Moderators of esketamine efficacy

**Impact of resistance stage**

|  | Interaction term between treatment group and Thase and Rush classification |
| --- | --- |
| Estimate | -4.12 |
| Standard error | 3.52 |
| pvalue | 0.211 |
| Number of subjects | 181 |

**Impact of age**

|  | Interaction term between treatment group and age |
| --- | --- |
| Estimate | 0.33 |
| Standard error | 0.13 |
| pvalue | 0.0079 |
| Number of subjects | 296 |

**Impact of country**

|  | Interaction term between treatment group and country |
| --- | --- |
| Estimate | 5.01 |
| Standard error | 3.49 |
| pvalue | 0.151 |
| Number of subjects | 296 |

**Analysis primary outcome with one country removed**

**Global population, forest plot:**

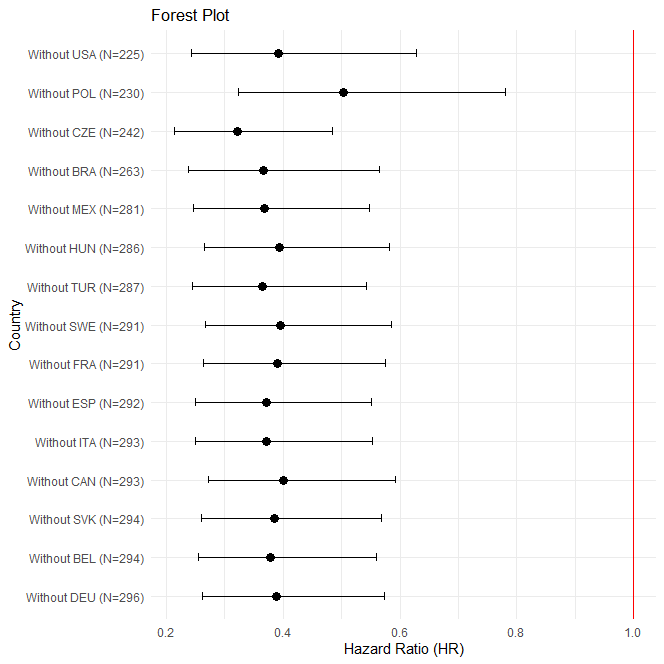

**Stable remission population, forest plot:**

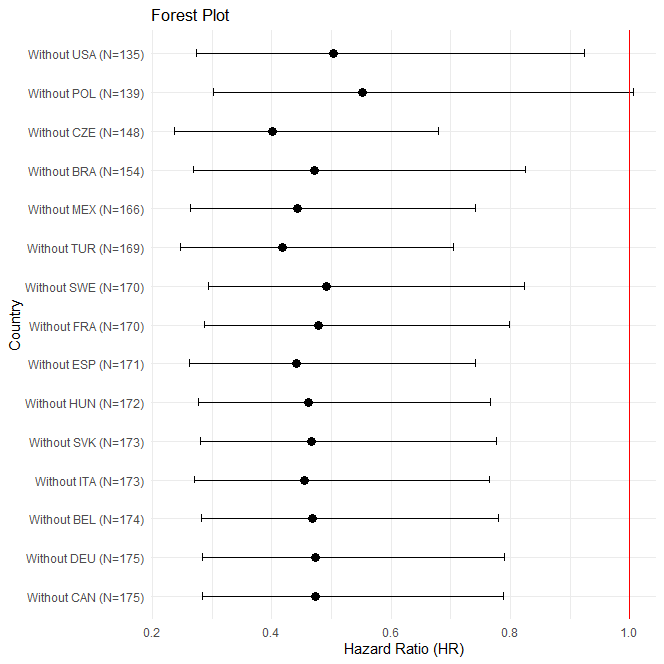

**Stable response population:**

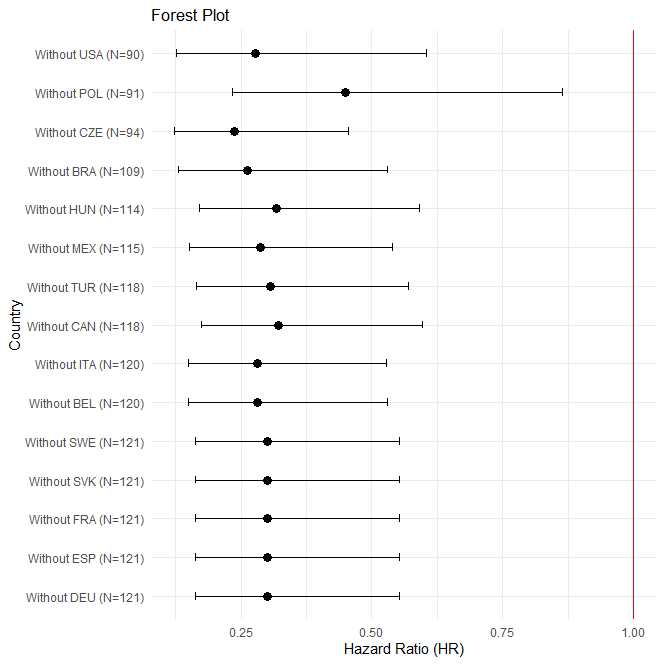
